# Behavioral nudges increase COVID-19 vaccinations: Two randomized controlled trials

**DOI:** 10.1101/2021.04.12.21254876

**Authors:** Hengchen Dai, Silvia Saccardo, Maria A Han, Lily Roh, Naveen Raja, Sitaram Vangala, Hardikkumar Modi, Shital Pandya, Daniel M Croymans

**Affiliations:** Anderson School of Management, University of California Los Angeles, Los Angeles, California, 90095. USA; Department of Social and Decision Sciences, Carnegie Mellon University, Pittsburgh, Pennsylvania, 15213. USA; Department of Medicine, David Geffen School of Medicine, University of California Los Angeles, Los Angeles, California, 90095. USA; Office of Population Health and Accountable Care, University of California Los Angeles, 10960 Wilshire Blvd, Los Angeles, 90095. USA; Department of Medicine Statistics Core, David Geffen School of Medicine, 1100 Glendon Avenue, Los Angeles, CA 90024, USA; Office of Health Informatics and Analytics, University of California Los Angeles Health Sciences, Los Angeles, 90095. USA

## Abstract

Fighting the COVID-19 pandemic requires quick and effective strategies to maximize vaccine uptake. We present two sequential randomized controlled trials (RCTs) that tackle this challenge with behavioral science insights. We deliver text-based nudges to UCLA Health patients one day (first RCT; N=113,229) and eight days (second RCT; N=90,662) after they receive notifications of vaccine eligibility. In the first RCT, text messages designed to make vaccination salient and easy to schedule boost appointment and vaccination rates by 86% and 26%, respectively. Nudges that make patients feel endowed with the vaccine heighten these effects, but addressing vaccine hesitancy via a video-based information intervention does not yield benefits beyond simple text. These results hold across ethnicity and age groups. By contrast, online experiments (N=2,003) soliciting hypothetical responses to the same messages reveal the opposite patterns, underscoring the importance of pilot-testing behavioral nudges in the real world before scaling them up. In the second RCT, we further find that receiving a second reminder boosts appointment and vaccination rates by 52% and 16%, respectively. Our findings suggest that text-based nudges can substantially increase and accelerate COVID-19 vaccinations at almost zero marginal cost, highlighting the promising role of behavioral science in addressing a critical component of the COVID-19 pandemic response.

## Introduction

Ending the COVID-19 health crisis requires vaccinating as many people as possible to quickly reach the herd immunity threshold (1). Enormous efforts and investments have ensured the development, authorization, and mass-production of safe and effective SARS-CoV-2 vaccines in record timing (2,3). Yet, potential recipients’ reluctance and failure to get inoculated threatens to undermine these efforts. As every American is promised to have access to the vaccine by May 1, 2021, developing evidence-based communication strategies to maximize vaccine uptake at scale is a pressing task (4). Nudges (i.e., interventions that incorporate behavioral science insights), which have been successfully deployed for behavior change across numerous policy-relevant domains (5–9), can contribute to this effort (4,10).

Prior work suggests two major approaches for increasing vaccination: shifting intentions and facilitating action (11). The former approach involves first identifying beliefs and misconceptions that drive vaccine hesitancy and then designing information interventions to tackle these drivers and thus boost *intentions* to receive the vaccine (11). This approach is worth exploring given that only about 60% of Americans reported planning to get a COVID-19 vaccine as of February 2021 (12) and that widespread misinformation further contributes to vaccine hesitancy (13). Yet, changing intentions alone is not enough, as well-intended individuals often fail to act on their intentions (14). Therefore, it is also crucial to facilitate action by helping people overcome barriers to follow-through, such as forgetfulness (15), hassle costs (16), and procrastination (17,18). Indeed, low-cost nudges such as the use of defaults (19), planning prompts (20), or reminders (21) have proven effective in encouraging influenza vaccinations. A recent review of the literature proposes that this latter approach—focusing on mobilizing people who already intend to get vaccinated—may be more effective than focusing on changing intentions (11).

In this paper, we report data from two large-scale randomized controlled trials (RCTs) that tackle the COVID-19 vaccine uptake challenge from both approaches. We sought to shift vaccination intentions and harness psychological insights to motivate action. To investigate the separate and joint effects of these approaches, we delivered text-based interventions in two sequential RCTs within a large integrated academic health system. Testing the effects of behavioral nudges on actual COVID-19 vaccine appointment scheduling and uptake behavior extends recent work that explores this question in hypothetical scenarios (22–24). To inform national vaccination outreach efforts as soon as possible, we report early results from these ongoing RCTs.

## Methods and Results

We conducted two RCTs with UCLA Health (clinicaltrials.gov numbers: NCT04800965 and NCT04801524). This research was approved by the UCLA Institutional Review Board, which granted a waiver of informed consent. The Amazon Mechanical Turk and Prolific Academic studies were reviewed and approved by the IRB at Carnegie Mellon University, the IRB#: IRBSTUDY2015_00000482. The sample for this early report consists of patients eligible to get vaccinated at UCLA Health in January-February 2021, including patients older than 65, patients with any transplant, and high-risk patients with qualifying pre-existing conditions. Additional information on vaccine eligibility and allocation is available in Supplementary Information (SI). Once becoming eligible, patients received an initial invitation. The two RCTs occurred afterwards following the timeline described below (also see Figure 1). All eligibility criteria and analyses in this report follow the pre-registration.

**Figure 1.**
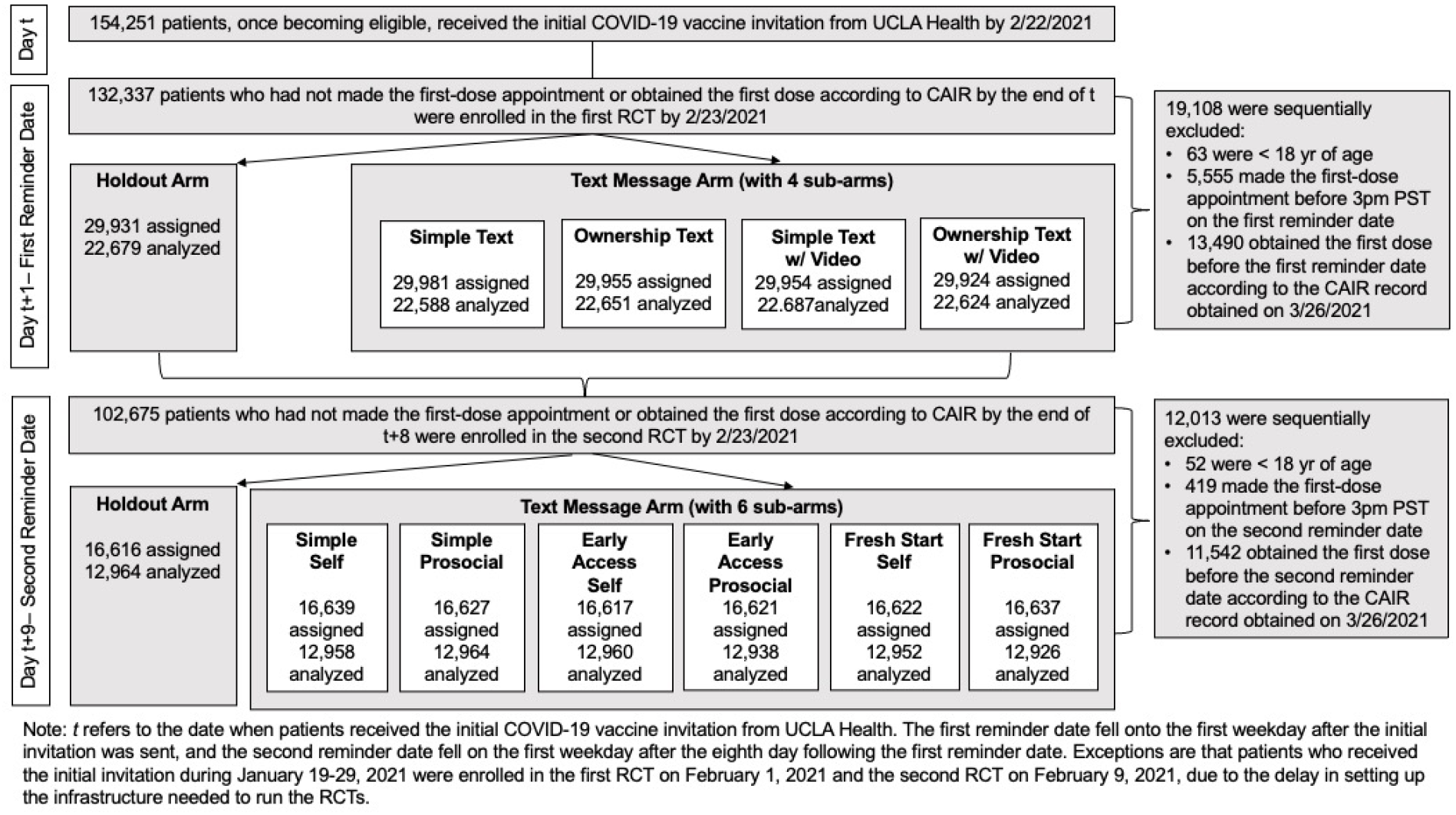
Assessment for eligibility, randomization, and timeline of two sequential RCTs

### First RCT

On the first weekday following the initial invitation (hereafter, *the first reminder date*), patients were enrolled in the first RCT if they (1) had a mobile phone number or SMS capable phone number at UCLA Health and (2) had not scheduled a (first dose) COVID-19 vaccination appointment at UCLA Health or obtained the first dose anywhere as recorded by the California Immunization Registry (CAIR) as of the day before the first reminder date. We randomly assigned patients at a 4:1 ratio to either the *Text Message* Arm, where patients received a text message at 3pm PST on the first reminder date encouraging them to schedule a vaccination appointment, or a *Holdout* arm with no text message. We designed the text message reminder to facilitate action in two ways. First, the reminder made vaccine availability top of mind, in order to overcome forgetfulness and prompt people to adopt the target behavior (15). Second, it sought to minimize potential sources of friction (e.g., inconvenience) by including a link to the appointment scheduling website, allowing patients to easily act right away (7). The earliest first reminder date was February 1, 2021. Patients whose preferred language was Spanish received the text message in Spanish.

To further test how to leverage psychological insights to encourage vaccine uptake, we nested a 2×2 factorial design within the *Text Message* Arm. The resulting 4 sub-arms (Table 1 Panel A) varied (1) whether the message attempted to induce feelings of psychological ownership for the vaccine (ownership intervention) and (2) whether the text message encouraged patients to watch a 2-minute video about COVID-19 designed to correct misconceptions about COVID-19 vaccines and thus reduce vaccine hesitancy (video intervention).

**Table 1.** Text Messages used in the Two Sequential RCTs

| Panel A: Text Messages Tested in the First RCT |  |
| --- | --- |
| Sub-arm Name | Text |
| Simple Text | UCLA Health: [Patient's first name], you can get the COVID-19 vaccine at UCLA Health. Make a vaccination appointment here: uclahealth.org/schedule |
| Ownership Text | UCLA Health: [Patient's first name], a COVID-19 vaccine has just been made available to you at UCLA Health. Claim your dose today by making a vaccination appointment here: uclahealth.org/schedule |
| Simple Text w/ Video | UCLA Health: [Patient's first name], you can get the COVID-19 vaccine at UCLA Health. Please watch this important 2 min video: [Link to Video]. Make a vaccination appointment here: uclahealth.org/schedule |
| Ownership Text w/ Video | UCLA Health: [Patient's first name], a COVID-19 vaccine has just been made available to you at UCLA Health. Please take 2 simple steps: 1. Watch this important 2 min video: [Link to Video]. 2. Claim your dose today by making a vaccination appointment here: uclahealth.org/schedule |
| Panel B: Text Messages Tested in the Second RCT |  |
| Sub-arm Name | Text |
| Simple Self | UCLA Health: [Patient's first name], to protect yourself, make your COVID-19 vaccine appointment here today: uclahealth.org/schedule |
| Simple Prosocial | UCLA Health: [Patient's first name], to protect your family, friends, and community, make your COVID-19 vaccine appointment here today: uclahealth.org/schedule |
| Early Access Self | UCLA Health: [Patient's first name], you are one of few Americans who have early access to the COVID-19 vaccine based on national guidelines. Take this opportunity to protect yourself. Make your COVID-19 vaccine appointment here today: uclahealth.org/schedule |
| Early Access Prosocial | UCLA Health: [Patient's first name], you are one of few Americans who have early access to the COVID-19 vaccine based on national guidelines. Take this opportunity to protect your family, friends, and community who may not have this access yet. Make your COVID-19 vaccine appointment here today: uclahealth.org/schedule |
| Fresh Start Self | UCLA Health: [Patient's first name], the past year has been tough. Now, the COVID-19 vaccine can offer the promise of a fresh start. Take this opportunity to protect yourself and chart a new path forward. Make your vaccine appointment here today: uclahealth.org/schedule |
| Fresh Start Prosocial | UCLA Health: [Patient's first name], the past year has been tough for many. Now, the COVID-19 vaccine can offer the promise of a fresh start. Take this opportunity to protect your family, friends, and community and help our nation chart a new path forward. Make your vaccine appointment here today: uclahealth.org/schedule |

The ownership intervention (used in the *Ownership Text* and *Ownership Text with Video* sub-arms) leveraged insights about the endowment effect and psychological ownership to further amplify people’s desire to act (25–27). The message indicated the vaccine had “*just been made available for you*” and encouraged patients to “*claim your dose*.” We hypothesized (and confirmed via studies reported in SI) that such language would make patients feel endowed with the vaccine. Recent experiments suggest that similar language like “*the flu vaccine is reserved /waiting for you*” can increase influenza vaccination uptake (21); psychological ownership could be one of the mechanisms at play.

The video intervention (used in the *Simple Text with Video* and *Ownership Text with Video* sub-arms) aimed to test whether adding information that attempted to shift vaccination intentions would further enhance the effectiveness of text messages designed to motivate action. We based the video on a literature review of vaccine hesitancy (11,28–29) and our survey of California residents in January 2021 (N=515; see SI). These efforts helped us identify common misconceptions about COVID-19 and authorized vaccines as well as discrepancies in beliefs between individuals who intended to get the vaccine and those feeling uncertain. The resulting video (https://player.vimeo.com/video/512342333) first highlighted the pandemic as a challenge, providing statistics on infections and the ease of transmission. Then, it proposed the vaccine as an easy and safe solution, providing information about the vaccine’s effectiveness and urging patients to schedule their appointment. Patients whose preferred language was Spanish received the video in Spanish. A similar video intervention significantly increased influenza vaccinations in recent work (30).

Our early analysis includes 113,229 patients who were enrolled in the first RCT no later than February 23, 2021, excluding those who were below 18 years old, had scheduled the first-dose appointment at UCLA Health before 3pm PST on the first reminder date, or had obtained the first dose before their first reminder date according to their latest CAIR record (obtained on March 26, 2021). These patients were on average 72.8 years old (s.d. = 10.1), 43.0% of them were male, 55.5% were non-Hispanic white, 4.4% preferred Spanish, and 51.2% received the flu shot in the 2019-2020 or 2020-2021 flu season. Study arms were well-balanced on demographics and prior flu vaccination status, except for a slight imbalance on the proportion of non-Hispanic white (see SI).

Our primary outcome measure was whether patients scheduled the first-dose appointment at UCLA Health within six days (from 3pm PST on the first reminder date to 11:59pm PST on the fifth ensuing day). The results are robust if we extend the observation period to March 26, 2021 (date of data extraction; see SI). Our secondary outcome measure was whether patients got the first dose at UCLA Health within four weeks of the first reminder date. This time window was appropriate because UCLA Health generally only allowed scheduling appointments less than four weeks in advance (see SI).

Figure 2 presents average appointment and vaccination rates by condition. All reported analyses use ordinary least squares (OLS) regressions and control for patient race/ethnicity, gender, preferred language, social vulnerability score, COVID-19 risk score, and fixed effects of initial invitation dates. The results are robust to removing control variables or using logistic regressions (see SI for exact specifications, all regression results, and robustness checks).

**Figure 2.**
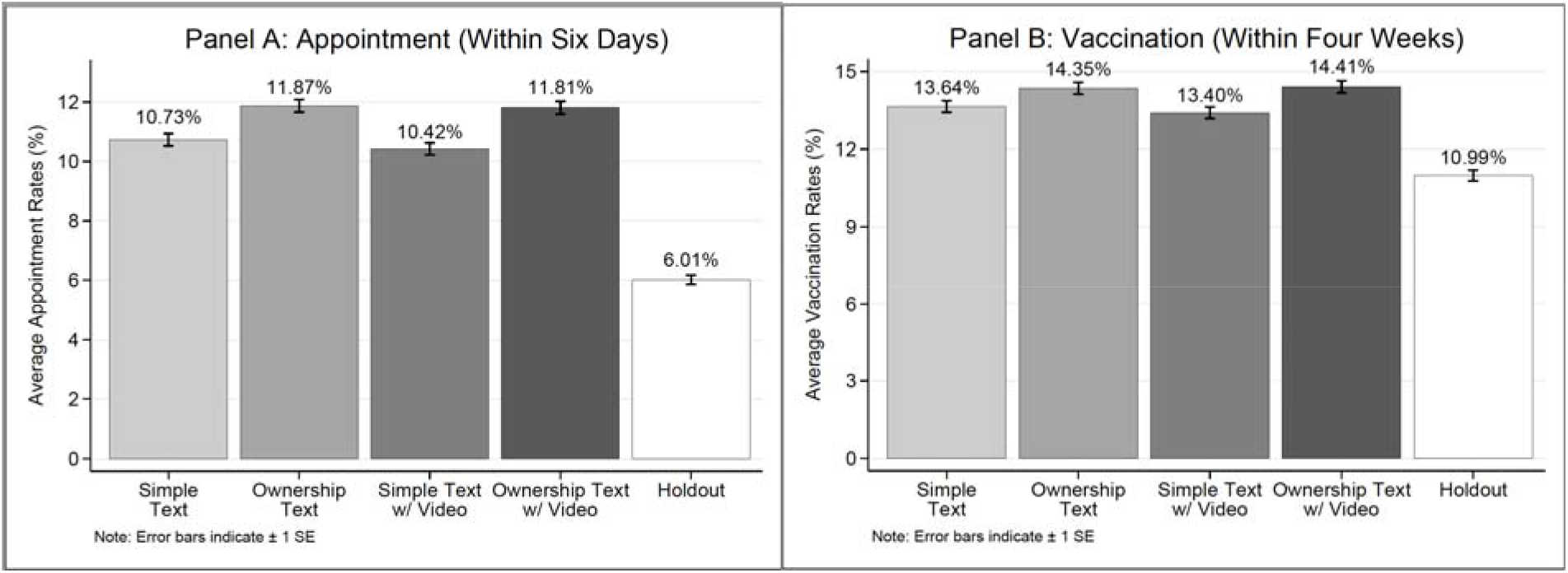
Appointment and vaccination rates by condition (first RCT)

In the *Holdout* arm, 6.01% of patients made the first-dose appointment within six days of the first reminder date and 10.99% received the first dose at UCLA Health within four weeks. Receiving a text message boosted appointment rates within six days by 5.14 percentage points (p<0.001) and vaccination rates within four weeks by 2.89 percentage points (p<0.001), amounting to a relative increase of 85.52% and 26.30%, respectively. Receiving a message also accelerated vaccinations: participants in the *Text Message* arm took only 17 days to reach the vaccination rates achieved by the *Holdout* arm in 27 days (see SI). We subsequently confirmed that all message types (Simple Text, texts highlighting ownership, texts with the video) outperformed the *Holdout* arm (p’s<0.001).

The average effect of a text message and the simple effect of each message type were significantly larger among patients who received the flu shot in either of the two recent seasons than those who did not (*p’*s<0.001). The former subpopulation tends to have stronger COVID-19 vaccination intentions (28), which is consistent with our observation that these patients’ vaccination rates in the *Holdout* arm more than doubled vaccination rates of patients who did not get the flu shot in either season. This finding suggests that our text messages are better at facilitating action among individuals with more favorable attitudes towards vaccines.

Within the *Text Message* arm, adding (vs. not including) the ownership language increased both appointment and vaccination rates at UCLA Health by 1.23 and 0.82 percentage points, respectively (*p’*s<0.001). By contrast, inviting patients to watch an educational video did not improve either outcome variable, relative to messages without a video (*p’*s>0.52). This may be due to the low video view rate (below 21%; see SI), possibly driven by patients’ busyness or by strategic information avoidance (31). Indeed, in a recent field study, a similar video-based information intervention (vs. a placebo video) significantly increased influenza vaccination rates among people who voluntarily watched it (30).

Since our sample consists of predominantly elderly and white patients, we confirmed (see Figure 3) that the effects of text message and ownership language largely hold for non-white patients (n=34,384) and patients below 65 years old (n=11,343). Notably, the average effects of receiving the text message on both appointments and vaccinations were comparable across White (n=62,856), Hispanic (n=11,758), African American (n=5,458), and Asian (n=9,182) patients (see SI).

**Figure 3.**
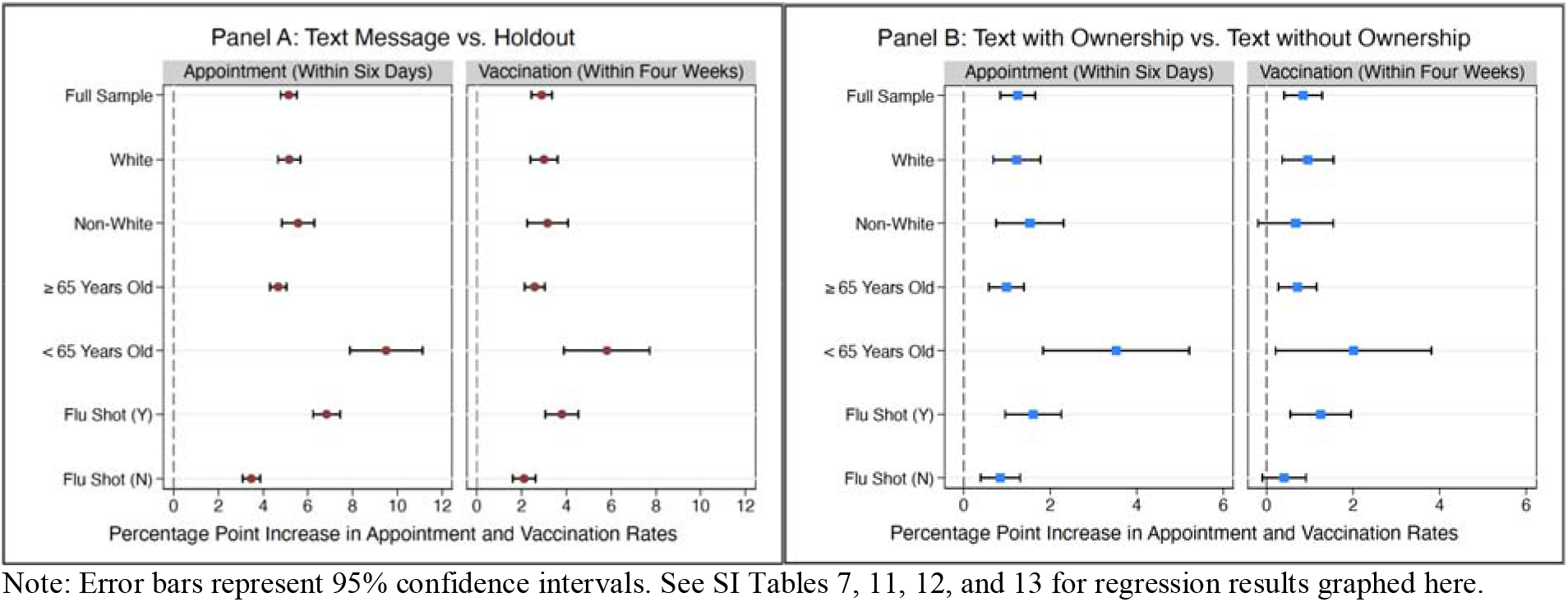
Regression-estimated increase in appointments and vaccinations induced by receiving a text message (vs. *Holdout*; Panel A) and by receiving a message with ownership language (vs. a message without ownership language; Panel B) across patient subgroups (first RCT) Note: Error bars represent 95% confidence intervals. See SI Tables 7, 11, 12, and 13 for regression results graphed here.

Altogether, early results of our first RCT suggest that messages aimed at encouraging follow-through (e.g., our simple reminder with easy access to appointment scheduling) are effective across patient subgroups and can be further heightened by inducing feelings of psychological ownership; combining such messages with a video-based information intervention targeting vaccination intentions did not further improve vaccine uptake.

### Second RCT

Even if patients received a reminder in the first RCT, those who did not schedule their appointment right away may have forgotten about it or kept procrastinating. Thus, we conducted the second RCT to study the effect of sending a second text reminder. On the first weekday after the eighth day following the initial invitation (hereafter, *the second-reminder date*), we enrolled patients who had not scheduled the first-dose appointment at UCLA Health or had not obtained the first dose anywhere according to CAIR. We randomized patients at a 6:1 ratio to either the *Text Message* arm where they received another text message reminding them of vaccine availability and providing easy access to the scheduling website, or the *Holdout* arm receiving no text message. Text messages were sent at 3pm PST on the second reminder date. Randomization was independent between the first and second RCTs (see SI). The earliest second reminder date was February 9, 2021.

To harness another set of psychological principles to motivate people to act, we randomized patients in the *Text Message* arm to receive one of six messages. These messages (Table 1 Panel B) intended to test the effects of emphasizing prosocial (vs. personal) benefits of vaccinations (32,33), highlighting exclusivity of having early access to the vaccine (34,35), and framing the act of obtaining the vaccine as an opportunity to chart a new path forward (36,37). In this early report, we only present results comparing text messages with the *Holdout* arm. We pre-registered to only compare sub-arms to each other once data collection has been completed due to uncertainty about how many people would be included in the early analysis of the second RCT.

Our early analysis (N=90,662) focuses on patients enrolled in the second RCT no later than February 23, 2021. The outcome measures and eligibility criteria were identical to those of the first RCT. Patients were on average 73.9 years old (s.d. = 9.4), 43.1% of them were male, 55.3% were non-Hispanic white, 4.4% preferred Spanish, and 48.2% received the flu shot in the 2019-2020 or 2020-2021 flu season. Study arms were well-balanced on demographics and previous flu vaccination status (see SI).

Getting a second reminder increased patients’ likelihood of scheduling the first-dose appointment within six days by 1.26 percentage points (a 52.28% increase; p<0.001) and getting the first dose at UCLA Health within four weeks by 0.68 percentage points (a 15.89% increase; p<0.001), relative to the *Holdout* arm (Figure 4). All message types (those highlighting personal benefits, prosocial benefits, early access, and opportunity for a fresh start) boosted appointments rates (*p’*s<.001) and vaccinations (*p’*s<0.03).

**Figure 4.**
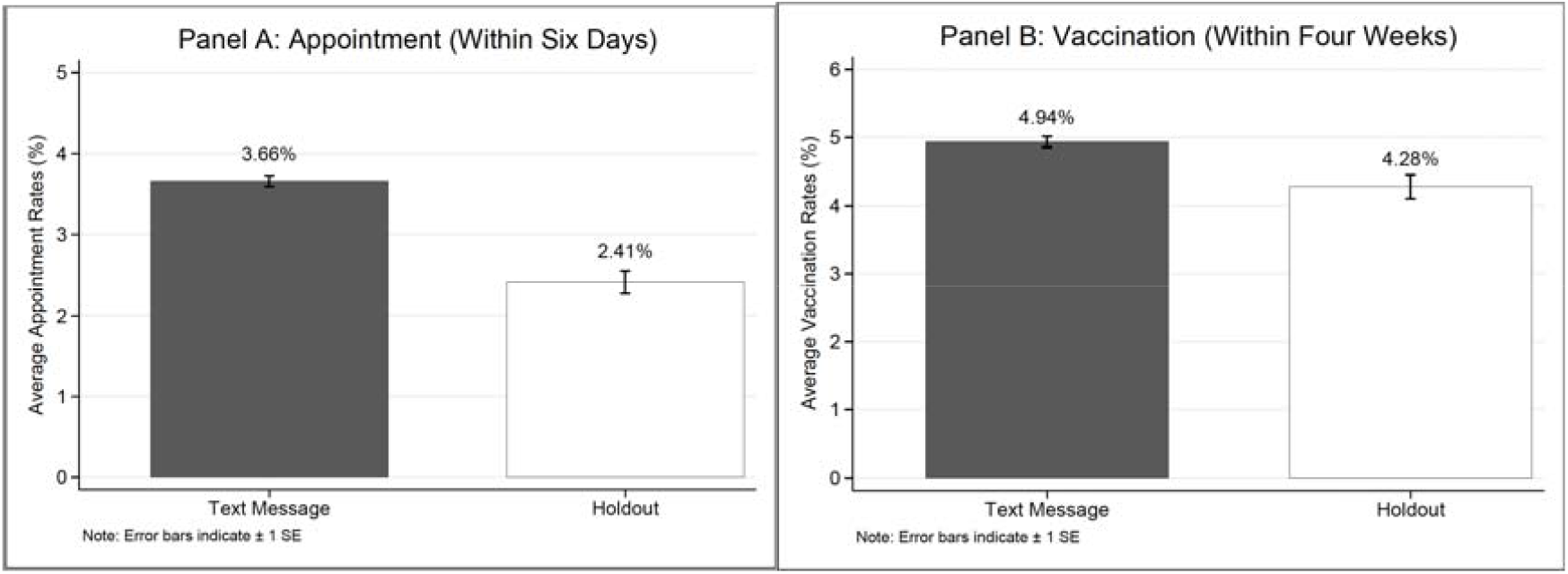
Appointment and vaccination rates in the *Text Message* vs. *Holdout* arm (Second RCT)

### Appointment Completion Rates and Second Dose

Among patients who scheduled the first-dose appointment at UCLA Health within six days of the first reminder date in the first RCT, only 12% cancelled their appointment without rescheduling a new one by March 26, 2021. Among the subset of patients who had an active first-dose appointment that was supposed to take place before March 26, 2021, only 0.1% did not attend it. Further, among those who were in the early analysis sample of either RCT and received the first dose of Moderna or Pfizer at UCLA Health by the time of our data extraction, 92.61% had already scheduled or obtained their second dose. This was facilitated by measures taken to ensure that patients would schedule their second dose immediately after completing the first dose. Overall, the low cancellation rate, the extremely low no-show rate, and the high second-dose scheduling rate suggest that the biggest barrier to tackle for the vaccine uptake challenge is getting patients to schedule an appointment for their first dose.

### Do Hypothetical Responses Reflect Actual Behaviors?

To explore how a different sample responded to the video and ownership interventions tested in the first RCT, we ran two pre-registered experiments on Amazon’s Mechanical Turk and Prolific Academic (https://aspredicted.org/blind.php?x=u2ng5c and https://aspredicted.org/blind.php?x=ae3ci5). We ran these experiments concurrently to the first RCT, in February 2021 (N=2,003; see SI for design and results). We randomized participants to receive one of the interventions from the first RCT. Contrary to the first RCT but consistent with other online studies (22,24), our online experiments found that the video intervention elevated people’s reported likelihood of scheduling an appointment (*p’*s_≤_0.01). Instead, adding ownership language did not increase (and directionally reduced) people’s stated appointment scheduling likelihood (p=0.13). This discrepancy is unlikely to be driven by differences in political attitudes between field and online samples (38), since we recruited a balanced number of Republicans and Democrats online and the aforementioned (null) effects generally held for both subpopulations. One potential explanation for the difference in the effect of video is that while we could require all online participants to watch the video, patients in the first RCT could opt in to watch it and chose to do so at a low rate (below 21%). Another possibility is that COVID-19 vaccine intentions were simply hard to change outside a controlled online experiment, especially as people likely had been receiving mass (accurate or inaccurate) information about the COVID-19 vaccine. As for why the ownership language did not motivate online participants, it may be because, in hypothetical settings, individuals failed to anticipate that the ownership manipulation would make them feel motivated to take action. Nevertheless, these results suggest that hypothetical responses to behavioral nudges should be taken with caution as they may not reflect change in actual vaccination behaviors.

## Discussion

As the United States approaches universal access to the COVID-19 vaccine, accelerating vaccinations is of the utmost importance. Our research highlights that behavioral science insights can substantially increase and speed up vaccinations at close to zero marginal cost.

Vaccination strategies should focus on helping people overcome barriers to scheduling the first dose. In our sample, most patients who scheduled their first-dose appointment 1) showed up and 2) made an appointment to receive their second dose. Our RCTs show that text-based nudges designed to make vaccination salient and reduce frictions in scheduling can effectively encourage vaccinations. Even after patients had received notifications of their vaccination eligibility, our text reminders boosted appointments and vaccination rates by 86% and 26%, respectively, and accelerated vaccinations. Further, for patients who failed to act after receiving a reminder, following up with a second reminder was effective, increasing vaccinations by about 16%.

Text reminders leveraging the endowment effect were the most effective. Reminders that induce feelings of psychological ownership for the vaccine outperformed reminders without such language. However, a video intervention attempting to correct misconceptions about the COVID-19 vaccine did not yield additional benefits. These results suggest that persuading those who are hesitant will likely require a different strategy from the video-based approach deployed in our trial.

Our text-based nudges yielded positive effects across different demographic groups. Our text reminders resulted in similar benefits for both white and non-white participants, in contrast with other recent studies (21). Also, our reminders were beneficial for older patients (≥ 65 years old), and even more so for younger ones (< 65 years old). This finding is especially important given vaccines are becoming available to the younger population, who are generally more accustomed to text messaging. These results demonstrate text-based nudges can be beneficial for a variety of different demographics.

Interestingly, people in our large-scale online experiments did not anticipate feeling motivated by the ownership intervention but updated COVID-19-related beliefs and reported greater likelihood of scheduling a vaccination appointment after exposure to the video intervention. This discrepancy pinpoints the importance of pilot-testing behavioral interventions in the real world before scaling them up.

Altogether, text-based nudges that harness insights from behavioral science are a promising avenue for tackling the challenge of COVID-19 vaccine uptake. As COVID-19 vaccines become more widely available, we hope that our research will inform nationwide vaccination campaigns and, ultimately, accelerate our society’s return to normalcy.

## Data Availability

The data analyzed in this paper about randomized controlled trials was provided by UCLA Health and contains protected health information. To protect patient privacy, we cannot publicly post individual-level data on vaccinations. Data about our online experiments and survey are available.

https://osf.io/qn8hr/?view_only=cf7b2bc590054aee8c4a2bae99ef20c5

## Data availability

The data analyzed in this paper about randomized controlled trials was provided by UCLA Health and contains protected health information. To protect patient privacy, we cannot publicly post individual-level data on vaccinations. Data about our online experiments and survey are available at: https://osf.io/qn8hr/?view_only=cf7b2bc590054aee8c4a2bae99ef20c5.

## Code availability

The code to replicate the analyses and figures in the manuscript and Supplementary Information is available at: https://osf.io/qn8hr/?view_only=cf7b2bc590054aee8c4a2bae99ef20c5.

## Author contributions

Conceptualization: D.M.C., H.D., and S.S.; Data Curation: H.D., H.M., S.P., and S.S.; Formal Analysis: H.D., S.S., and S.V.; Funding Acquisition: D.M.C. and M.A.H.; Investigation: D.M.C., H.D., S.S., M.A.H., N.R., and L.R.; Methodology: D.M.C., H.D., S.S., and M.A.H.; Project Administration: D.M.C., H.D., and S.S.; Supervision D.M.C., H.D., and S.S.; Validation: H.D., S.S., S.V.; Visualization: H.D. and S.S.; Writing - Original Draft: H.D. and S.S.; Writing - Review & Editing: D.M.C., H.D., S.S., M.A.H., H.M., S.P., L.R., N.R., and S.V.

## Additional Information

Supplementary Information is available for this paper.

## Acknowledgments

Funding support for this research was provided by UCLA Health, Anderson School of Management, and Carnegie Mellon University. We thank UCLA Health, particularly Linda Ho, Jomarr Ileto, Anne Machalinski, and Tuong Tieu, for making this research possible. We thank Saurabh Bhargava, Gretchen Chapman, Noah Goldstein, Ian Hurst, Christine Nyungen, and Katy Milkman for feedback on the paper. We thank Jose Cervantez, Mandy Lanyon, and Stephanie Permut for assistance with this research.

