## Supplementary Information for "Behavioral nudges increase COVID-19 vaccinations: Two randomized controlled trials"

April 2021

### Contents

**1 Randomized Controlled Trials** **3**
1.1 Method 3
1.1.1 Participants, Eligibility, Enrollment 3
1.1.2 Interventions 5
1.2 Outcome Measures 8
1.3 Analyses and Results about the First RCT 9
1.3.1 The Average Effect of Receiving a Text Reminder 10
1.3.2 Comparing Different Types of Text Reminders with Holdout 11
1.3.3 The Effects of Ownership Language and the Educational Video 14
1.3.4 The Heterogeneous Treatment Effect by Flu Vaccination Status 16
1.3.5 Generalizability Across Racial/Ethnic and Age Groups 22
1.4 Results about the Second RCT 26
1.4.1 The Average Effect of Receiving the Second Text Reminder 26
1.4.2 Comparing Different Types of Text Reminders with Holdout 26
1.5 Additional Information and Robustness Checks 31

**2 Online Experiments Assessing Intentions and Mechanisms** **37**
2.1 Method 37
2.1.1 Participants 37
2.1.2 Procedure 38
2.1.3 Measures 39
2.2 Results 41
2.2.1 Scheduling Likelihood 41
2.2.2 Message Persuasiveness 41
2.2.3 Mechanisms for the Ownership Text 42
2.2.4 Mechanisms for the Video 43
2.2.5 Heterogeneous Treatment Effects 46

**3 Online Survey Comparing Beliefs and Perceptions Across People with Different Vaccination Intentions** **60**
3.1 Method 60
3.1.1 Participants 60
3.1.2 Procedure 60
3.2 Results 63

### 1 Randomized Controlled Trials

#### 1.1 Method

##### 1.1.1 Participants, Eligibility, Enrollment

Our randomized control trials (RCT) are part of the vaccination outreach effort at the University of California, Los Angeles (UCLA) Health, a large academic healthcare system in California (ClinicalTrials.gov numbers: NCT04800965 and NCT04801524). The RCTs were reviewed and approved by the institutional review board (IRB) at University of California, Los Angeles, which determined that a waiver of informed consent was appropriate. Starting from January 19, 2021, UCLA Health invited patients who were eligible for the COVID-19 vaccine at the time to get vaccinated. UCLA Health sent out invitations to patients in batches via their preferred contact method.[1]

On the first weekday[2] following the initial invitation (hereafter "the first reminder date"), patients could receive a text reminder from UCLA Health if they satisfied the following criteria: (1) they had a mobile phone number or SMS-capable phone number in UCLA Health's database, (2) they had not already scheduled the first-dose vaccination appointment at UCLA Health by the end of the previous day, and (3) they had not obtained the first dose inside or outside UCLA Health according to the latest records in California Immunization Registry (CAIR) by the end of the previous day. Patients who satisfied these criteria were automatically enrolled in our first RCT and were randomly assigned into one of the arms using the simple randomization method (see Section 1.1.2). The earliest first reminder date was February 1, 2021. Text reminders were always sent at 3pm PST.

On the first weekday after the eighth day following the initial invitation (hereafter, "the second reminder date"),[3] patients could receive another text reminder at 3pm PST from UCLA Health if they had a mobile phone number or SMS-capable phone number in UCLA Health's database as well as if they had not scheduled the first-dose vaccination appointment at UCLA Health or obtained the first dose as recorded in CAIR by the end of the day before. Patients who satisfied these criteria were automatically enrolled in our second RCT and were randomly assigned into one of the arms using the simple randomization method (see Section 1.1.2). The earliest second reminder date was February 9, 2021.

Following our pre-registration, we conducted an early analysis of the first RCT using data about patients whose first reminder date was no later than February 23, 2021; similarly, we conducted an early analysis of the second RCT using data about patients whose second reminder date was no later than February 23, 2021. Those patients were eligible for the COVID-19 vaccine because they were at or above 65 years old, had

[1]UCLA Health is implementing national Advisory Committee on Immunization Practices (ACIP), state, and county guidelines to determine patient phasing. The phasing recommendation from the State of California is accessible at `https://covid19.ca.gov/vaccines/#California's-vaccination-plan`. As UCLA Health has large volumes in each phase, UCLA Health has developed a risk model that incorporates clinical risk and social risk to sub-prioritize within each phase due to limited vaccine supply. Batches of vaccine invitations were decided daily based on a) available doses, b) available appointment slots, and c) expected acceptance rate (which is different by phase).

[2]Since the infrastructure needed to run the RCTs was not ready until February 2021, patients who received the initial invitation during January 19-29, 2021 were enrolled in the first RCT on February 1, 2021. All other batches of patients were enrolled in the first RCT on the first weekday after the initial invitation was sent out.

[3]Since the infrastructure needed to run the RCTs was not ready until February 2021, patients who received the initial invitation during January 19-29, 2021 were enrolled in the second RCT on February 9, 2021. All other batches of patients were enrolled in the second RCT on the first weekday after the eighth day following the initial invitation.

any transplant, or had high-risk conditions stipulated by the Centers for Disease Control and Prevention. Per our pre-registration, we excluded the small number of patients who were were below 18 years old. We further excluded patients who were enrolled in the first (second) RCT but either scheduled a vaccination appointment at UCLA Health by 3pm PST on their corresponding first (second) reminder date or obtained a COVID-19 vaccine before their corresponding first (second) reminder date according to the CAIR record as of March 26, 2021.[4] See Figure 1 for a diagram illustrating eligibility, timeline, and randomization for both RCTs.

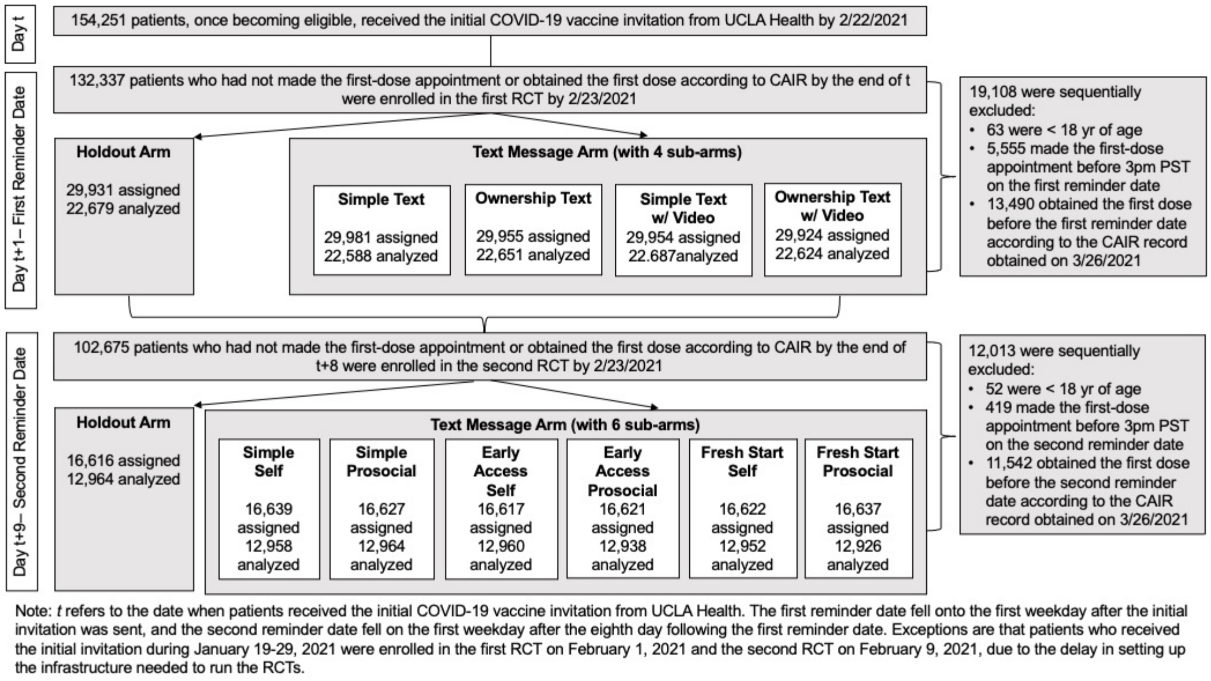


Figure 1: The Flow Chart of the First and Second RCTs

In the end, our early analysis about the first RCT included 113,229 patients. They were an average of 72.81 years old (s.d. = 10.11), 43.02% were male, 55.51% were non-Hispanic white, 4.40% preferred Spanish, and 51.17% received the flu shot in either the past (2019-2020) or current (2020-2021) flu season. Our early analysis about the second RCT included 90,662 patients. They were an average of 73.85 years old (s.d. = 9.42), 43.08% were male, 55.31% were non-Hispanic white, 4.44% preferred Spanish, and 48.21% received the flu shot in either the past (2019-2020) or current (2020-2021) flu season. The arms of the first RCT were well-balanced on age, gender, language preference, and flu vaccination status (p-values from all F-tests > 0.05, see Table 1). The p-value for F-test on whether a patient was non-Hispanic white was below 0.05 because there were slightly more non-Hispanic white patients in the *Holdout arm* (56.01%) than in the *Simple Text* sub-arm (55.06%) and the *Simple Text with Video* sub-arm (54.84%). As explained later, our

[4]We tried our best to ensure that patients were only enrolled in an RCT if they did not have an appointment at UCLA Health and they had not already obtained the vaccine somewhere based on the CAIR record by the end of the day before their first (second) reminder date. However, the records at UCLA Health and CAIR were not always up to date, so we had to exclude patients based on their latest records on March 26, 2021 (when we extracted data for early analyses). Also, 6,056 patients scheduled an appointment at UCLA Health between 12:00am and 3pm PST on the first (second) reminder date, and the primary outcome variable of these patients could not be affected by our text message sent at 3pm PST.

regressions control for patient race and ethnicity. The arms of the second RCT were well-balanced on age, gender, race, language preference, and flu vaccination status (p-values from all F-tests > 0.05, see Table 2).

Table 1: Randomization Check of the First RCT

| Variable | Simple Text Mean/SD | Ownership Text Mean/SD | Simple Text with Video Mean/SD | Ownership Text with Video Mean/SD | Holdout Mean/SD | F-test for Joint Significance (p-value) |
|---|---|---|---|---|---|---|
| Age (years) | 72.697 | 72.862 | 72.773 | 72.764 | 72.951 | 0.07 |
| | (10.105) | (10.100) | (10.064) | (10.213) | (10.071) | |
| Male | 0.430 | 0.432 | 0.433 | 0.425 | 0.432 | 0.44 |
| | (0.495) | (0.495) | (0.495) | (0.494) | (0.495) | |
| Non-Hispanic White | 0.551 | 0.558 | 0.548 | 0.558 | 0.560 | 0.04 |
| | (0.497) | (0.497) | (0.498) | (0.497) | (0.496) | |
| Spanish as the | 0.044 | 0.045 | 0.045 | 0.043 | 0.043 | 0.67 |
| Preferred Language | (0.204) | (0.208) | (0.207) | (0.202) | (0.204) | |
| Received Flu Shot | 0.509 | 0.510 | 0.511 | 0.514 | 0.514 | 0.81 |
| in Two Recent Seasons | (0.500) | (0.500) | (0.500) | (0.500) | (0.500) | |

Table 2: Randomization Check of the Second RCT

| Variable | Simple Self Mean/SD | Simple Prosocial Mean/SD | Early Access Self Mean/SD | Early Access Prosocial Mean/SD | Fresh Start Self Mean/SD | Fresh Start Prosocial Mean/SD | Holdout Mean/SD | F-test for Joint Sig. (p-value) |
|---|---|---|---|---|---|---|---|---|
| Age (years) | 73.855 | 73.762 | 73.843 | 73.861 | 73.815 | 74.022 | 73.797 | 0.42 |
| | (9.524) | (9.506) | (9.338) | (9.503) | (9.364) | (9.376) | (9.293) | |
| Male | 0.425 | 0.429 | 0.434 | 0.438 | 0.429 | 0.428 | 0.433 | 0.36 |
| | (0.494) | (0.495) | (0.496) | (0.496) | (0.495) | (0.495) | (0.496) | |
| Non-Hispanic White | 0.554 | 0.551 | 0.555 | 0.550 | 0.556 | 0.555 | 0.550 | 0.93 |
| | (0.497) | (0.497) | (0.497) | (0.497) | (0.497) | (0.497) | (0.498) | |
| Spanish as the | 0.046 | 0.044 | 0.043 | 0.044 | 0.043 | 0.045 | 0.046 | 0.79 |
| Preferred Language | (0.209) | (0.206) | (0.202) | (0.205) | (0.202) | (0.207) | (0.209) | |
| Received Flu Shot | 0.479 | 0.485 | 0.483 | 0.481 | 0.482 | 0.480 | 0.485 | 0.92 |
| in Two Recent Seasons | (0.500) | (0.500) | (0.500) | (0.500) | (0.500) | (0.500) | (0.500) | |

##### 1.1.2 Interventions

**The first RCT**. We randomly assigned patients at a 4:1 ratio to either the *Text Message* arm or the *Holdout* arm. Patients in the *Text Message* arm received a text encouraging them to schedule a vaccination appointment at UCLA Health, which was sent at 3pm PST on their first reminder date. The text message sought to remind patients of their vaccination eligibility, made vaccine availability top of mind, and make it easier for patients to schedule an appointment immediately by including a link (`https://uclahealth.org/schedule`) to UCLA Health's appointment scheduling website. Patients in the *Holdout* arm did not receive such a text message.

We nested a 2x2 factorial design within the *Text Message* arm. The first factor was whether or not the text message leveraged endowment effect (Kahneman et al., 1990) and psychological ownership (Pierce et al., 2001; Shu and Peck, 2011) to prompt patients to take action. We created text content to make patients feel that a COVID-19 vaccine had just been made available to them and they should claim their dose (rather than missing out on it). Recent experiments about influenza vaccination suggest that similar language like "the flu vaccine is reserved /waiting for you" can increase uptake (Milkman et al., 2021); psychological ownership could be one of the mechanisms at play.

The second factor was whether or not the text message contained the link to a video (`https://player.vimeo.com/video/512342333`) that we designed to shift patients' beliefs and perceptions about COVID-19 and the COVID-19 vaccine, thus boosting their intentions to get the vaccine. We based the content of the video on (1) a literature review about vaccine hesitancy (Brewer et al., 2017; Lazarus et al., 2020; Ruiz and Bell, 2021) and (2) our survey of 515 California residents in January 2021 designed to identify discrepancies in beliefs and perceptions between people who intended to get the COVID-19 vaccine and those feeling uncertain (see Section 3 in Supplementary Information). The resulting two-minute video first highlighted the pandemic as a challenge, providing statistics about the prevalence and contagiousness of COVID-19; then it proposed the vaccine as an easy and safe solution, discussing the effectiveness and safety of the authorized COVID-19 vaccines; and it finally urged patients to get the vaccine (e.g., emphasizing the importance of getting vaccinated, showing healthcare workers at UCLA Health getting vaccinated as role models).

In accordance with our 2x2 factorial design, patients in the *Text Message* arm were randomly assigned with an equal probability to either the *Simple Text*, *Ownership Text*, *Simple Text with Video*, or *Ownership Text with Video* condition. In all conditions within the *Text Message* arm, patients whose preferred language was Spanish received the text message in Spanish; and these patients were sent the Spanish version of our educational video if they were in the *Simple Text with Video* condition or the *Ownership Text with Video* condition.

- In the *Simple Text* condition, the message read, *"UCLA Health: [Patient's first name], you can get the COVID-19 vaccine at UCLA Health. Make a vaccination appointment here: uclahealth.org/schedule"*.
- In the *Ownership Text* condition, the message read, *"UCLA Health: [Patient's first name], a COVID-19 vaccine has just been made available to you at UCLA Health. Claim your dose today by making a vaccination appointment here: uclahealth.org/schedule"*.
- In the *Simple Text with Video* condition, the message read, *"UCLA Health: [Patient's first name], you can get the COVID-19 vaccine at UCLA Health. Please watch this important 2 min video: [Link to Video]. Make a vaccination appointment here: uclahealth.org/schedule"*. The first link to the video directed patients to a survey where we embedded the video. A sample of the survey can be accessed here: `https://tinyurl.com/f3hfwwsh`. Patients could start and stop the video at any time.
- In the *Ownership Text with Video* condition, the message read, *"UCLA Health: [Patient's first name], a COVID-19 vaccine has just been made available to you at UCLA Health. Please take 2 simple steps: 1. Watch this important 2 min video: [Link to Video]. 2. Claim your dose today by making a vaccination appointment here: uclahealth.org/schedule"*. The first link directed patients to a survey where we embedded the video. Readers can experience it here: `https://tinyurl.com/nv9wktsf`. Different from the *Simple Text with Video* condition, the survey in the Ownership Text with Video condition

displayed a button below the video 60 seconds after patients landed in the survey. The button reiterated the first sentence and the second step in the text message, reading, *"A COVID-19 vaccine is waiting for you at UCLA Health. Claim your dose today by scheduling your appointment here."* The button redirected patients to uclahealth.org/schedule. The button was intended to help patients follow through on their intentions to get the COVID-19 vaccine, after their intentions to get the vaccine had (hopefully) been boosted by the video.

**The second RCT**. We randomly assigned patients who did not schedule a vaccination appointment nine days after the initial invitation at a 6:1 ratio to either the *Text Message* arm or the *Holdout* arm. Patients in the *Text Message* arm received a second text reminder encouraging them to obtain the COVID-19 vaccine at UCLA Health, which was sent at 3pm PST on their second reminder date. Again, all text messages included a link (`https://uclahealth.org/schedule`) to UCLA Health's COVID-19 vaccination appointment scheduling website, so as to make it easier for patients to immediately schedule an appointment. Patients in the *Holdout* arm did not receive the second text reminder.[5]

We nested a 2x3 factorial design within the *Text Message* arm. The first factor was whether the text message focused on patients' personal or prosocial benefits of getting vaccinated. The second factor had three levels and was whether the text message highlighted the early access patients had to the vaccine, whether it highlighted that the COVID-19 vaccine offered the promise of a fresh start, or neither. Thus, patients in the *Text Message* arm were randomly assigned with an equal probability to one of six conditions. Patients whose preferred language was Spanish received the text message in Spanish.

- In the *Simple Self* condition, the message read, *"UCLA Health: [Patient's first name], to protect yourself, make your COVID-19 vaccine appointment here today: uclahealth.org/schedule"*.
- In the *Simple Prosocial* condition, the message read, *"UCLA Health: [Patient's first name], to protect your family, friends, and community, make your COVID-19 vaccine appointment here today: uclahealth.org/schedule"*.
- In the *Early Access Self* condition, the message read, *"UCLA Health: [Patient's first name], you are one of few Americans who have early access to the COVID-19 vaccine based on national guidelines. Take this opportunity to protect yourself. Make your COVID-19 vaccine appointment here today: uclahealth.org/schedule"*.
- In the *Early Access Prosocial* condition, the message read, *"UCLA Health: [Patient's first name], you are one of few Americans who have early access to the COVID-19 vaccine based on national guidelines. Take this opportunity to protect your family, friends, and community who may not have this access yet. Make your COVID-19 vaccine appointment here today: uclahealth.org/schedule"*.
- In the *Fresh Start Self* condition, the message read, *"UCLA Health: [Patient's first name], the past year has been tough. Now, the COVID-19 vaccine can offer the promise of a fresh start. Take this*

[5] Note that since randomization was conducted independently for the first and seconds RCTs (as confirmed in Table 3), some patients in the *Holdout* arm of the first RCT were assigned to the *Text Message* arm in the second RCT. The text message sent to these patients on their second reminder date was the first reminder they received from UCLA Health encouraging them to get the COVID-19 vaccine. But for simplicity, we refer to the text message sent on the second reminder date as the second text reminder for everyone.

*opportunity to protect yourself and chart a new path forward. Make your vaccine appointment here today: uclahealth.org/schedule"*.

- In the *Fresh Start Prosocial* condition, the message read, *"UCLA Health: [Patient's first name], the past year has been tough for many. Now, the COVID-19 vaccine can offer the promise of a fresh start. Take this opportunity to protect your family, friends, and community and help our nation chart a new path forward. Make your vaccine appointment here today: uclahealth.org/schedule"*.

Table 3: Randomization Independence Between Two RCTs

| | Random Assignment in the First RCT | | | | |
|---|---|---|---|---|---|
| | Simple Text | Ownership Text w/ Video | Simple Text w/ Video | Ownership Text | Holdout |
| Simple Self | 2,512 | 2,525 | 2,553 | 2,583 | 2,689 |
| | 14.11% | 14.06% | 14.41% | 14.53% | 14.29% |
| Simple Prosocial | 2,576 | 2,553 | 2,487 | 2,602 | 2,671 |
| | 14.47% | 14.21% | 14.03% | 14.64% | 14.19% |
| Early Access Self | 2,516 | 2,601 | 2,524 | 2,550 | 2,690 |
| | 14.13% | 14.48% | 14.24% | 14.35% | 14.29% |
| Early Access Prosocial | 2,568 | 2,622 | 2,519 | 2,458 | 2,699 |
| | 14.42% | 14.60% | 14.21% | 13.83% | 14.34% |
| Fresh Start Self | 2,602 | 2,552 | 2,524 | 2,522 | 2,670 |
| | 14.61% | 14.21% | 14.24% | 14.19% | 14.18% |
| Fresh Start Prosocial | 2,510 | 2,621 | 2,515 | 2,501 | 2,681 |
| | 14.10% | 14.59% | 14.19% | 14.07% | 14.24% |
| Holdout | 2,523 | 2,490 | 2,600 | 2,555 | 2,723 |
| | 14.17% | 13.86% | 14.67% | 14.38% | 14.47% |

*Notes*: This table presents the number and percentage of patients who were assigned to a specific condition in the second RCT, among those who were in a given condition in the first RCT and were eligible for the second RCT. For example, among patients who were in the *Simple Text* condition of the first RCT and were eligible for the second RCT, 2,512 (or 14.11%) were assigned to the *Simple Self* condition in the second RCT. Overall, this table shows that patients in any condition of the first RCT, if enrolled in the second RCT, had an equal rate (approximately one seventh) to be assigned to one of the seven conditions in the second RCT. This confirms that group assignment in the first RCT was independent of that in the second RCT.

#### 1.2 Outcome Measures

As pre-registered, our primary outcome variable, *Appointment at UCLA*, was a binary variable indicating whether patients scheduled a vaccination appointment for the first dose of COVID-19 vaccine at UCLA Health within six days of the first (second) reminder date for the first (second) RCT. Specifically, if $s$ denotes a patient's first (second) reminder date, we tracked whether the patient scheduled any appointment between 3pm PST on day $s$ and 11:59pm PST on day $s+5$. We pre-registered this time window because UCLA Health targeted additional outreach effort to patients who still had not scheduled their appointment six days after the second reminder date. Thus, for the second RCT, patients' decisions about whether and when to schedule an appointment six days later could be affected by the additional outreach effort. To more cleanly estimate the effects of interventions in our second RCT, the time window for our primary outcome started from 3pm on the second reminder date $s$ and ended five days later (end of $t{+}s$). For consistency between RCTs, we pre-registered the same time window for the first RCT. Our results are qualitatively the same if we analyze whether patients ever scheduled an appointment after 3pm on their first (second) reminder date by the time of our data extraction (i.e., March 26, 2021). Results of this robustness check are reported in Tables 17 and 18.

Our pre-registration listed three secondary dependent variables. The first one, *Vaccinated at UCLA*, was a binary variable indicating whether patients received the first dose of COVID-19 vaccine at UCLA Health within one month following the first (or second) reminder date. We pre-registered one month because UCLA Health generally only allowed patients to make an appointment for less than four weeks ahead. Consistent with this practice, 96.21% of the first-dose appointments made by patients in our first RCT early analyses at UCLA Health occurred within four weeks from the day they were scheduled. The second one, *Vaccinated Anywhere*, was a binary variable indicating whether patients received the first dose of COVID-19 vaccine at UCLA Health or another location reporting to the CAIR within two months of the first (or second) reminder date. The third one, *Vaccination Time Lag*, was the number of days between a patient's first (or second) reminder date and the date when she received the first dose of COVID-19 vaccine anywhere as documented by the CAIR within two months of the first (or second) reminder date. We pre-registered two months (rather than one month) for the latter two outcomes to observe the effects of our interventions on vaccinations in an even longer term. Given that the earliest date patients were enrolled in our RCTs was February 1, 2021 and our data extraction occurred on March 26, 2021, we could not evaluate *Vaccinated Anywhere* and *Vaccination Time Lag* in the early analyses. We could, however, examine *Vaccinated at UCLA* with a one-month observation window, in our early analyses.[6]

##### 1.3 Analyses and Results about the First RCT

As pre-registered, our early analyses about the first RCT sought to answer the following questions.

1. What is the average effect of our behaviorally informed text reminders?
2. Do all message types—the Simple Text message, the two messages containing ownership language, and the two messages containing a video—all outperform the *Holdout* arm?
3. What is the effect of including a brief video-based information intervention in the text message?
4. What is the effect of adding ownership language to the text message?
5. Do the aforementioned effects vary based on whether or not patients received the flu vaccination in either the 2019-2020 season or the 2020-2021 season?

Due to uncertainty about how many people would be qualified for the early report, we were concerned about not having enough power to test the interaction between the video intervention and the ownership intervention. Thus, we pre-registered to only test the interaction once data collection has been completed.

In addition to reporting regression analyses, we also pre-registered that we would show the raw data for each condition without conducting hypothesis testing. Figure 2 in the main text of the paper displays the

[6]Since February 2021 has 28 days and the first (second) reminder date was always in February, 2021, we use four full weeks to represent one month in our early analyses.

average appointment rate and the average vaccination rate at UCLA Health in each of the five conditions (four sub-arms within the *Text Message* arm plus the *Holdout* arm).

##### 1.3.1 The Average Effect of Receiving a Text Reminder

To estimate the effect of receiving a text reminder that provides easy access to scheduling an appointment, we used the following ordinary least squares (OLS) regression specification:

$$Y_i = \beta_0 + \beta_1 \, Text\ Message_i + \beta_2 X_i + \epsilon_i, \tag{1}$$

where $Y_i$ represents the outcome variable of patient $i$ (including *Appointment at UCLA* and *Vaccinated at UCLA*), and $TextMessage_i$ equals one if patient $i$ was assigned to the *Text Message* arm and zero if patient $i$ was assigned to the *Holdout* arm. $X_i$ represents a host of pre-registered control variables, including (1) patient age,[7] (2) indicators for patient race/ethnicity (non-Hispanic Black, non-Hispanic Asian, Hispanic, other/mixed, unknown; with non-Hispanic White being the reference group), (3) indicators for patient gender (female, other/unknown; with male being the reference group), (4) an indicator for whether the patient preferred English or Spanish, (5) patient's Social Vulnerability Index (SVI),[8] (6) patient's COVID-19 risk score,[9] and (7) batch fixed effects (indicators for the batch in which the patient received the initial invitation). We report robust standard errors. All of our results about these RCTs remain qualitatively the same if we do not include any controls as well as if we use logistic regressions to predict our binary primary outcome variable (see Tables 19-22).

As shown in Table 4 Column 1, receiving a text reminder significantly boosted patients' likelihood of scheduling an appointment at UCLA Health within six days of the first reminder date by 5.14 percentage points (B = 0.051, SE = 0.002, $p < 0.001$). This amounts to an 85.52% increase relative to the average appointment rates of 6.01 percentage points in the *Holdout* arm. Also, as shown in Column 2, our first text reminders on average significantly boosted patients' likelihood of getting the first dose at UCLA Health within one month (precisely, four weeks) of the first reminder date by 2.89 percentage points (B = 0.029, SE = 0.002, $p < 0.001$). This corresponds to a 26.30% increase relative to the average vaccination rates of 10.99 percentage points in the *Holdout* arm.

To understand whether and how much receiving a text message accelerated vaccinations, we plotted the Kaplan-Meier curves separately for patients in the *Text Message* arm and the *Holdout* arm. The outcome

[7] For 19 patients with missing age information, we filled in the average age of patients in the first RCT sample.

[8] To support health disparity reduction, UCLA Health is incorporating the Centers for Disease Control and Prevention (CDC) Social Vulnerability Index (SVI) specific to the state of California to assign points for social risk. SVI is available at the census tract level and is applied to patients in the prioritization model. SVI incorporates the following features of each census tract: Socioeconomic status (e.g. poverty, income, education levels), household composition and disability (e.g. age distribution, single-parent households), minority status and language, and housing type and transportation (e.g. multi-unit structures, crowding, transportation access).

[9] UCLA uses the CDC high risk conditions in the Covid-19 prioritization model. UCLA Health has developed a clinical data infrastructure to identify conditions that are at increased risk or may be at increased risk for severe illness from COVID-19, based on the patients' problem list, administrative data (such as billing, encounter, admitting, discharge diagnoses, and health plan claims), lab data, and visit data. The clinical risk is then combined with social risk. The output of this COVID-19 Risk Model is the COVID-19 risk score we used as a covariate. For patients who did not have a value for social vulnerability index score or COVID-19 risk score, we filled in the average value in the first RCT sample for each corresponding variable. Since a nontrivial number of patients had missing values (e.g., 7,840 patients missed the social vulnerability index score and 27,428 patients missed COVID-19 risk score in the first RCT sample), we also controlled for whether patients had missing values.

Table 4: Effects of Receiving a Text Message on Appointments and Vaccinations at UCLA Health

| | Appointment at UCLA (Within Six Days) (1) | Vaccinated at UCLA (Within Four Weeks) (2) |
|---|---|---|
| Text Message | 0.051*** | 0.029*** |
| | (0.002) | (0.002) |
| Gender-Female | -0.005** | -0.012*** |
| | (0.002) | (0.002) |
| Gender-Other | 0.003 | -0.003 |
| | (0.032) | (0.033) |
| Age | -0.002*** | -0.003*** |
| | (0.000) | (0.000) |
| Hispanic | 0.013*** | 0.030*** |
| | (0.004) | (0.004) |
| Non-Hispanic Black | 0.042*** | 0.069*** |
| | (0.005) | (0.006) |
| Non-Hispanic Asian | 0.010** | 0.015*** |
| | (0.004) | (0.004) |
| Ethnicity-Other | -0.014*** | -0.011** |
| | (0.003) | (0.004) |
| Ethnicity-Unknown | -0.032*** | -0.041*** |
| | (0.002) | (0.002) |
| Preferred Spanish | 0.010 | 0.023*** |
| | (0.005) | (0.006) |
| Social Vulnerability Index (SVI) | -0.001*** | -0.001*** |
| | (0.000) | (0.000) |
| Missing SVI | -0.051*** | -0.077*** |
| | (0.003) | (0.003) |
| COVID-19 Risk Score | -0.000 | -0.000 |
| | (0.000) | (0.000) |
| Missing Risk Score | -0.049*** | -0.062*** |
| | (0.003) | (0.003) |
| $N$ | 113229 | 113229 |
| $R^2$ | 0.042 | 0.051 |

Robust standard errors are reported in parentheses. Controls for batch fixed effects are omitted from the regression table.
* $p < 0.05$, ** $p < 0.01$, *** $p < 0.001$

*Notes*: Text Message is a dummy variable coded as one when participants received one of the text messages in the first RCT.

variable used for this plot captures how many days had elapsed since a patient's first reminder date by the time they obtained the first dose at UCLA Health. We right-censored all patients after day 27, since our secondary outcome variable, *Vaccination at UCLA*, assessed whether each patient obtained the first dose at UCLA Health within four weeks of the corresponding first reminder date. As shown in Figure 2, the *Text Message* arm only took 17 days to reach the vaccination rates reached by the *Holdout* arm in 27 days. This suggests that our text messages not only increased vaccination rates but also accelerated vaccinations.

##### 1.3.2 Comparing Different Types of Text Reminders with Holdout

So far, we have shown that our text reminders overall effectively boosted both appointment rates and vaccination rates. Next, we break down the conditions within the *Text Message* arm and test whether the Simple Text condition, the two conditions containing ownership language, and the two conditions containing

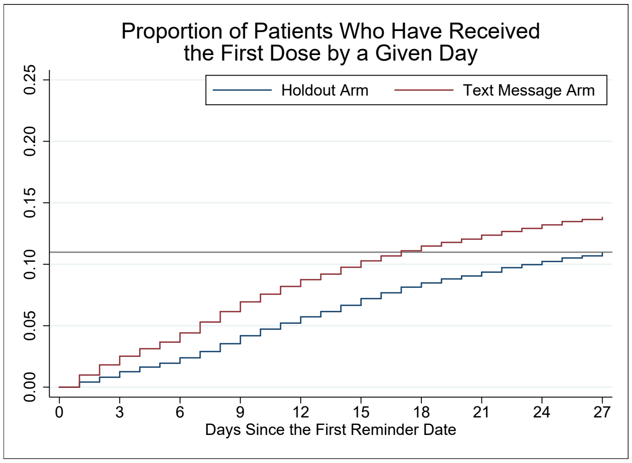


Figure 2: Kaplan-Meier Curves Reflecting the Proportion of Patients Who Have Received the First Dose by a Given Day After the First Reminder Date

a video all outperformed the *Holdout* arm. To answer this question, we conducted three regressions:

$$Y_i = \beta_0 + \beta_1 Simple\ Text_i + \beta_2 X_i + \epsilon_i, \tag{2}$$

Specification (2) only includes patients in the *Holdout* arm and the *Simple Text* condition. $SimpleText_i$ equals one if patient $i$ was assigned to the *Simple Text* condition and zero otherwise.

$$Y_i = \beta_0 + \beta_1 Ownership_i + \beta_2 X_i + \epsilon_i, \tag{3}$$

Specification (3) only includes patients in the *Holdout* arm, *Ownership Text* condition, and *Ownership Text with Video* condition. $Ownership_i$ equals one if patient $i$ was assigned to the latter two conditions that contained ownership language and zero otherwise.

$$Y_i = \beta_0 + \beta_1 Video_i + \beta_2 X_i + \epsilon_i, \tag{4}$$

Specification (4) only includes patients in the *Holdout* arm, *Simple Text with Video* condition, and *Ownership Text with Video* condition. $Video_i$ equals one if patient $i$ was assigned to the latter two conditions that contained the video and zero otherwise.

Across specifications (2)-(4), $Y_i$ represents the outcome variable of patient $i$ (including *Appointment at UCLA* and *Vaccinated at UCLA*). $X_i$ represents the same host of pre-registered control variables as mentioned in equation (1). We report robust standard errors.

Table 5 reports the results from specifications (2)-(4). First, we find that our *Simple Text* alone, which was designed to remind people of their vaccination eligibility and simplify the scheduling process, significantly boosted both appointment rates (by 4.67 percentage points or 77.70%) and vaccination rates (by 2.59 percentage points or 23.57%), relative to the *Holdout* arm. Also, the two text reminders containing ownership language significantly lifted appointment rates (by 5.75 percentage points or 95.69%) and vaccination rates (by 3.30 percentage points or 30.00%), compared to the *Holdout* arm. Finally, the two text reminders containing a video significantly outperformed the *Holdout* arm in both appointment rates (by 5.02 percentage points or 83.61%) and vaccination rates (by 2.81 percentage points or 25.61%).

Table 5: Effects of Different Types of Messages on Appointments and Vaccinations at UCLA Health

| | Appointment at UCLA (Within Six Days) | | | Vaccinated at UCLA (Within Four Weeks) | | |
|---|---|---|---|---|---|---|
| | (1) | (2) | (3) | (4) | (5) | (6) |
| Simple Text | 0.047*** | | | 0.026*** | | |
| | (0.003) | | | (0.003) | | |
| Ownership | | 0.058*** | | | 0.033*** | |
| | | (0.002) | | | (0.003) | |
| Video | | | 0.050*** | | | 0.028*** |
| | | | (0.002) | | | (0.003) |
| Gender-Female | -0.004 | -0.007** | -0.007** | -0.013*** | -0.015*** | -0.015*** |
| | (0.003) | (0.002) | (0.002) | (0.003) | (0.003) | (0.003) |
| Gender-Other | -0.006 | -0.013 | 0.034 | -0.009 | -0.020 | 0.028 |
| | (0.014) | (0.017) | (0.056) | (0.016) | (0.020) | (0.058) |
| Age | -0.001*** | -0.001*** | -0.001*** | -0.001*** | -0.002*** | -0.001*** |
| | (0.000) | (0.000) | (0.000) | (0.000) | (0.000) | (0.000) |
| Hispanic | 0.011 | 0.013** | 0.010* | 0.032*** | 0.029*** | 0.032*** |
| | (0.006) | (0.005) | (0.005) | (0.007) | (0.006) | (0.006) |
| Non-Hispanic Black | 0.030*** | 0.051*** | 0.045*** | 0.064*** | 0.076*** | 0.080*** |
| | (0.007) | (0.007) | (0.006) | (0.009) | (0.007) | (0.007) |
| Non-Hispanic Asian | 0.008 | 0.008 | 0.014** | 0.017** | 0.010 | 0.016** |
| | (0.005) | (0.005) | (0.004) | (0.006) | (0.005) | (0.005) |
| Ethnicity-Other | -0.009 | -0.015*** | -0.009* | -0.010 | -0.008 | 0.000 |
| | (0.005) | (0.004) | (0.004) | (0.006) | (0.005) | (0.005) |
| Ethnicity-Unknown | -0.032*** | -0.038*** | -0.038*** | -0.045*** | -0.049*** | -0.047*** |
| | (0.003) | (0.003) | (0.003) | (0.004) | (0.003) | (0.003) |
| Preferred Spanish | 0.024** | 0.013 | 0.010 | 0.034*** | 0.031*** | 0.023** |
| | (0.008) | (0.007) | (0.007) | (0.010) | (0.008) | (0.008) |
| Social Vulnerability | -0.000*** | -0.000*** | -0.000*** | -0.000*** | -0.000*** | -0.000*** |
| Index (SVI) | (0.000) | (0.000) | (0.000) | (0.000) | (0.000) | (0.000) |
| Missing SVI | -0.044*** | -0.056*** | -0.044*** | -0.076*** | -0.081*** | -0.073*** |
| | (0.004) | (0.003) | (0.003) | (0.004) | (0.004) | (0.004) |
| COVID-19 Risk Score | -0.000*** | -0.000*** | -0.001*** | -0.000*** | -0.000*** | -0.000*** |
| | (0.000) | (0.000) | (0.000) | (0.000) | (0.000) | (0.000) |
| Missing Risk Score | -0.060*** | -0.067*** | -0.065*** | -0.093*** | -0.096*** | -0.093*** |
| | (0.002) | (0.002) | (0.002) | (0.003) | (0.002) | (0.002) |
| $N$ | 45267 | 67954 | 67990 | 45267 | 67954 | 67990 |
| $R^2$ | 0.031 | 0.036 | 0.033 | 0.036 | 0.038 | 0.036 |
| Conditions | Simple Text | Ownership Text | Simple Text w/ Video | Simple Text | Ownership Text | Simple Text w/ Video |
| Included | | Ownership w/ Video | Ownership w/ Video | | Ownership w/ Video | Ownership w/ Video |
| | Holdout | Holdout | Holdout | Holdout | Holdout | Holdout |

Robust standard errors are reported in parentheses. Controls for batch fixed effects are omitted from the regression table.

* $p < 0.05$, ** $p < 0.01$, *** $p < 0.001$

*Notes*: Simple Text is a dummy variable coded as one when participants received the *Simple Text* message in the first RCT. Ownership is a dummy variable coded as one when participants received one of the messages containing the ownership language in the first RCT. Video is a dummy variable coded as one when participants received one of the messages containing a link to the video in the first RCT.

###### 1.3.3 The Effects of Ownership Language and the Educational Video

Next, we estimate the effects of (1) adding language designed to induce feelings of psychological ownership and (2) adding a link to the educational video designed to shift peoples' beliefs and perceptions about COVID-19 and the vaccine. We used the following OLS regression specification, which only involves people in the *Text Message* arm, given our focus on the additive effects of ownership language and video.

$$Y_i = \beta_0 + \beta_1 Ownership_i + \beta_2 Video_i + \beta_3 X_i + \epsilon_i, \tag{5}$$

where $Y_i$ represents the outcome variable of patient $i$ (including *Appointment at UCLA* and *Vaccinated at UCLA*); $Ownership_i$ and $Video_i$ are defined as in equations (3) and (4). $X_i$ represents the same host of pre-registered control variables as mentioned in equation (1). We report robust standard errors.

Table 6 show a significant, positive effect of adding the ownership language. Specifically, patients were more likely to schedule a vaccination appointment within six days by 1.23 percentage points after receiving a text reminder containing the ownership language than after receiving a text reminder without such language (B = 0.012, SE = 0.002, $p < 0.001$). Further, adding the ownership language increased patients' likelihood of getting the first dose at UCLA Health within four weeks of the first reminder date by 0.82 percentage points (B = 0.008, SE = 0.002, $p < 0.001$).

However, we found no evidence that adding a link to the educational video and inviting people to watch it boosted either appointment rates or vaccination completion rates, as shown in Table 6. We anonymously tracked the total number of clicks on the link to the video, the total number of video views, their watch time, and the total number of patients who finished watching the video each day. We focused on patients whose first reminder date was between February 1 and February 17, 2021, because no text messages for the first RCT were sent during February 18-21, 2021. We could attribute all link clicks and video views that happened by the end of February 21 to patients who had been invited to watch the video by February 17, 2021, and those patients were unlikely to go back to the text message and watch the video after February 21 since it was five or more days after their first reminder date. Using this approach, we count that the average number of link clicks per patient was 0.28 per patient, the average number of video views per patient was 0.21, and the average number of finishes per patient was 0.14. This means that at most 28% of patients clicked the link to the video, 21% watched the video and 14% finished watching it. Conditional on starting to watch the video, patients watched about 77% of the video. The low watch rates may be one of the reasons for why the video intervention did not yield a significant increase in appointment scheduling beyond the effect of the simple text message.

Table 6: Effects of Ownership Language and Video on Appointments and Vaccinations at UCLA Health

| | Appointment at UCLA (Within Six Days) | Vaccinated at UCLA (Within Four Weeks) |
|---|---|---|
| | (1) | (2) |
| Ownership | 0.012*** | 0.008*** |
| | (0.002) | (0.002) |
| Video | -0.002 | -0.001 |
| | (0.002) | (0.002) |
| Gender-Female | -0.007*** | -0.013*** |
| | (0.002) | (0.002) |
| Gender-Other | -0.003 | -0.008 |
| | (0.043) | (0.044) |
| Age | -0.002*** | -0.003*** |
| | (0.000) | (0.000) |
| Hispanic | 0.014** | 0.031*** |
| | (0.004) | (0.005) |
| Non-Hispanic Black | 0.048*** | 0.072*** |
| | (0.006) | (0.006) |
| Non-Hispanic Asian | 0.009* | 0.015*** |
| | (0.004) | (0.004) |
| Ethnicity-Other | -0.014*** | -0.012** |
| | (0.004) | (0.004) |
| Ethnicity-Unknown | -0.033*** | -0.043*** |
| | (0.003) | (0.003) |
| Preferred Spanish | 0.009 | 0.023*** |
| | (0.006) | (0.007) |
| Social Vulnerability Index (SVI) | -0.001*** | -0.001*** |
| | (0.000) | (0.000) |
| Missing SVI | -0.057*** | -0.080*** |
| | (0.003) | (0.003) |
| COVID-19 Risk Score | -0.000 | 0.000 |
| | (0.000) | (0.000) |
| Missing Risk Score | -0.053*** | -0.065*** |
| | (0.003) | (0.003) |
| $N$ | 90550 | 90550 |
| $R^2$ | 0.040 | 0.051 |
| Conditions Included | Conditions within the Text Message arm: Simple Text, Ownership Text, Simple Text w/ Video, and Ownership Text w/ Video | |

Robust standard errors are reported in parentheses. Controls for batch fixed effects are omitted from the regression table.
$^{*}$ $p < 0.05$, $^{**}$ $p < 0.01$, $^{***}$ $p < 0.001$

*Notes*: Ownership is a dummy variable coded as one when participants received one of the messages containing the ownership language in the first RCT. Video is a dummy variable coded as one when participants received one of the messages containing a link to the video in the first RCT.

###### 1.3.4 The Heterogeneous Treatment Effect by Flu Vaccination Status

As pre-registered, we explored how the effects of our interventions varied depending on whether patients got the flu shot in either the 2019-2020 season (from August 1, 2019 to March 31, 2020) or the 2020-2021 season (from August 1, 2020 to the date of our data extraction) based on patients' medical record at UCLA Health. We used flu vaccination status as a proxy for people's baseline vaccination intentions, as some survey evidence suggests that individuals' intentions to get vaccinated against COVID-19 are higher among people with previous flu vaccination behavior (Caserotti et al., 2021; Kreps et al., 2020). We separately applied regression specifications (1)-(5) to the two subgroups of patients (with or without getting the flu vaccine), and we also interacted our predictor(s) in each specification with a dummy variable (*Flu Shot*) that equals one for patients who got the flu shot in either of the two seasons and zero otherwise.

All results are presented in Tables 7-11. We highlight three observations. First of all, the average effect of receiving a text message, the values of all types of messages (relative to the *Holdout* arm), and the additional benefit of including ownership language on appointments and vaccinations hold for both patients who got the flu shot in either season and those who did not (all p-values $< 0.001$). Second, the average effect of receiving a text message is statistically significantly higher among patients who got the flu shot in either season (6.84 percentage points for appointments and 3.78 percentage points for vaccinations) than among patients who did not (3.47 percentage points for appointments and 2.11 percentage points for vaccinations; p-values for the interaction $< 0.001$). Third, the effect of each type of message (relative the *Holdout* arm) was significantly larger among patients who got the flu shot in either season than among those who did not (all p-values for the interaction $< 0.02$).

Table 7: Effects of Receiving a Text Message by Prior Flu Vaccination Status

| | Appointment at UCLA (Within Six Days) | | | Vaccinated at UCLA (Within Four Weeks) | | |
|---|---|---|---|---|---|---|
| | Flu Shot=0 | Flu Shot=1 | Both | Flu Shot=0 | Flu Shot=1 | Both |
| | (1) | (2) | (3) | (4) | (5) | (6) |
| Text Message | 0.035*** | 0.068*** | 0.034*** | 0.021*** | 0.038*** | 0.020*** |
| | (0.002) | (0.003) | (0.002) | (0.003) | (0.004) | (0.003) |
| Flu Shot | | | 0.039*** | | | 0.070*** |
| | | | (0.003) | | | (0.004) |
| Text Message x Flu Shot | | | 0.034*** | | | 0.017*** |
| | | | (0.004) | | | (0.005) |
| Gender-Female | -0.002 | -0.006* | -0.004* | -0.009*** | -0.011*** | -0.011*** |
| | (0.002) | (0.003) | (0.002) | (0.002) | (0.003) | (0.002) |
| Gender-Other | 0.016 | -0.097*** | 0.022 | 0.005 | -0.106*** | 0.021 |
| | (0.036) | (0.013) | (0.032) | (0.036) | (0.013) | (0.033) |
| Age | -0.002*** | -0.002*** | -0.002*** | -0.002*** | -0.003*** | -0.003*** |
| | (0.000) | (0.000) | (0.000) | (0.000) | (0.000) | (0.000) |
| Hispanic | 0.005 | 0.014* | 0.011** | 0.014** | 0.036*** | 0.028*** |
| | (0.004) | (0.006) | (0.004) | (0.005) | (0.007) | (0.004) |
| Non-Hispanic Black | 0.018*** | 0.072*** | 0.045*** | 0.035*** | 0.111*** | 0.072*** |
| | (0.006) | (0.008) | (0.005) | (0.006) | (0.009) | (0.006) |
| Non-Hispanic Asian | 0.002 | 0.014* | 0.009* | 0.001 | 0.025*** | 0.014*** |
| | (0.004) | (0.005) | (0.003) | (0.004) | (0.006) | (0.004) |
| Ethnicity-Other | -0.009* | -0.011 | -0.010** | -0.008* | -0.003 | -0.006 |
| | (0.004) | (0.006) | (0.003) | (0.004) | (0.007) | (0.004) |
| Ethnicity-Unknown | -0.020*** | -0.040*** | -0.028*** | -0.025*** | -0.053*** | -0.036*** |
| | (0.002) | (0.004) | (0.002) | (0.003) | (0.004) | (0.002) |
| Preferred Spanish | -0.009 | 0.024** | 0.008 | -0.007 | 0.044*** | 0.020*** |
| | (0.006) | (0.008) | (0.005) | (0.007) | (0.010) | (0.006) |
| Social Vulnerability Index (SVI) | -0.000*** | -0.001*** | -0.001*** | -0.001*** | -0.001*** | -0.001*** |
| | (0.000) | (0.000) | (0.000) | (0.000) | (0.000) | (0.000) |
| Missing SVI | -0.036*** | -0.057*** | -0.043*** | -0.053*** | -0.093*** | -0.067*** |
| | (0.002) | (0.005) | (0.003) | (0.003) | (0.006) | (0.003) |
| COVID-19 Risk Score | -0.000** | 0.000* | -0.000 | -0.000* | 0.000** | 0.000 |
| | (0.000) | (0.000) | (0.000) | (0.000) | (0.000) | (0.000) |
| Missing Risk Score | -0.027*** | -0.058*** | -0.038*** | -0.038*** | -0.073*** | -0.049*** |
| | (0.003) | (0.005) | (0.003) | (0.003) | (0.005) | (0.003) |
| $N$ | 55294 | 57935 | 113229 | 55294 | 57935 | 113229 |
| $R^2$ | 0.030 | 0.043 | 0.054 | 0.035 | 0.052 | 0.065 |

Robust standard errors are reported in parentheses. Controls for batch fixed effects are omitted from the regression table.
* $p < 0.05$, ** $p < 0.01$, *** $p < 0.001$

*Notes*: Heterogeneous treatment effects by whether patients got the flu shot in either the 2019-2020 or the 2020-2021 flu season based on their medical records. Columns 1 and 4 focus on patients who got the flu shot, Columns 2 and 5 focus on patients who did not get the flu shot, and Columns 3 and 6 focus on the combined sample. Text Message is a dummy variable coded as one when participants received one of the text messages in the first RCT.

Table 8: Effects of Simple Text Message (vs. Holdout) by Prior Flu Vaccination Status

| | Appointment at UCLA (Within Six Days) | | | Vaccinated at UCLA (Within Four Weeks) | | |
|---|---|---|---|---|---|---|
| | Flu Shot=0 | Flu Shot=1 | Both | Flu Shot=0 | Flu Shot=1 | Both |
| | (1) | (2) | (3) | (4) | (5) | (6) |
| Simple Text | 0.032*** | 0.062*** | 0.031*** | 0.020*** | 0.034*** | 0.019*** |
| | (0.003) | (0.004) | (0.003) | (0.003) | (0.005) | (0.003) |
| Flu Shot | | | 0.041*** | | | 0.071*** |
| | | | (0.003) | | | (0.004) |
| Simple Text x Flu Shot | | | 0.031*** | | | 0.014* |
| | | | (0.005) | | | (0.006) |
| Gender-Female | 0.004 | -0.006 | -0.002 | -0.005 | -0.010* | -0.009** |
| | (0.003) | (0.004) | (0.003) | (0.003) | (0.005) | (0.003) |
| Gender-Other | -0.010 | -0.084*** | 0.004 | -0.019 | -0.103*** | 0.005 |
| | (0.012) | (0.008) | (0.015) | (0.014) | (0.010) | (0.018) |
| Age | -0.001*** | -0.001*** | -0.001*** | -0.002*** | -0.002*** | -0.002*** |
| | (0.000) | (0.000) | (0.000) | (0.000) | (0.000) | (0.000) |
| Hispanic | -0.001 | 0.015 | 0.009 | 0.015 | 0.034*** | 0.027*** |
| | (0.006) | (0.009) | (0.006) | (0.008) | (0.010) | (0.007) |
| Non-Hispanic Black | 0.009 | 0.048*** | 0.028*** | 0.022* | 0.098*** | 0.059*** |
| | (0.008) | (0.012) | (0.007) | (0.009) | (0.014) | (0.009) |
| Non-Hispanic Asian | 0.008 | 0.008 | 0.008 | 0.011 | 0.022* | 0.017** |
| | (0.006) | (0.008) | (0.005) | (0.007) | (0.009) | (0.006) |
| Ethnicity-Other | -0.002 | -0.014 | -0.008 | -0.002 | -0.015 | -0.009 |
| | (0.006) | (0.008) | (0.005) | (0.007) | (0.010) | (0.006) |
| Ethnicity-Unknown | -0.018*** | -0.034*** | -0.025*** | -0.023*** | -0.046*** | -0.032*** |
| | (0.003) | (0.006) | (0.003) | (0.004) | (0.007) | (0.004) |
| Preferred Spanish | 0.000 | 0.042** | 0.021** | 0.009 | 0.050** | 0.030** |
| | (0.008) | (0.013) | (0.008) | (0.011) | (0.015) | (0.010) |
| Social Vulnerability Index (SVI) | -0.000*** | -0.001*** | -0.001*** | -0.001*** | -0.001*** | -0.001*** |
| | (0.000) | (0.000) | (0.000) | (0.000) | (0.000) | (0.000) |
| Missing SVI | -0.026*** | -0.053*** | -0.035*** | -0.048*** | -0.089*** | -0.062*** |
| | (0.004) | (0.008) | (0.004) | (0.004) | (0.008) | (0.004) |
| COVID-19 Risk Score | -0.000* | 0.000 | -0.000 | -0.000 | 0.000 | 0.000 |
| | (0.000) | (0.000) | (0.000) | (0.000) | (0.000) | (0.000) |
| Missing Risk Score | -0.024*** | -0.051*** | -0.033*** | -0.037*** | -0.072*** | -0.048*** |
| | (0.004) | (0.007) | (0.004) | (0.005) | (0.008) | (0.004) |
| *N* | 22106 | 23161 | 45267 | 22106 | 23161 | 45267 |
| $R^2$ | 0.031 | 0.041 | 0.051 | 0.036 | 0.049 | 0.064 |
| Conditions Included | Simple Text and Holdout | | | | | |

Robust standard errors are reported in parentheses. Controls for batch fixed effects are omitted from the regression table.
* $p < 0.05$, ** $p < 0.01$, *** $p < 0.001$

*Notes*: Heterogeneous treatment effects by whether patients got the flu shot in either the 2019-2020 or the 2020-2021 flu season based on their medical records. Columns 1 and 4 focus on patients who got the flu shot, Columns 2 and 5 focus on patients who did not get the flu shot, and Columns 3 and 6 focus on the combined sample. Simple Text is a dummy variable coded as one when participants received the *Simple Text* message in the first RCT.

Table 9: Effects of Ownership Messages (vs. Holdout) by Prior Flu Vaccination Status

| | Appointment at UCLA (Within Six Days) | | | Vaccinated at UCLA (Within Four Weeks) | | |
|---|---|---|---|---|---|---|
| | Flu Shot=0 | Flu Shot=1 | Both | Flu Shot=0 | Flu Shot=1 | Both |
| | (1) | (2) | (3) | (4) | (5) | (6) |
| Ownership | 0.039*** | 0.076*** | 0.039*** | 0.023*** | 0.044*** | 0.022*** |
| | (0.002) | (0.004) | (0.002) | (0.003) | (0.004) | (0.003) |
| Flu Shot | | | 0.039*** | | | 0.070*** |
| | | | (0.003) | | | (0.004) |
| Ownership x Flu Shot | | | 0.038*** | | | 0.021*** |
| | | | (0.004) | | | (0.005) |
| Gender-Female | -0.005 | -0.005 | -0.005* | -0.010*** | -0.012** | -0.011*** |
| | (0.003) | (0.004) | (0.002) | (0.003) | (0.004) | (0.003) |
| Gender-Other | -0.007 | -0.122*** | 0.008 | -0.021 | -0.129*** | 0.003 |
| | (0.011) | (0.008) | (0.017) | (0.012) | (0.009) | (0.019) |
| Age | -0.002*** | -0.002*** | -0.002*** | -0.002*** | -0.003*** | -0.003*** |
| | (0.000) | (0.000) | (0.000) | (0.000) | (0.000) | (0.000) |
| Hispanic | 0.002 | 0.018* | 0.012* | 0.006 | 0.038*** | 0.025*** |
| | (0.006) | (0.007) | (0.005) | (0.006) | (0.009) | (0.006) |
| Non-Hispanic Black | 0.025*** | 0.072*** | 0.049*** | 0.033*** | 0.108*** | 0.071*** |
| | (0.007) | (0.011) | (0.006) | (0.008) | (0.012) | (0.007) |
| Non-Hispanic Asian | 0.002 | 0.014 | 0.009* | -0.001 | 0.019* | 0.011* |
| | (0.005) | (0.007) | (0.004) | (0.006) | (0.008) | (0.005) |
| Ethnicity-Other | -0.012* | -0.011 | -0.011** | -0.005 | -0.002 | -0.003 |
| | (0.005) | (0.007) | (0.004) | (0.006) | (0.009) | (0.005) |
| Ethnicity-Unknown | -0.019*** | -0.043*** | -0.029*** | -0.023*** | -0.052*** | -0.035*** |
| | (0.003) | (0.005) | (0.003) | (0.003) | (0.006) | (0.003) |
| Preferred Spanish | -0.005 | 0.017 | 0.006 | -0.007 | 0.046*** | 0.022** |
| | (0.007) | (0.011) | (0.007) | (0.008) | (0.012) | (0.008) |
| Social Vulnerability Index (SVI) | -0.000*** | -0.001*** | -0.001*** | -0.000*** | -0.001*** | -0.001*** |
| | (0.000) | (0.000) | (0.000) | (0.000) | (0.000) | (0.000) |
| Missing SVI | -0.037*** | -0.062*** | -0.046*** | -0.052*** | -0.096*** | -0.067*** |
| | (0.003) | (0.007) | (0.003) | (0.003) | (0.007) | (0.004) |
| COVID-19 Risk Score | -0.000* | 0.000 | -0.000 | -0.000 | 0.000 | -0.000 |
| | (0.000) | (0.000) | (0.000) | (0.000) | (0.000) | (0.000) |
| Missing Risk Score | -0.028*** | -0.054*** | -0.037*** | -0.037*** | -0.067*** | -0.046*** |
| | (0.004) | (0.006) | (0.003) | (0.004) | (0.007) | (0.004) |
| *N* | 33128 | 34826 | 67954 | 33128 | 34826 | 67954 |
| $R^2$ | 0.035 | 0.048 | 0.058 | 0.037 | 0.053 | 0.067 |
| Conditions Included | Ownership Text, Ownership Text w/ Video, and Holdout | | | | | |

Robust standard errors are reported in parentheses. Controls for batch fixed effects are omitted from the regression table.
* $p < 0.05$, ** $p < 0.01$, *** $p < 0.001$

*Notes*: Heterogeneous treatment effects by whether patients got the flu shot in either the 2019-2020 or the 2020-2021 flu season based on their medical records. Columns 1 and 4 focus on patients who got the flu shot, Columns 2 and 5 focus on patients who did not get the flu shot, and Columns 3 and 6 focus on the combined sample. Ownership is a dummy variable coded as one when participants received one of the messages containing the ownership language in the first RCT.

Table 10: Effects of Video Messages (vs. Holdout) by Prior Flu Vaccination Status

| | Appointment at UCLA (Within Six Days) | | | Vaccinated at UCLA (Within Four Weeks) | | |
|---|---|---|---|---|---|---|
| | Flu Shot=0 | Flu Shot=1 | Both | Flu Shot=0 | Flu Shot=1 | Both |
| | (1) | (2) | (3) | (4) | (5) | (6) |
| Video | 0.035*** | 0.066*** | 0.034*** | 0.022*** | 0.036*** | 0.021*** |
| | (0.002) | (0.003) | (0.002) | (0.003) | (0.004) | (0.003) |
| Flu Shot | | | 0.040*** | | | 0.071*** |
| | | | (0.003) | | | (0.004) |
| Video x Flu Shot | | | 0.032*** | | | 0.015** |
| | | | (0.004) | | | (0.005) |
| Gender-Female | -0.002 | -0.007 | -0.005* | -0.010*** | -0.011** | -0.012*** |
| | (0.002) | (0.004) | (0.002) | (0.003) | (0.004) | (0.003) |
| Gender-Other | 0.037 | -0.102*** | 0.041 | 0.023 | -0.112*** | 0.041 |
| | (0.056) | (0.008) | (0.050) | (0.056) | (0.009) | (0.051) |
| Age | -0.001*** | -0.002*** | -0.002*** | -0.002*** | -0.003*** | -0.002*** |
| | (0.000) | (0.000) | (0.000) | (0.000) | (0.000) | (0.000) |
| Hispanic | 0.006 | 0.010 | 0.009 | 0.014* | 0.037*** | 0.028*** |
| | (0.006) | (0.007) | (0.005) | (0.007) | (0.008) | (0.006) |
| Non-Hispanic Black | 0.016* | 0.069*** | 0.042*** | 0.034*** | 0.116*** | 0.074*** |
| | (0.007) | (0.010) | (0.006) | (0.008) | (0.012) | (0.007) |
| Non-Hispanic Asian | 0.007 | 0.019** | 0.014** | 0.005 | 0.025** | 0.016*** |
| | (0.005) | (0.007) | (0.004) | (0.006) | (0.008) | (0.005) |
| Ethnicity-Other | -0.007 | -0.007 | -0.006 | 0.001 | 0.003 | 0.003 |
| | (0.005) | (0.007) | (0.004) | (0.006) | (0.008) | (0.005) |
| Ethnicity-Unknown | -0.020*** | -0.043*** | -0.030*** | -0.021*** | -0.052*** | -0.034*** |
| | (0.003) | (0.005) | (0.003) | (0.003) | (0.005) | (0.003) |
| Preferred Spanish | -0.007 | 0.018 | 0.006 | -0.006 | 0.033** | 0.016* |
| | (0.007) | (0.010) | (0.006) | (0.009) | (0.012) | (0.008) |
| Social Vulnerability Index (SVI) | -0.000*** | -0.001*** | -0.001*** | -0.001*** | -0.001*** | -0.001*** |
| | (0.000) | (0.000) | (0.000) | (0.000) | (0.000) | (0.000) |
| Missing SVI | -0.031*** | -0.043*** | -0.035*** | -0.048*** | -0.081*** | -0.060*** |
| | (0.003) | (0.007) | (0.003) | (0.004) | (0.007) | (0.004) |
| COVID-19 Risk Score | -0.000** | 0.000 | -0.000 | -0.000* | 0.000 | -0.000 |
| | (0.000) | (0.000) | (0.000) | (0.000) | (0.000) | (0.000) |
| Missing Risk Score | -0.024*** | -0.060*** | -0.037*** | -0.036*** | -0.074*** | -0.049*** |
| | (0.004) | (0.006) | (0.003) | (0.004) | (0.007) | (0.004) |
| $N$ | 33110 | 34880 | 67990 | 33110 | 34880 | 67990 |
| $R^2$ | 0.030 | 0.045 | 0.053 | 0.033 | 0.051 | 0.063 |
| Conditions Included | Simple Text w/ Video, Ownership Text w/ Video, and Holdout | | | | | |

Robust standard errors are reported in parentheses. Controls for batch fixed effects are omitted from the regression table.
* $p < 0.05$, ** $p < 0.01$, *** $p < 0.001$

*Notes*: Heterogeneous treatment effects by whether patients got the flu shot in either the 2019-2020 or the 2020-2021 flu season based on their medical records. Columns 1 and 4 focus on patients who got the flu shot, Columns 2 and 5 focus on patients who did not get the flu shot, and Columns 3 and 6 focus on the combined sample. Video is a dummy variable coded as one when participants received one of the messages containing a link to the video in the first RCT.

Table 11: Effects of Ownership Language and Video by Prior Flu Vaccination Status

| | Appointment at UCLA (Within Six Days) | | | Vaccinated at UCLA (Within Four Weeks) | | |
|---|---|---|---|---|---|---|
| | Flu Shot=0 | Flu Shot=1 | Both | Flu Shot=0 | Flu Shot=1 | Both |
| | (1) | (2) | (3) | (4) | (5) | (6) |
| Ownership | 0.009*** | 0.016*** | 0.009*** | 0.004 | 0.012*** | 0.004 |
| | (0.002) | (0.003) | (0.002) | (0.003) | (0.004) | (0.003) |
| Video | -0.000 | -0.004 | -0.000 | 0.001 | -0.003 | 0.001 |
| | (0.002) | (0.003) | (0.002) | (0.003) | (0.004) | (0.003) |
| Flu Shot | | | 0.070*** | | | 0.085*** |
| | | | (0.003) | | | (0.004) |
| Ownership x Flu Shot | | | 0.007 | | | 0.008 |
| | | | (0.004) | | | (0.004) |
| Video x Flu Shot | | | -0.003 | | | -0.004 |
| | | | (0.004) | | | (0.004) |
| Gender-Female | -0.004 | -0.007* | -0.006** | -0.011*** | -0.011** | -0.012*** |
| | (0.002) | (0.003) | (0.002) | (0.003) | (0.004) | (0.002) |
| Gender-Other | 0.019 | -0.095*** | 0.018 | 0.010 | -0.105*** | 0.018 |
| | (0.050) | (0.023) | (0.043) | (0.050) | (0.019) | (0.044) |
| Age | -0.002*** | -0.003*** | -0.002*** | -0.002*** | -0.003*** | -0.003*** |
| | (0.000) | (0.000) | (0.000) | (0.000) | (0.000) | (0.000) |
| Hispanic | 0.007 | 0.014* | 0.012** | 0.016** | 0.035*** | 0.028*** |
| | (0.005) | (0.007) | (0.004) | (0.006) | (0.007) | (0.005) |
| Non-Hispanic Black | 0.020** | 0.082*** | 0.051*** | 0.039*** | 0.113*** | 0.076*** |
| | (0.006) | (0.010) | (0.006) | (0.007) | (0.010) | (0.006) |
| Non-Hispanic Asian | -0.001 | 0.014* | 0.008* | -0.003 | 0.028*** | 0.014** |
| | (0.005) | (0.006) | (0.004) | (0.005) | (0.007) | (0.004) |
| Ethnicity-Other | -0.010* | -0.009 | -0.009* | -0.012* | -0.001 | -0.006 |
| | (0.004) | (0.007) | (0.004) | (0.005) | (0.008) | (0.004) |
| Ethnicity-Unknown | -0.021*** | -0.040*** | -0.029*** | -0.026*** | -0.055*** | -0.038*** |
| | (0.003) | (0.005) | (0.003) | (0.003) | (0.005) | (0.003) |
| Preferred Spanish | -0.012 | 0.023* | 0.007 | -0.012 | 0.049*** | 0.021** |
| | (0.006) | (0.010) | (0.006) | (0.008) | (0.011) | (0.007) |
| Social Vulnerability Index (SVI) | -0.001*** | -0.001*** | -0.001*** | -0.001*** | -0.001*** | -0.001*** |
| | (0.000) | (0.000) | (0.000) | (0.000) | (0.000) | (0.000) |
| Missing SVI | -0.041*** | -0.063*** | -0.048*** | -0.056*** | -0.098*** | -0.070*** |
| | (0.003) | (0.006) | (0.003) | (0.003) | (0.006) | (0.003) |
| COVID-19 Risk Score | -0.000** | 0.000** | 0.000 | -0.000 | 0.000*** | 0.000 |
| | (0.000) | (0.000) | (0.000) | (0.000) | (0.000) | (0.000) |
| Missing Risk Score | -0.029*** | -0.063*** | -0.041*** | -0.039*** | -0.076*** | -0.051*** |
| | (0.004) | (0.006) | (0.003) | (0.004) | (0.006) | (0.003) |
| $N$ | 44270 | 46280 | 90550 | 44270 | 46280 | 90550 |
| $R^2$ | 0.028 | 0.040 | 0.052 | 0.034 | 0.053 | 0.066 |
| Conditions Included | Conditions within the Text Message arm: Simple Text, Ownership Text, Simple Text w/ Video, and Ownership Text w/ Video | | | | | |

Robust standard errors are reported in parentheses. Controls for batch fixed effects are omitted from the regression table.
* $p < 0.05$, ** $p < 0.01$, *** $p < 0.001$

*Notes*: Heterogeneous treatment effects by whether patients got the flu shot in either the 2019-2020 or the 2020-2021 flu season based on their medical records. Columns 1 and 4 focus on patients who got the flu shot, Columns 2 and 5 focus on patients who did not get the flu shot, and Columns 3 and 6 focus on the combined sample. Ownership is a dummy variable coded as one when participants received one of the messages containing the ownership language in the first RCT. Video is a dummy variable coded as one when participants received one of the messages containing a link to the video in the first RCT.

##### 1.3.5 Generalizability Across Racial/Ethnic and Age Groups

Given that our sample has predominantly non-Hispanic white patients and recent national surveys involving US adults (Funk and Tyson, 2020; Hamel et al., 2020; Ruiz and Bell, 2021) suggest that racial/ethnic minority groups (such as non-Hispanic African American and Hispanic people) have weaker intentions to get vaccinated for COVID-19, we explored whether the average effect of our text message and the effect of ownership language hold for both white and racial/ethnic minority patients (including Hispanic patients, non-Hispanic African American patients, non-Hispanic Asian patients, and patients whose race/ethnicity is categorized as other or mixed race). As shown in Table 12 Columns 1-2 and 5-6, receiving a text message significantly boosted appointment rates and vaccination rates for non-Hispanic white patients (5.16 and 2.99 percentage points; ps $< 0.001$) and racial/ethnic minority patients (5.57 and 3.16 percentage points; ps $< 0.001$). Columns 3 and 4 show that the inclusion of ownership language produced additional benefits for appointment scheduling among non-Hispanic white patients (1.23 percentage points; $p < 0.001$) and racial/ethnic minority patients (1.53 percentage points; $p < 0.001$). Columns 7 and 8 suggest that the inclusion of ownership language further boosted vaccination rates among non-Hispanic white patients (0.95 percentage points; $p = 0.002$), and this effect is positive but not statistically significant among racial/ethnic minority patients (0.67 percentage points; $p = 0.13$).

Importantly, we also confirmed that the average effect of receiving a text message was all statistically significant and had a comparable magnitude across white (Table 12 Columns 1 and 5), Hispanic (Table 13 Columns 1 and 4), African American (Table 13 Columns 2 and 5), and Asian patients (Table 13 Columns 3 and 6).

We also explored whether the average effect of our text message and the effect of ownership language hold for both patients at or above 65 years old (the vast majority of our sample) and patients below 65 years old (to whom vaccines will become available in the near future in U.S. at the time of our data extraction). We applied regression specifications (1) and (5) separately to these subgroups of patients. As shown in Table 14 Columns 1-2 and 5-6, receiving a text message significantly boosted appointment rates and vaccination rates for both patients at or above 65 years old (4.67 and 2.58 percentage points; ps $< 0.001$) and patients below 65 (9.50 and 5.81 percentage points; ps $< 0.001$). Also, Columns 3-4 and 7-8 show that the additional benefits of ownership language for appointment rates and vaccination rates held among patients at or above 65 years old (1.00 and 0.71 percentage points; ps $< 0.001$) and patients below 65 (3.53 and 2.00 percentage points; ps $< 0.03$).

Table 12: Effects of Receiving a Text Message and Ownership Language among non-Hispanic White Patients vs. Minority Patients

| | Appointment at UCLA (Within Six Days) | | | | Vaccinated at UCLA (Within Four Weeks) | | | |
|---|---|---|---|---|---|---|---|---|
| | White | Minority | White | Minority | White | Minority | White | Minority |
| | (1) | (2) | (3) | (4) | (5) | (6) | (7) | (8) |
| Text Message | 0.052*** | 0.056*** | | | 0.030*** | 0.032*** | | |
| | (0.003) | (0.004) | | | (0.003) | (0.005) | | |
| Ownership | | | 0.012*** | 0.015*** | | | 0.010** | 0.007 |
| | | | (0.003) | (0.004) | | | (0.003) | (0.004) |
| Video | | | -0.002 | 0.001 | | | -0.004 | 0.004 |
| | | | (0.003) | (0.004) | | | (0.003) | (0.004) |
| Gender-Female | -0.009*** | 0.002 | -0.011*** | 0.001 | -0.015*** | -0.006 | -0.016*** | -0.007 |
| | (0.002) | (0.003) | (0.003) | (0.004) | (0.003) | (0.004) | (0.003) | (0.005) |
| Gender-Other | 0.112 | -0.070*** | 0.114 | -0.070*** | 0.100 | -0.076*** | 0.102 | -0.076*** |
| | (0.151) | (0.008) | (0.150) | (0.011) | (0.150) | (0.008) | (0.151) | (0.011) |
| Age | -0.003*** | -0.002*** | -0.003*** | -0.002*** | -0.003*** | -0.003*** | -0.003*** | -0.003*** |
| | (0.000) | (0.000) | (0.000) | (0.000) | (0.000) | (0.000) | (0.000) | (0.000) |
| Hispanic | 0.000 | 0.028*** | 0.000 | 0.028*** | 0.000 | 0.040*** | 0.000 | 0.041*** |
| | (.) | (0.005) | (.) | (0.006) | (.) | (0.006) | (.) | (0.007) |
| Non-Hispanic Black | 0.000 | 0.057*** | 0.000 | 0.062*** | 0.000 | 0.080*** | 0.000 | 0.085*** |
| | (.) | (0.006) | (.) | (0.007) | (.) | (0.007) | (.) | (0.008) |
| Non-Hispanic Asian | 0.000 | 0.023*** | 0.000 | 0.022*** | 0.000 | 0.025*** | 0.000 | 0.026*** |
| | (.) | (0.005) | (.) | (0.005) | (.) | (0.005) | (.) | (0.006) |
| Ethnicity-Other | 0.000 | 0.000 | 0.000 | 0.000 | 0.000 | 0.000 | 0.000 | 0.000 |
| | (.) | (.) | (.) | (.) | (.) | (.) | (.) | (.) |
| Ethnicity-Unknown | 0.000 | 0.000 | 0.000 | 0.000 | 0.000 | 0.000 | 0.000 | 0.000 |
| | (.) | (.) | (.) | (.) | (.) | (.) | (.) | (.) |
| Preferred Spanish | -0.031 | 0.011 | -0.032 | 0.010 | -0.001 | 0.027*** | -0.004 | 0.028*** |
| | (0.019) | (0.006) | (0.022) | (0.007) | (0.026) | (0.007) | (0.029) | (0.008) |
| Social Vulnerability Index (SVI) | -0.001*** | -0.001*** | -0.001*** | -0.001*** | -0.001*** | -0.001*** | -0.001*** | -0.001*** |
| | (0.000) | (0.000) | (0.000) | (0.000) | (0.000) | (0.000) | (0.000) | (0.000) |
| Missing SVI | -0.055*** | -0.061*** | -0.061*** | -0.067*** | -0.079*** | -0.098*** | -0.083*** | -0.103*** |
| | (0.003) | (0.006) | (0.004) | (0.007) | (0.004) | (0.006) | (0.004) | (0.007) |
| COVID-19 Risk Score | 0.000 | -0.000 | 0.000 | -0.000 | 0.000 | -0.000 | 0.000 | -0.000 |
| | (0.000) | (0.000) | (0.000) | (0.000) | (0.000) | (0.000) | (0.000) | (0.000) |
| Missing Risk Score | -0.048*** | -0.062*** | -0.051*** | -0.068*** | -0.059*** | -0.086*** | -0.062*** | -0.086*** |
| | (0.004) | (0.006) | (0.004) | (0.007) | (0.004) | (0.006) | (0.004) | (0.007) |
| $N$ | 62856 | 34384 | 50154 | 27597 | 62856 | 34384 | 50154 | 27597 |
| $R^2$ | 0.041 | 0.037 | 0.040 | 0.035 | 0.047 | 0.046 | 0.047 | 0.046 |
| Conditions Included | All | | Conditions within the Text Message arm | | All | | Conditions within the Text Message arm | |

Robust standard errors are reported in parentheses. Controls for batch fixed effects are omitted from the regression table.

* $p < 0.05$, ** $p < 0.01$, *** $p < 0.001$

*Notes*: Columns 1, 3, 5, and 7 focus on non-Hispanic white patients, and Columns 2, 4, 6, and 8 focus on minority patients. Text Message is a dummy variable coded as one when participants received one of the text messages in the first RCT. Ownership is a dummy variable coded as one when participants received one of the messages containing the ownership language in the first RCT. Video is a dummy variable coded as one when participants received one of the messages containing a link to the video in the first RCT.

Table 13: Effects of Receiving a Text Message by Minority Subgroups

| | Appointment at UCLA (Within Six Days) | | | Vaccinated at UCLA (Within Four Weeks) | | |
|---|---|---|---|---|---|---|
| | Hispanic | African American | Asian | Hispanic | African American | Asian |
| | (1) | (2) | (3) | (4) | (5) | (6) |
| Text Message | 0.054*** | 0.079*** | 0.047*** | 0.033*** | 0.047*** | 0.027** |
| | (0.007) | (0.010) | (0.007) | (0.008) | (0.013) | (0.009) |
| Gender-Female | -0.002 | 0.011 | 0.011 | -0.011 | -0.001 | 0.009 |
| | (0.006) | (0.010) | (0.007) | (0.007) | (0.011) | (0.008) |
| Gender-Other | | | -0.072*** | | | -0.085*** |
| | | | (0.016) | | | (0.017) |
| Age | -0.001* | -0.001 | -0.003*** | -0.002*** | -0.002** | -0.004*** |
| | (0.000) | (0.000) | (0.000) | (0.000) | (0.001) | (0.000) |
| Hispanic | 0.000 | 0.000 | 0.000 | 0.000 | 0.000 | 0.000 |
| | (.) | (.) | (.) | (.) | (.) | (.) |
| Non-Hispanic Black | 0.000 | 0.000 | 0.000 | 0.000 | 0.000 | 0.000 |
| | (.) | (.) | (.) | (.) | (.) | (.) |
| Non-Hispanic Asian | 0.000 | 0.000 | 0.000 | 0.000 | 0.000 | 0.000 |
| | (.) | (.) | (.) | (.) | (.) | (.) |
| Ethnicity-Other | 0.000 | 0.000 | 0.000 | 0.000 | 0.000 | 0.000 |
| | (.) | (.) | (.) | (.) | (.) | (.) |
| Ethnicity-Unknown | 0.000 | 0.000 | 0.000 | 0.000 | 0.000 | 0.000 |
| | (.) | (.) | (.) | (.) | (.) | (.) |
| Preferred Spanish | 0.012 | -0.209*** | 0.273 | 0.028*** | -0.291*** | 0.437* |
| | (0.007) | (0.026) | (0.224) | (0.008) | (0.029) | (0.221) |
| Social Vulnerability Index (SVI) | -0.001*** | -0.001*** | -0.001*** | -0.001*** | -0.001*** | -0.001*** |
| | (0.000) | (0.000) | (0.000) | (0.000) | (0.000) | (0.000) |
| Missing SVI | -0.086*** | -0.055*** | -0.059*** | -0.129*** | -0.112*** | -0.088*** |
| | (0.011) | (0.016) | (0.012) | (0.012) | (0.017) | (0.013) |
| COVID-19 Risk Score | -0.000 | -0.000 | 0.000 | -0.000 | -0.000 | -0.000 |
| | (0.000) | (0.000) | (0.000) | (0.000) | (0.000) | (0.000) |
| Missing Risk Score | -0.078*** | -0.066** | -0.063*** | -0.113*** | -0.112*** | -0.070*** |
| | (0.011) | (0.022) | (0.010) | (0.013) | (0.024) | (0.012) |
| *N* | 11758 | 5458 | 9182 | 11758 | 5458 | 9182 |
| $R^2$ | 0.037 | 0.034 | 0.048 | 0.046 | 0.043 | 0.054 |

Robust standard errors are reported in parentheses. Controls for batch fixed effects are omitted from the regression table.

$^{*}$ $p < 0.05$, $^{**}$ $p < 0.01$, $^{***}$ $p < 0.001$

*Notes*: Columns 1 and 4 focus on Hispanic patients, Columns 2 and 5 focus on African American patients, and Columns 3 and 6 focus on Asian patients. Text Message is a dummy variable coded as one when participants received one of the text messages in the first RCT.

Table 14: Effects of Receiving a Text Message and Ownership Language by Patient Age

| | Appointment at UCLA (Within Six Days) | | | | Vaccinated at UCLA (Within Four Weeks) | | | |
|---|---|---|---|---|---|---|---|---|
| | Age $\geq$ 65 | Age < 65 | Age $\geq$ 65 | Age < 65 | Age $\geq$ 65 | Age < 65 | Age $\geq$ 65 | Age < 65 |
| | (1) | (2) | (3) | (4) | (5) | (6) | (7) | (8) |
| Text Message | 0.047*** | 0.095*** | | | 0.026*** | 0.058*** | | |
| | (0.002) | (0.008) | | | (0.002) | (0.010) | | |
| Ownership | | | 0.010*** | 0.035*** | | | 0.007** | 0.020* |
| | | | (0.002) | (0.009) | | | (0.002) | (0.009) |
| Video | | | -0.002 | -0.006 | | | -0.002 | 0.001 |
| | | | (0.002) | (0.009) | | | (0.002) | (0.009) |
| Gender-Female | -0.001 | -0.018* | -0.002 | -0.024** | -0.005** | -0.031*** | -0.006* | -0.035*** |
| | (0.002) | (0.008) | (0.002) | (0.009) | (0.002) | (0.008) | (0.002) | (0.010) |
| Gender-Other | -0.040*** | 0.943*** | -0.056*** | 0.979*** | -0.050*** | 0.983*** | -0.063*** | 1.001*** |
| | (0.010) | (0.018) | (0.009) | (0.022) | (0.011) | (0.020) | (0.011) | (0.023) |
| Age | -0.003*** | 0.002*** | -0.003*** | 0.002*** | -0.003*** | 0.003*** | -0.003*** | 0.003*** |
| | (0.000) | (0.000) | (0.000) | (0.000) | (0.000) | (0.000) | (0.000) | (0.000) |
| Hispanic | 0.011** | 0.003 | 0.012* | 0.001 | 0.023*** | 0.025 | 0.024*** | 0.016 |
| | (0.004) | (0.012) | (0.005) | (0.014) | (0.005) | (0.013) | (0.005) | (0.015) |
| Non-Hispanic Black | 0.050*** | -0.025 | 0.056*** | -0.026 | 0.075*** | -0.006 | 0.078*** | -0.005 |
| | (0.005) | (0.014) | (0.006) | (0.017) | (0.006) | (0.016) | (0.007) | (0.018) |
| Non-Hispanic Asian | 0.008* | 0.025 | 0.008 | 0.016 | 0.012** | 0.044** | 0.013** | 0.032 |
| | (0.004) | (0.015) | (0.004) | (0.017) | (0.004) | (0.016) | (0.004) | (0.018) |
| Ethnicity-Other | -0.012*** | -0.027 | -0.013*** | -0.016 | -0.007 | -0.043** | -0.010* | -0.036 |
| | (0.003) | (0.016) | (0.004) | (0.019) | (0.004) | (0.017) | (0.004) | (0.019) |
| Ethnicity-Unknown | -0.027*** | -0.078*** | -0.027*** | -0.078*** | -0.035*** | -0.082*** | -0.037*** | -0.081*** |
| | (0.002) | (0.012) | (0.003) | (0.014) | (0.002) | (0.013) | (0.003) | (0.015) |
| Preferred Spanish | 0.009 | -0.009 | 0.007 | -0.010 | 0.018** | 0.019 | 0.018* | 0.023 |
| | (0.005) | (0.016) | (0.006) | (0.018) | (0.006) | (0.019) | (0.007) | (0.021) |
| Social Vulnerability Index (SVI) | -0.001*** | -0.001*** | -0.001*** | -0.001*** | -0.001*** | -0.001*** | -0.001*** | -0.001*** |
| | (0.000) | (0.000) | (0.000) | (0.000) | (0.000) | (0.000) | (0.000) | (0.000) |
| Missing SVI | -0.044*** | -0.132*** | -0.049*** | -0.146*** | -0.067*** | -0.181*** | -0.071*** | -0.192*** |
| | (0.003) | (0.013) | (0.003) | (0.015) | (0.003) | (0.014) | (0.003) | (0.016) |
| COVID-19 Risk Score | -0.000 | 0.000 | -0.000 | 0.000 | -0.000* | 0.000** | -0.000* | 0.000** |
| | (0.000) | (0.000) | (0.000) | (0.000) | (0.000) | (0.000) | (0.000) | (0.000) |
| Missing Risk Score | -0.032*** | -0.093*** | -0.034*** | -0.097*** | -0.039*** | -0.114*** | -0.041*** | -0.115*** |
| | (0.003) | (0.011) | (0.004) | (0.013) | (0.003) | (0.012) | (0.004) | (0.014) |
| $N$ | 101886 | 11343 | 81426 | 9124 | 101886 | 11343 | 81426 | 9124 |
| $R^2$ | 0.032 | 0.043 | 0.030 | 0.037 | 0.045 | 0.037 | 0.045 | 0.036 |
| Conditions Included | All | | Conditions within the Text Message arm | | All | | Conditions within the Text Message arm | |

Robust standard errors are reported in parentheses. Controls for batch fixed effects are omitted from the regression table.

* $p < 0.05$, ** $p < 0.01$, *** $p < 0.001$

*Notes*: Columns 1, 3, 5, and 7 focus on patients at or above 65 years old, and Columns 2, 4, 6, and 8 focus on patients below 65. Text Message is a dummy variable coded as one when participants received one of the text messages in the first RCT. Ownership is a dummy variable coded as one when participants received one of the messages containing the ownership language in the first RCT. Video is a dummy variable coded as one when participants received one of the messages containing a link to the video in the first RCT.

#### 1.4 Results about the Second RCT

Following the pre-registration, our early analyses about the second RCT sought to answer the following questions:

1. What is the average effect of sending behaviorally informed text reminders among patients in the second RCT?

2. Do all types of text messages—including the three messages mentioning personal benefits, the three messages emphasizing prosocial benefits, the two messages highlighting exclusive access to the vaccine, and the two messages referencing the opportunity for a fresh start—all outperform the *Holdout* arm?

Because we were uncertain about how many people would be qualified for the early report of the second RCT, we were concerned that we might not have enough power to test the main effects of prosocial messaging, exclusivity framing, and fresh start framing as well as their interaction. Thus, we pre-registered to only answer those questions and conduct hypothesis testing between sub-arms once data collection has been completed.

In addition to reporting regression analyses, we also pre-registered that we would show the raw data for each condition without conducting hypothesis testing. Figure 3 in Supplementary Information displays the average appointment rate and the average vaccination completion rate at UCLA Health in each of the seven conditions (six sub-arms within the *Text Message* arm plus the *Holdout* arm).

##### 1.4.1 The Average Effect of Receiving the Second Text Reminder

We used the same regression specification as equation (1) to estimate the average effect of receiving the second text reminder. As shown in Table 15 Column 1, receiving the second text reminder on average significantly boosted patients' likelihood of scheduling an appointment at UCLA Health within six days of the second reminder date by 1.26 percentage points (B = 0.013, SE = 0.002, $p < 0.001$). This amounts to a 43.08% increase relative to the average appointment rates of 2.41 percentage points in the *Holdout* arm. Also, as shown in Column 2, our second text reminders on average significantly boosted patients' likelihood of getting the first dose at UCLA Health within four weeks of the second reminder date by 0.68 percentage points (B = 0.068, SE = 0.002, $p < 0.001$). This corresponds to a 15.89% increase relative to the average vaccination rates of 4.28 percentage points in the *Holdout* arm.

Among patients in the *Text Message* arm of the second RCT, only 20.67% were in the *Holdout* arm of the first RCT, to whom the text message sent in the second RCT was the first reminder they received from UCLA Health. When we exclude from analysis patients in the *Holdout* arm of the first RCT, the aforementioned finding about the average effect of receiving a text reminder in the second RCT was basically unchanged (as shown in Table 15 Columns 3-4). This suggests that there is indeed a significant, positive effect of sending a *second* text reminder.

##### 1.4.2 Comparing Different Types of Text Reminders with Holdout

After showing that our second text reminders overall boosted both appointment rates and vaccination rates, we tested whether all types of messages—the three reminders mentioning personal benefits, the three reminders emphasizing prosocial benefits, the two reminders highlighting patients' early access to the vaccine, and the two reminders referencing the opportunity for a fresh start—outperformed the *Holdout* arm. To answer this question, we conducted four regressions:

$$Y_i = \beta_0 + \beta_1 Self_i + \beta_2 X_i + \epsilon_i, \tag{6}$$

Specification (6) only includes patients in the *Holdout* arm, the *Simple Self* condition, the *Early Access Self* condition, and the *Fresh Start Self* condition. $Self_i$ equals one if patient $i$ was assigned to the three latter conditions that highlighted personal benefits and zero otherwise.

$$Y_i = \beta_0 + \beta_1 Prosocial_i + \beta_2 X_i + \epsilon_i, \tag{7}$$

Specification (7) only includes patients in the *Holdout* arm, the *Simple Prosocial* condition, the *Early Access Prosocial* condition, and the *Fresh Start Prosocial* condition. $Prosocial_i$ equals one if patient $i$ was assigned to the three latter conditions that highlighted prosocial benefits and zero otherwise.

$$Y_i = \beta_0 + \beta_1 Early\ Access_i + \beta_2 X_i + \epsilon_i, \tag{8}$$

Specification (8) only includes patients in the *Holdout* arm, the *Early Access Self* condition, and the *Early Access Prosocial* condition. $EarlyAccess_i$ equals one if patient $i$ was assigned to the two latter conditions that highlighted their early access to the vaccine and zero, otherwise.

$$Y_i = \beta_0 + \beta_1 Fresh\ Start_i + \beta_2 X_i + \epsilon_i, \tag{9}$$

Specification (9) only includes patients in the *Holdout* arm, the *Fresh Start Self* condition, and the *Fresh Start Prosocial* condition. $FreshStart_i$ equals one if patient $i$ was assigned to the two latter conditions that framed getting the vaccine as an opportunity for having a fresh start and zero, otherwise.

Across specifications (6)-(9), $Y_i$ represents the outcome variable of patient $i$ (including *Appointment at UCLA* and *Vaccinated at UCLA*). $X_i$ represents the same set of pre-registered control variables as mentioned in equation 1. We report robust standard errors.

Table 16 reports the results from specifications (6)-(9). First, the three text reminders that referenced personal benefits significantly boosted both appointment rates (by 1.17 percentage points or 48.58%) and vaccination rates (by 0.54 percentage points or 12.51%), relative to the *Holdout* arm. Second, the three text reminders that referenced prosocial benefits significantly boosted both appointment rates (by 1.34 percentage points or 55.63%) and vaccination rates (by 0.81 percentage points or 19.01%), relative to the *Holdout* arm. Also, the two text reminders highlighting patients' early access to the vaccine significantly outperformed the *Holdout* arm in both appointment rates (by 1.50 percentage points or 62.31%) and vaccination rates (by 0.09 percentage points or 21.29%). Further, the two text reminders containing the fresh start framing significantly lifted appointment rates (by 0.98 percentage points or 40.54%) and marginally significantly boosted vaccination rates (by 0.05 percentage points or 10.98%), compared to the *Holdout* arm.

Table 15: Effects of Receiving the Second Text Message on Appointments and Vaccinations at UCLA Health

| | Appointment at UCLA (Within Six Days) | | Vaccinated at UCLA (Within Four Weeks) | |
|---|---|---|---|---|
| | All Patients | Excl. Patients in the Holdout Arm of the first RCT | All Patients | Excl. Patients in the Holdout Arm of the first RCT |
| | (1) | (2) | (3) | (4) |
| Text Message | 0.013*** | 0.010*** | 0.007*** | 0.006** |
| | (0.002) | (0.002) | (0.002) | (0.002) |
| Gender-Female | -0.001 | -0.001 | -0.003* | -0.003 |
| | (0.001) | (0.001) | (0.001) | (0.002) |
| Gender-Other | -0.013*** | -0.012*** | -0.016*** | -0.017*** |
| | (0.003) | (0.004) | (0.004) | (0.004) |
| Age | -0.001*** | -0.001*** | -0.001*** | -0.001*** |
| | (0.000) | (0.000) | (0.000) | (0.000) |
| Hispanic | 0.013*** | 0.009** | 0.021*** | 0.020*** |
| | (0.003) | (0.003) | (0.003) | (0.004) |
| Non-Hispanic Black | 0.016*** | 0.013*** | 0.035*** | 0.034*** |
| | (0.004) | (0.004) | (0.005) | (0.005) |
| Non-Hispanic Asian | 0.005* | 0.005 | 0.009*** | 0.010** |
| | (0.002) | (0.003) | (0.003) | (0.003) |
| Ethnicity-Other | 0.001 | -0.000 | 0.005 | 0.005 |
| | (0.002) | (0.003) | (0.003) | (0.003) |
| Ethnicity-Unknown | -0.007*** | -0.009*** | -0.010*** | -0.010*** |
| | (0.001) | (0.002) | (0.002) | (0.002) |
| Preferred Spanish | 0.008* | 0.009* | 0.013** | 0.015** |
| | (0.004) | (0.004) | (0.005) | (0.005) |
| Social Vulnerability Index (SVI) | -0.000*** | -0.000*** | -0.000*** | -0.000*** |
| | (0.000) | (0.000) | (0.000) | (0.000) |
| Missing SVI | -0.022*** | -0.020*** | -0.032*** | -0.030*** |
| | (0.002) | (0.002) | (0.002) | (0.002) |
| COVID-19 Risk Score | -0.001*** | -0.000*** | -0.001*** | -0.001*** |
| | (0.000) | (0.000) | (0.000) | (0.000) |
| Missing Risk Score | -0.017*** | -0.016*** | -0.019*** | -0.019*** |
| | (0.002) | (0.002) | (0.002) | (0.002) |
| $N$ | 90662 | 71839 | 90662 | 71839 |
| $R^2$ | 0.011 | 0.010 | 0.023 | 0.022 |

Robust standard errors are reported in parentheses. Controls for batch fixed effects are omitted from the regression table.

* $p < 0.05$, ** $p < 0.01$, *** $p < 0.001$

*Notes*: Columns 1 and 3 include all patients in the early analysis sample of the second RCT. Columns 2 and 4 exclude patients who were in the *Holdout* arm of the first RCT. Text Message is a dummy variable coded as one when participants received one of the text messages in the second RCT.

Table 16: Effects of Different Types of Second Text Messages on Appointments and Vaccinations at UCLA Health

| | Appointment at UCLA (Within Six Days) | | | | Vaccinated at UCLA (Within Four Weeks) | | | |
|---|---|---|---|---|---|---|---|---|
| | (1) | (2) | (3) | (4) | (5) | (6) | (7) | (8) |
| Self | 0.012*** | | | | 0.006** | | | |
| | (0.002) | | | | (0.002) | | | |
| Prosocial | | 0.013*** | | | | 0.008*** | | |
| | | (0.002) | | | | (0.002) | | |
| Early Access | | | 0.015*** | | | | 0.009*** | |
| | | | (0.002) | | | | (0.002) | |
| Fresh Start | | | | 0.010*** | | | | 0.005* |
| | | | | (0.002) | | | | (0.002) |
| Gender-Female | -0.002 | -0.002 | -0.002 | -0.001 | -0.007*** | -0.004* | -0.005* | -0.005* |
| | (0.002) | (0.002) | (0.002) | (0.002) | (0.002) | (0.002) | (0.002) | (0.002) |
| Gender-Other | -0.009* | -0.005 | -0.004 | -0.002 | -0.011* | -0.006 | -0.004 | -0.003 |
| | (0.004) | (0.005) | (0.005) | (0.004) | (0.005) | (0.006) | (0.007) | (0.005) |
| Age | -0.000** | -0.000*** | -0.000*** | -0.000 | -0.000** | -0.000* | -0.001*** | -0.000* |
| | (0.000) | (0.000) | (0.000) | (0.000) | (0.000) | (0.000) | (0.000) | (0.000) |
| Hispanic | 0.014*** | 0.016*** | 0.016*** | 0.016*** | 0.025*** | 0.027*** | 0.024*** | 0.026*** |
| | (0.004) | (0.004) | (0.004) | (0.004) | (0.005) | (0.005) | (0.005) | (0.005) |
| Non-Hispanic Black | 0.018*** | 0.017*** | 0.021*** | 0.016** | 0.043*** | 0.042*** | 0.042*** | 0.040*** |
| | (0.005) | (0.005) | (0.005) | (0.005) | (0.006) | (0.006) | (0.007) | (0.007) |
| Non-Hispanic Asian | 0.004 | 0.006 | 0.012** | 0.001 | 0.008* | 0.009* | 0.012** | 0.009* |
| | (0.003) | (0.003) | (0.004) | (0.003) | (0.004) | (0.004) | (0.004) | (0.004) |
| Ethnicity-Other | -0.002 | 0.005 | 0.001 | 0.001 | 0.004 | 0.010* | 0.006 | 0.009* |
| | (0.003) | (0.003) | (0.004) | (0.003) | (0.004) | (0.004) | (0.004) | (0.004) |
| Ethnicity-Unknown | -0.010*** | -0.009*** | -0.009*** | -0.008*** | -0.015*** | -0.014*** | -0.014*** | -0.014*** |
| | (0.002) | (0.002) | (0.002) | (0.002) | (0.002) | (0.002) | (0.003) | (0.002) |
| Preferred Spanish | 0.002 | 0.012* | 0.005 | 0.008 | 0.007 | 0.021** | 0.022** | 0.012 |
| | (0.005) | (0.005) | (0.006) | (0.006) | (0.006) | (0.007) | (0.008) | (0.007) |
| Social Vulnerability | -0.000*** | -0.000*** | -0.000*** | -0.000*** | -0.000 | -0.000 | -0.000 | -0.000 |
| Index (SVI) | (0.000) | (0.000) | (0.000) | (0.000) | (0.000) | (0.000) | (0.000) | (0.000) |
| Missing SVI | -0.021*** | -0.021*** | -0.023*** | -0.020*** | -0.033*** | -0.032*** | -0.033*** | -0.033*** |
| | (0.002) | (0.002) | (0.002) | (0.002) | (0.002) | (0.002) | (0.003) | (0.002) |
| COVID-19 Risk Score | -0.000*** | -0.000*** | -0.000** | -0.000*** | -0.000*** | -0.000** | -0.000* | -0.000** |
| | (0.000) | (0.000) | (0.000) | (0.000) | (0.000) | (0.000) | (0.000) | (0.000) |
| Missing Risk Score | -0.026*** | -0.026*** | -0.024*** | -0.022*** | -0.039*** | -0.040*** | -0.039*** | -0.037*** |
| | (0.002) | (0.002) | (0.002) | (0.002) | (0.002) | (0.002) | (0.002) | (0.002) |
| $N$ | 51834 | 51792 | 38862 | 38842 | 51834 | 51792 | 38862 | 38842 |
| $R^2$ | 0.009 | 0.009 | 0.010 | 0.008 | 0.015 | 0.015 | 0.015 | 0.014 |
| Conditions | Self: | Prosocial: | Early Access: | Fresh Start: | Self: | Prosocial: | Early Access: | Fresh Start: |
| Included | Simple | Simple | Self | Self | Simple | Simple | Self | Self |
| | Early Access | Early Access | Prosocial | Prosocial | Early Access | Early Access | Prosocial | Prosocial |
| | Fresh Start | Fresh Start | | | Fresh Start | Fresh Start | | |
| | Holdout | Holdout | Holdout | Holdout | Holdout | Holdout | Holdout | Holdout |

Robust standard errors are reported in parentheses. Controls for batch fixed effects are omitted from the regression table.

* $p < 0.05$, ** $p < 0.01$, *** $p < 0.001$

*Notes*: Self is a dummy variable coded as one when participants received one of the text messages highlighting personal benefits. Prosocial is a dummy variable coded as one when participants received one of the text messages highlighting prosocial benefits. Early Access is a dummy variable coded as one when participants received one of the text messages highlighting their early access to the vaccine. Fresh Start is a dummy variable coded as one when participants received one of the text messages framing getting vaccinated as an opportunity for a fresh start.

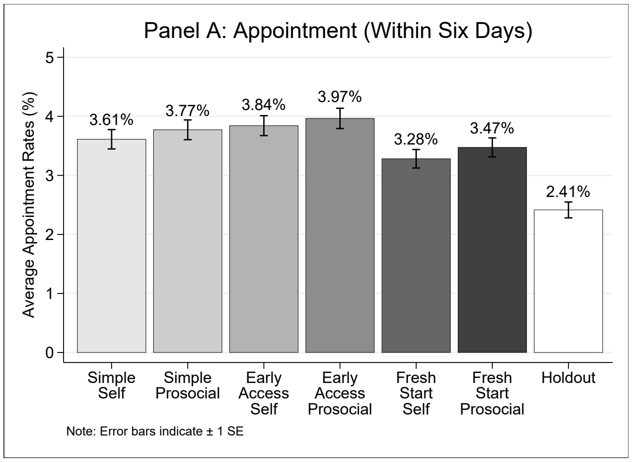


(a) Panel A

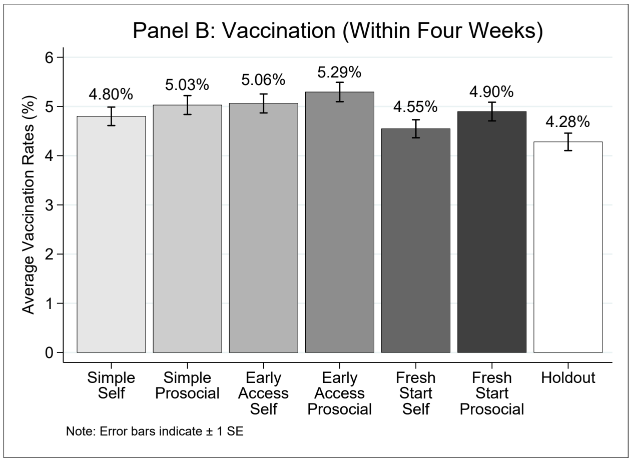


(b) Panel B

Figure 3: Appointment and Vaccination Rates by Condition (Second RCT)

#### 1.5 Additional Information and Robustness Checks

In this section, we present additional information about patients' vaccination behavior that may be of interest to policymakers and the public. We also report regression tables for three robustness checks: (1) assessing whether patients scheduled the first-dose appointment anytime after 3pm on the first (second) reminder date by the time of our data extraction (Tables 17-18); (2) excluding control variables (Tables 19-20); and (3) conducting logistic regressions (Tables 21-22).

Among patients in our first RCT sample who scheduled the first-dose appointment at UCLA Health within six days of the first reminder date (n=11,515), 88.05% either had obtained their first dose or had an active, upcoming appointment as of March 26, 2021. That is, only 11.95% patients cancelled their appointments without rescheduling a new one by March 26, 2021. Among the subset of patients who had an active first-dose appointment that was supposed to take place before March 26, 2021 (n=8263), 99.90% completed it; only eight patients did not show up for their active first-dose appointments, indicating a 0.10% no-show rate. The low no-show rate may occur because UCLA Health exerted extra effort to remind people of their vaccination appointment or because patients who scheduled an appointment were determined to receive the vaccine. The relatively low cancellation rate and the extremely low no-show rate suggest that the biggest challenge to tackle for the vaccine uptake challenge is to get people to schedule an appointment.

Among people who were in the early analysis sample of either RCT and obtained the first dose of Moderna or Pfizer at UCLA Health as of our data extraction on March 26, 2021, 92.61% had an appointment for the second dose or already obtained the second dose as of March 26, 2021.[10] UCLA Health took great measures to ensure that patients schedule their second-dose appointment right after they obtain the first dose, which helps combat barriers patients may otherwise face (e.g., forgetfulness, procrastination) if they are left to schedule the second-dose appointment later online. Also, the high appointment rate for the second dose may also be driven by the strong intentions to receive full vaccination among those who cared enough to get the first dose.

[10]Since we do not have data about appointments made outside of UCLA Health, we could only report statistics based on UCLA Health records.

Table 17: Effects of Receiving a Text Message and Ownership Language on Appointments Observed in a Longer Time Window

| | Appointment at UCLA Any Time After the First Reminder Date | |
|---|---|---|
| | (1) | (2) |
| Text Message | 0.030*** | |
| | (0.003) | |
| Ownership | | 0.009*** |
| | | (0.003) |
| Video | | -0.002 |
| | | (0.003) |
| Gender-Female | -0.003 | -0.005* |
| | (0.002) | (0.003) |
| Gender-Other | -0.023 | -0.034 |
| | (0.034) | (0.044) |
| Age | -0.004*** | -0.004*** |
| | (0.000) | (0.000) |
| Hispanic | 0.040*** | 0.039*** |
| | (0.005) | (0.006) |
| Non-Hispanic Black | 0.108*** | 0.110*** |
| | (0.006) | (0.007) |
| Non-Hispanic Asian | 0.018*** | 0.019*** |
| | (0.004) | (0.005) |
| Ethnicity-Other | -0.002 | -0.005 |
| | (0.005) | (0.005) |
| Ethnicity-Unknown | -0.051*** | -0.051*** |
| | (0.003) | (0.003) |
| Preferred Spanish | 0.035*** | 0.033*** |
| | (0.007) | (0.008) |
| Social Vulnerability Index (SVI) | -0.001*** | -0.001*** |
| | (0.000) | (0.000) |
| Missing SVI | -0.101*** | -0.104*** |
| | (0.003) | (0.004) |
| COVID-19 Risk Score | -0.000 | -0.000 |
| | (0.000) | (0.000) |
| Missing Risk Score | -0.084*** | -0.086*** |
| | (0.003) | (0.004) |
| $N$ | 113229 | 90550 |
| $R^2$ | 0.066 | 0.065 |
| Conditions Included | All | Conditions within the Text Message arm |

Robust standard errors are reported in parentheses. Controls for batch fixed effects are omitted from the regression table.
* $p < 0.05$, ** $p < 0.01$, *** $p < 0.001$

*Notes*: Text Message is a dummy variable coded as one when participants received one of the text messages in the second RCT.

Table 18: Effects of Receiving the Second Text Message on Appointments Observed in a Longer Time Window

| | Appointment at UCLA Any Time After the Second Reminder Date | |
|---|---|---|
| | All Patients | Excl. Patients in the Holdout Arm of the First RCT |
| | (1) | (2) |
| Text Message | 0.007** | 0.008** |
| | (0.003) | (0.003) |
| Gender-Female | 0.004* | 0.003 |
| | (0.002) | (0.002) |
| Gender-Other | -0.024*** | -0.026*** |
| | (0.006) | (0.007) |
| Age | -0.002*** | -0.002*** |
| | (0.000) | (0.000) |
| Hispanic | 0.030*** | 0.026*** |
| | (0.004) | (0.005) |
| Non-Hispanic Black | 0.069*** | 0.067*** |
| | (0.006) | (0.006) |
| Non-Hispanic Asian | 0.012*** | 0.014*** |
| | (0.004) | (0.004) |
| Ethnicity-Other | 0.012** | 0.009* |
| | (0.004) | (0.004) |
| Ethnicity-Unknown | -0.017*** | -0.017*** |
| | (0.002) | (0.002) |
| Preferred Spanish | 0.026*** | 0.026*** |
| | (0.006) | (0.007) |
| Social Vulnerability Index (SVI) | -0.001*** | -0.000*** |
| | (0.000) | (0.000) |
| Missing SVI | -0.051*** | -0.049*** |
| | (0.002) | (0.003) |
| COVID-19 Risk Score | -0.001*** | -0.001*** |
| | (0.000) | (0.000) |
| Missing Risk Score | -0.033*** | -0.033*** |
| | (0.002) | (0.003) |
| *N* | 90662 | 71839 |
| $R^2$ | 0.038 | 0.036 |

Robust standard errors are reported in parentheses. Controls for batch fixed effects are omitted from the regression table.
$^{*}$ $p < 0.05$, $^{**}$ $p < 0.01$, $^{***}$ $p < 0.001$

*Notes*: Column 1 includes all patients in the early analysis sample of the second RCT. Column 2 excludes patients who were in the *Holdout* arm of the first RCT. Text Message is a dummy variable coded as one when participants received one of the text messages in the second RCT. Ownership is a dummy variable coded as one when participants received one of the messages containing the ownership language in the first RCT. Video is a dummy variable coded as one when participants received one of the messages containing a link to the video in the first RCT.

Table 19: Effects of Receiving a Text Message and Ownership Language (Without Controls)

| | Appointment at UCLA (Within Six Days) | | Vaccinated at UCLA (Within Four Weeks) | |
|---|---|---|---|---|
| | (1) | (2) | (3) | (4) |
| Text Message | 0.052*** | | 0.030*** | |
| | (0.002) | | (0.002) | |
| Ownership | | 0.013*** | | 0.009*** |
| | | (0.002) | | (0.002) |
| Video | | -0.002 | | -0.001 |
| | | (0.002) | | (0.002) |
| $N$ | 113229 | 90550 | 113229 | 90550 |
| $R^2$ | 0.005 | 0.000 | 0.001 | 0.000 |
| Conditions Included | All | Conditions within the Text Message arm | All | Conditions within the Text Message arm |

Robust standard errors are reported in parentheses. Controls for batch fixed effects are omitted from the regression table.
$^{*}$ $p < 0.05$, $^{**}$ $p < 0.01$, $^{***}$ $p < 0.001$

*Notes*: Text Message is a dummy variable coded as one when participants received one of the text messages in the first RCT. Ownership is a dummy variable coded as one when participants received one of the messages containing the ownership language in the first RCT. Video is a dummy variable coded as one when participants received one of the messages containing a link to the video in the first RCT.

Table 20: Effects of Receiving the Second Text Message (Without Controls)

| | Appointment at UCLA (Within Six Days) | | Vaccinated at UCLA (Within Four Weeks) | |
|---|---|---|---|---|
| | All Patients | Excl. Patients in the Holdout Arm of the first RCT | All Patients | Excl. Patients in the Holdout Arm of the first RCT |
| | (1) | (2) | (3) | (4) |
| Text Message | 0.012*** | 0.010*** | 0.007*** | 0.006** |
| | (0.002) | (0.002) | (0.002) | (0.002) |
| $N$ | 90662 | 71839 | 90662 | 71839 |
| $R^2$ | 0.001 | 0.000 | 0.000 | 0.000 |

Robust standard errors are reported in parentheses. Controls for batch fixed effects are omitted from the regression table.
$^{*}$ $p < 0.05$, $^{**}$ $p < 0.01$, $^{***}$ $p < 0.001$

*Notes*: Columns 1 and 3 include all patients in the early analysis sample of the second RCT. Columns 2 and 4 exclude patients who were in the *Holdout* arm of the first RCT. Text Message is a dummy variable coded as one when participants received one of the text messages in the second RCT.

Table 21: Effects of Receiving a Text Message and Ownership Language (Logistic Regressions)

| | Appointment at UCLA (Within Six Days) | | Vaccinated at UCLA (Within Four Weeks) | |
|---|---|---|---|---|
| | (1) | (2) | (3) | (4) |
| Text Message | 0.698*** | | 0.282*** | |
| | (0.030) | | (0.024) | |
| Ownership | | 0.131*** | | 0.074*** |
| | | (0.022) | | (0.020) |
| Video | | -0.021 | | -0.010 |
| | | (0.022) | | (0.020) |
| Gender-Female | -0.064** | -0.074*** | -0.109*** | -0.113*** |
| | (0.020) | (0.022) | (0.018) | (0.020) |
| Gender-Other | -0.123 | -0.039 | -0.250 | -0.113 |
| | (1.029) | (1.035) | (1.027) | (1.035) |
| Age | -0.025*** | -0.026*** | -0.025*** | -0.026*** |
| | (0.001) | (0.001) | (0.001) | (0.001) |
| Hispanic | 0.133*** | 0.131** | 0.234*** | 0.232*** |
| | (0.038) | (0.041) | (0.033) | (0.037) |
| Non-Hispanic Black | 0.407*** | 0.424*** | 0.510*** | 0.520*** |
| | (0.042) | (0.045) | (0.037) | (0.041) |
| Non-Hispanic Asian | 0.098** | 0.084* | 0.128*** | 0.122*** |
| | (0.036) | (0.039) | (0.032) | (0.036) |
| Ethnicity-Other | -0.162*** | -0.147*** | -0.102** | -0.108** |
| | (0.042) | (0.044) | (0.037) | (0.040) |
| Ethnicity-Unknown | -0.500*** | -0.460*** | -0.523*** | -0.523*** |
| | (0.038) | (0.040) | (0.034) | (0.037) |
| Preferred Spanish | 0.121* | 0.102 | 0.200*** | 0.199*** |
| | (0.056) | (0.061) | (0.048) | (0.053) |
| Social Vulnerability Index (SVI) | -0.007*** | -0.007*** | -0.006*** | -0.006*** |
| | (0.000) | (0.000) | (0.000) | (0.000) |
| Missing SVI | -0.809*** | -0.816*** | -1.025*** | -1.026*** |
| | (0.055) | (0.058) | (0.052) | (0.057) |
| COVID-19 Risk Score | 0.001*** | 0.001*** | 0.001*** | 0.001*** |
| | (0.000) | (0.000) | (0.000) | (0.000) |
| Missing Risk Score | -0.784*** | -0.763*** | -0.839*** | -0.824*** |
| | (0.043) | (0.045) | (0.039) | (0.043) |
| *N* | 113229 | 90550 | 113229 | 90550 |
| Conditions Included | All | Conditions within the Text Message arm | All | Conditions within the Text Message arm |

Controls for batch fixed effects are omitted from the regression table.

* $p < 0.05$, ** $p < 0.01$, *** $p < 0.001$

*Notes*: Text Message is a dummy variable coded as one when participants received one of the text messages in the first RCT. Ownership is a dummy variable coded as one when participants received one of the messages containing the ownership language in the first RCT. Video is a dummy variable coded as one when participants received one of the messages containing a link to the video in the first RCT.

Table 22: Effects of Receiving the Second Text Message (Logistic Regressions)

| | Appointment at UCLA (Within Six Days) | | Vaccinated at UCLA (Within Four Weeks) | |
|---|---|---|---|---|
| | All Patients | Excl. Patients in the Holdout Arm of the first RCT | All Patients | Excl. Patients in the Holdout Arm of the first RCT |
| | (1) | (2) | (3) | (4) |
| Text Message | 0.436*** | 0.386*** | 0.156*** | 0.156** |
| | (0.061) | (0.070) | (0.047) | (0.054) |
| Gender-Female | -0.032 | -0.018 | -0.067* | -0.069 |
| | (0.037) | (0.043) | (0.032) | (0.037) |
| Gender-Other | 0.000 | 0.000 | 0.000 | 0.000 |
| | (.) | (.) | (.) | (.) |
| Age | -0.018*** | -0.017*** | -0.020*** | -0.018*** |
| | (0.002) | (0.003) | (0.002) | (0.002) |
| Hispanic | 0.320*** | 0.255** | 0.372*** | 0.376*** |
| | (0.067) | (0.079) | (0.057) | (0.065) |
| Non-Hispanic Black | 0.418*** | 0.361*** | 0.593*** | 0.614*** |
| | (0.078) | (0.092) | (0.062) | (0.071) |
| Non-Hispanic Asian | 0.153* | 0.153* | 0.206*** | 0.231*** |
| | (0.067) | (0.077) | (0.057) | (0.065) |
| Ethnicity-Other | 0.031 | -0.005 | 0.110 | 0.120 |
| | (0.073) | (0.085) | (0.061) | (0.070) |
| Ethnicity-Unknown | -0.326*** | -0.403*** | -0.348*** | -0.399*** |
| | (0.066) | (0.079) | (0.059) | (0.070) |
| Preferred Spanish | 0.216* | 0.249* | 0.226** | 0.269** |
| | (0.091) | (0.106) | (0.075) | (0.086) |
| Social Vulnerability Index (SVI) | -0.008*** | -0.007*** | -0.006*** | -0.007*** |
| | (0.001) | (0.001) | (0.001) | (0.001) |
| Missing SVI | -1.010*** | -0.998*** | -1.217*** | -1.182*** |
| | (0.109) | (0.126) | (0.102) | (0.115) |
| COVID-19 Risk Score | -0.009*** | -0.009*** | -0.012*** | -0.012*** |
| | (0.001) | (0.001) | (0.001) | (0.001) |
| Missing Risk Score | -0.643*** | -0.643*** | -0.643*** | -0.659*** |
| | (0.079) | (0.092) | (0.075) | (0.086) |
| *N* | 90637 | 71820 | 90637 | 71820 |

Controls for batch fixed effects are omitted from the regression table.

* $p < 0.05$, ** $p < 0.01$, *** $p < 0.001$

*Notes*: Columns 1 and 3 include all patients in the early analysis sample of the second RCT. Columns 2 and 4 exclude patients who were in the *Holdout* arm of the first RCT. Text Message is a dummy variable coded as one when participants received one of the text messages in the second RCT.

### 2 Online Experiments Assessing Intentions and Mechanisms

We conducted two pre-registered online experiments in February 2021 with three objectives. First, using a different sample of respondents than patients at UCLA Health, we sought to examine how the text messages from the first RCT affect people's reported likelihood of scheduling the COVID-19 vaccine and how people perceive these messages' persuasiveness. Second, we wanted to check if the ownership language tested in the first RCT indeed creates a sense of psychological ownership of the vaccine, as we intended. Third, we explored whether the video shifts certain beliefs and perceptions about COVID-19 and the COVID-19 vaccine. We pre-registered the experiments on aspredicted.org (`https://aspredicted.org/blind.php?x=u2ng5c` and `https://aspredicted.org/blind.php?x=ae3ci5`). The two experiments had the same design and analysis plan but the second experiment was a replication and had a different recruitment method. The key results are similar between the two experiments (see Tables 33-??), so we combined the data, which gave us a larger sample for analyzing heterogeneous treatment effects.

#### 2.1 Method

##### 2.1.1 Participants

In the first online experiment, we aimed to recruit 1,200 participants who fit our pre-registered selection criteria. People on Amazon's Mechanical Turk (MTurk) and Prolific Academic (Prolific) who passed a Captcha and an attention check question, had not already received the COVID-19 vaccine or did not already have a COVID-19 vaccination appointment, and indicated that they were either Republicans or Democrats were eligible to take our study.[11] Since the majority of participants on MTurk and Prolific identify with the Democratic Party based on our prior data collection experiences and prior research (Levay et al., 2016; Pedersen and Favero, 2020), we had to take extra measures to obtain a more balanced number of Republicans and Democrats as study participants in order to conduct heterogeneity analyses by political affiliation. We were interested in this analysis because recent national surveys suggest Republicans are more hesitant toward the vaccine than Democrats (Funk and Tyson, 2020; Hamel et al., 2020; Ruiz and Bell, 2021).[12] When collecting data on MTurk, we used preset quotas in our survey on Qualtrics such that once the number of Democrats hit 600, only people who identified with Republicans could take the study. When we later collected data on Prolific, we used Prolific's screening tools to separately target Republicans and Democrats. Our sample for the first experiment consists of 1,163 participants (51.59% female, M_age = 39.64) who satisfied the aforementioned criteria, at least completed the portion of survey involving our pre-registered primary dependent variables, did not report having technical issues with watching the video, and did not

[11] We first asked participants several screening questions. Only those who satisfied the criteria were allowed to take our study; those who were not eligible for the study were offered a payment of $0.10 as a compensation for their time. Possibly because of the unusual nature of our recruitment method, our data collection on MTurk became very slow after we hit about 500 Democrats and 450 Republicans, at which point we switched to Prolific to speed up data collection. This is a minor deviation from our pre-registration. On Prolific, we initially used the same recruitment method as on MTurk but later were informed that we could not kick people out of a study on Prolific even if we compensated them for the time they spent taking a few screening questions. Thus, for about 40 participants recruited in the end of the first experiment (as well as all participants taking our second experiment on Prolific), we did not kick people out in the survey but only kept those who satisfied our criteria in the analysis.

[12] The survey we conducted to inform our design of the video also revealed that Republicans had significantly lower self-reported intentions to take the COVID-19 vaccine, compared to Democrats. See Section 3.

report having taken a similar study on MTurk (only relevant to the Prolific sample).[13]

In the second online experiment, we aimed to recruit 800 participants from Prolific who fit our pre-registered selection criteria: people who passed an attention check question, had not already received the COVID-19 vaccine or did not already have a COVID-19 vaccination appointment, did not report having taken a similar study on MTurk, and had no technical problems with watching our COVID-19 video. We again used Prolific's screening tools and sought to recruit 400 Republicans and 400 Democrats. Prolific's screening tools are not perfect as some people marked by Prolific as Democrats (Republicans) identified themselves as Republicans (Democrats) or neither Republicans or Democrats when responding to our survey. Our sample for the second experiment consists of 840 participants (50.12% female, M_age = 35.46) who satisfied the aforementioned criteria and at least completed the portion of survey involving our pre-registered primary dependent variables.

Altogether, across the two online experiments, our final sample consists of 2,003 participants (50.97% female, M_age = 37.89). If a person took the survey and completed our pre-registered dependent variables more than once, we kept their first response. Participants received $0.90 on MTurk or $1.10 on Prolific for completing our 6-minute survey.

###### 2.1.2 Procedure

Upon passing the screening, we instructed participants to imagine receiving a text message from their healthcare provider encouraging them to get the COVID-19 vaccine. We randomly assigned participants to one of four conditions: *Simple Text*, *Ownership Text*, *Simple Text with Video*, or *Ownership Text with Video* condition. Participants in each condition were presented with a text message that was identical to the message used in the corresponding condition of our first RCT, except that we used placeholders for patient first name, video link, and the link to the vaccination appointment website and we replaced "UCLA Health" with "our clinics". Specifically,

- In the *Simple Text* condition, participants saw the following message: "[Your first name], you can get the COVID-19 vaccine at our clinics. Make a vaccination appointment here: [link to the appointment website]".
- In the *Ownership Text* condition, the message read, "[Your first name], a COVID-19 vaccine has just been made available to you at our clinics. Claim your dose today by making a vaccination appointment here: [link to the appointment website]".
- In the *Simple Text with Video* condition, the message read, "[Your first name], you can get the COVID-19 vaccine at our clinics. Please watch this important 2 min video: [link to the video]. Make a vaccination appointment here: [link to the appointment website]".
- In the *Ownership Text with Video* condition, the message read, "[Your first name], a COVID-19 vaccine has just been made available to you at our clinics. Please take 2 simple steps: 1. Watch this important

[13] We did not pre-register dropping people who had technical problems with the video or had taken a similar study on MTurk in the first experiment, but we realized these issues later and added these exclusion criteria when pre-registering the second experiment. Here, we drop these people from the first experiment for consistency but keeping these observations does not qualitatively change the results.

2 min video: [link to the video]. 2. Claim your dose today by making a vaccination appointment here: [link to the appointment websites]".

In the two video conditions, we then asked participants to watch the 2-minute video we sent to patients in the corresponding conditions of the first RCT. As stated above, we designed the video to influence people's beliefs about the prevalence and ease of transmission of COVID-19, educate them about the effectiveness and safety of the COVID-19 vaccine, and recommend them to get the vaccine. The full survey instructions are available at `https://osf.io/qn8hr/?view_only=cf7b2bc590054aee8c4a2bae99ef20c5`.

##### 2.1.3 Measures

After people read the text message presented to them and watched the video (if they were in one of the video conditions), we collected a number of measures.

**Primary Dependent Measures.** Our first primary dependent measure was participants' self-reported likelihood of scheduling a COVID-19 vaccination appointment upon receiving the message (*Scheduling Likelihood*): "How likely would you be to schedule a vaccination appointment after receiving this message from your healthcare provider?" (from 1 = Not at all Likely to 7 = Very Likely). Our second dependent measure was participants' perceived persuasiveness of the text message (*Persuasiveness*): "How persuasive do you think the message is?" (from 1 = Not at all persuasive to 7 = Very persuasive).

**Mechanism Measures for the Ownership Text.** To test whether the language in the messages containing ownership text created a sense of endowment toward the vaccine, we measured *Psychological Ownership*, "To what extent does the text message make you feel that the COVID-19 vaccine is already yours?" (from 1 = Not at all to 7 = Very much). To address the possibility that our messages containing ownership text are longer and thus could make getting the COVID-19 vaccine seem more difficult, we measured *Easiness*, "To what extent does the text message make you feel that getting the COVID-19 vaccine is easy?" (from 1 = Not at all to 7 = Very much).

**Mechanism Measures for the Video.** To test whether the video affected participants' beliefs about the prevalence of COVID-19 and the effectiveness of the COVID-19 vaccine as well as additional perceptions about COVID-19 and the vaccine, we collected the following measures:

- *Infection Likelihood without Vaccine*: We asked participants to report their likelihood of getting infected with COVID-19 (including cases with and without symptoms) if they did not get the COVID-19 vaccine (from 0% = I certainly won't get infected with COVID-19 to 100% = I certainly will get infected with COVID-19). We also asked participants to report their likelihood of developing symptomatic COVID-19 if they did not get the COVID-19 vaccine (from 0% = I certainly won't develop it to 100% = I certainly will develop it). Responses to those questions were highly correlated ($r = 0.82$, $p < 0.0001$) and thus were averaged to form a measure of *Infection Likelihood without Vaccine*.

- *Infection Likelihood with Vaccine*: Participants also separately rated their likelihood of getting infected with COVID-19 or developing symptomatic COVID-19 if they got the COVID-19 vaccine, using the same 0%-100% scales as described above.[14] Responses to those questions were highly correlated ($r = 0.71$, $p < 0.0001$) and were averaged to form a measure of *Infection Likelihood with Vaccine*.

[14]The two questions about COVID-19 infections and the two questions about symptomatic COVID-19 were counterbalanced. The results reported later about people's beliefs in infection likelihood and effectiveness are robust if we separately examine people's beliefs about general COVID-19 infections versus symptomatic COVID-19.

- *Vaccine Effectiveness*: For each participant, we subtracted *Infection Likelihood with Vaccine* from *Infection Likelihood without Vaccine* to capture her belief in the effectiveness of the COVID-19 vaccine. The value of this variable in theory could range from -100% to 100%, and a positive value indicates that participants believed getting the vaccine could reduce their infection likelihood. In our analysis, for this and the aforementioned two measures about infection likelihood, we multiplied the raw data by 100.

- *Worry of Transmission*: Participants indicated the extent to which they worried about passing COVID-19 to other people (from 1 = Not at all to 7 = Very much).

- *Anticipated Regret*: Participants indicated how much they would regret not getting the COVID-19 vaccine if, when eligible, they decided not to get it and ended up getting symptomatic COVID-19 (from 1 = Not at all to 7 = Very much).

- *Trust in Vaccine*: We measured trust in the safety of the vaccine and the vaccine development process with five questions on a seven-point scale (from 1 = Not at all to 7 = Very Much). The questions assessed worries about side effects ("To what extent are you concerned about potential side effects of the COVID-19 vaccine?"), concerns about potential negative long-term impacts of the vaccine ("To what extent do you worry that the COVID-19 vaccine will have negative long-term impacts on your health?"), concerns about the speed of the vaccine authorization process ("To what extent are you concerned that the current COVID-19 vaccines were authorized for use too quickly?"), overall trust in the development of the vaccine ("How much do you trust the research and development process of COVID-19 vaccines?"), and perceived safety of the vaccine ("How confident are you that the research and development process of COVID-19 vaccine has produced a safe vaccine?"). Participants' responses to these five questions reached a high inter-item reliability (Cronbach's alpha = 0.93); thus, we averaged responses to these questions (after reverse coding the first three questions) to form a composite score of *Trust in Vaccine*. The higher one's composite score is, the more they trusted the safety of vaccine and its development process.

**Additional Measures.** In the conditions containing the video, we also measured the perceived persuasiveness ("How persuasive do you think the video is?") and the perceived accuracy of the video ("How accurate do you think the video is?") (from a 1 = Not at all to 7 = Very much). Finally, participants reported whether they got the flu shot during each of the two recent flu seasons (2019-2020 and 2020-2021 seasons), the frequency with which they washed their hands when coming back from being outside and the frequency with which they wore a mask when they could not maintain social distance outside (from 1 = Never or almost never to 5 = Always or almost always), whether they ever tested positive for COVID-19 (or thought they had COVID-19), their demographics and socio-economics status (age, gender, race/ethnicity, highest level of education attained, pre-tax household income in 2020, whether they had children under 18[15], whether they were living with anyone in the same household, and which state they were living in). Participants in the video conditions additionally reported whether they had any technical issue with the video. Participants recruited from Prolific were also asked whether they took a similar study on MTurk.

[15]This question was added after our first day of data collection because we learned from a colleague's presentation that people with children reported lower COVID-19 vaccination intentions than those without children. We decided to add this for exploratory heterogeneity analyses.

#### 2.2 Results

As pre-registered, we used ordinary least squares (OLS) regressions with robust standard errors to predict dependent variables. When we analyze the main effect of ownership language, the key predictor is a binary variable indicating whether or not participants read one of the text messages containing the ownership language (*Ownership*). It equals one for participants in the *Ownership Text* or *Ownership Text with Video* condition and zero otherwise. When we analyze the main effect of our educational video, the key predictor is a binary variable indicating whether or not participants read one of the text messages containing the video (*Video*). It equals one for participants in the *Simple Text with Video* or *Ownership Text with Video* condition and zero otherwise. When we analyze the simple effect of ownership language (when the message does not contain the video), the simple effect of our educational video (when the message does not contain ownership language), and the interaction between these two interventions, our key predictors include the two binary indicators (Ownership and Video) as well as their interaction term. All regressions control for participants' gender (indicators for female and other/unknown, with male as the reference group), age, and race/ethnicity (indicators for non-Hispanic Black, non-Hispanic Asian, Hispanic, other/mixed race/unknown, with non-Hispanic White as the reference group), as well as an indicator for whether participants did not respond to demographic questions.[16]

##### 2.2.1 Scheduling Likelihood

Figure 4 Panel A displays the mean rating of scheduling likelihood for each message based on raw data. Table 23 Columns 1-2 report regression results about scheduling likelihood. Column 1 indicates that adding ownership language did not have a positive main effect. Instead, it shows a positive main effect of adding the video to the text message: people overall indicated a greater likelihood of scheduling a vaccination appointment after receiving a message containing the video than after receiving a message without the video (B = 0.25, SE = 0.10, p = 0.01).

Furthermore, as shown in Column 2, we observed a positive simple effect of video whereby the *Simple Text with Video* message significantly increased people's self-reported likelihood of scheduling a vaccination appointment, relative to the *Simple Text* message (B = 0.40, SE = 0.13, p = 0.003). There is not a simple effect of ownership text (B = -0.002, SE = 0.14, p = 0.99), neither is there a significant interaction between the two interventions (B = -0.30, SE = 0.19, p = 0.13). Specifically, Wald tests suggest that participants' likelihood of scheduling an appointment did not significantly differ between the *Ownership Text with Video* message and the *Simple Text* message (difference = 0.10, SE = 0.14, p = 0.47) or the *Ownership Text* message (difference = 0.10, SE = 0.14, p = 0.48); but it was lower in the *Ownership Text with Video* condition than in the *Simple Text with Video* condition (difference = -0.30, SE = 0.13, p = 0.03).

##### 2.2.2 Message Persuasiveness

Figure 4 Panel B displays the mean persuasiveness rating for each message based on raw data. Table 23 Columns 3-4 report regression results about *Persuasiveness.* Column 3 shows a positive main effect of adding ownership language: people rated the two messages containing ownership text as significantly more

[16] When pre-registering the study, we did not consider the possibility that some participants (n = 21) responded to our primary dependent variables but did not provide demographics. We treated those people's gender and race/ethnicity as unknown and assigned the average value of age as their age, and added a control for whether people missed reporting demographics.

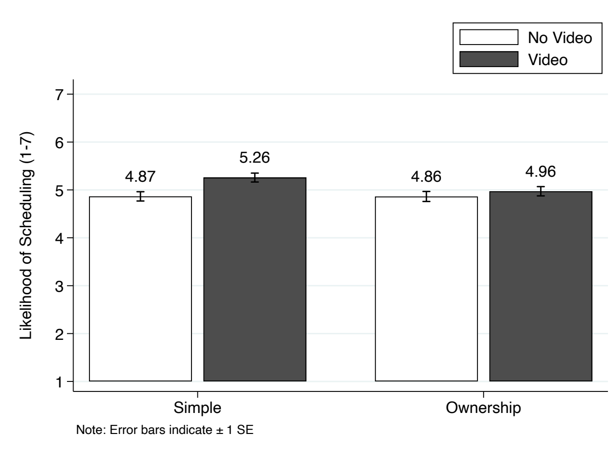


(a) Panel A

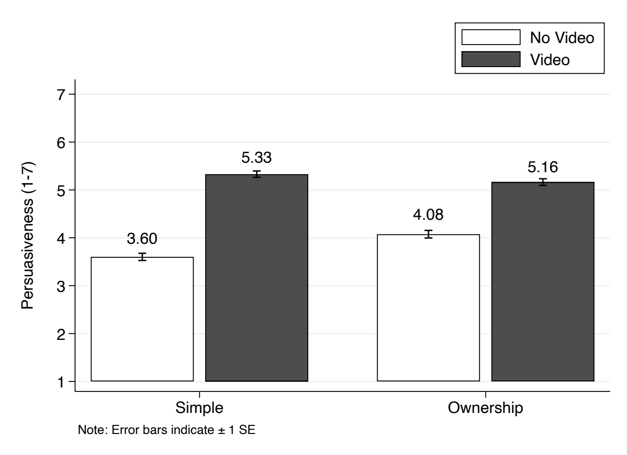


(b) Panel B

Figure 4: Likelihood of Scheduling and Persuasiveness by Message

persuasive than the other two messages (B = 0.18, SE = 0.08, p = 0.02). It also shows a positive main effect of adding the video to the text message: people rated the two messages containing the video as significantly more persuasive than the two messages that do not contain the video (B = 1.42, SE = 0.07, p < 0.001).

Furthermore, as shown in Column 4, we observed a positive simple effect of ownership text whereby the *Ownership Text* message was viewed as more persuasive than the *Simple Text* message (B = 0.47, SE = 0.11, p < 0.001), and a positive simple effect of video whereby the *Simple Text with Video* message was perceived as more persuasive than the *Simple Text* message (B = 1.72, SE = 0.01, p < 0.001). We also observed a significant, negative interaction between the *Video* indicator and the *Ownership Text* indicator (B = -0.63, SE = 0.15, p < 0.001), which suggests that the impact of adding the educational video on persuasiveness and that of adding ownership language are not additive. Specifically, Wald tests suggest that the *Ownership Text with Video* message was more persuasive than both the *Simple Text* message (difference = 1.56, SE = 0.10, p < 0.001) and the *Ownership Text* message (difference = 1.10, SE = 0.10, p < 0.001), and was marginally significantly less persuasive than the *Simple Text with Video* message (difference = -0.16, SE = 0.10, p = 0.09).

##### 2.2.3 Mechanisms for the Ownership Text

Next, we tested how our ownership language changes people's psychological ownership of the vaccine and the perceived ease of getting the vaccine. Table 24 Columns 1-2 report the results of OLS regressions that predict *Psychological Ownership* and *Easiness* as a function of an indicator for whether people read a text messaging containing ownership language and the control variables. Consistent with the intention behind our design of the ownership language, the ownership language elevated participants' feeling that a COVID-19 vaccine was already theirs (B = 0.38, SE = 0.08, p < 0.001; Column 1). Also, the ownership language had virtually no impact on people's perceptions of how easy it would be to receive the COVID-19 vaccine (B = 0.003, SE = 0.07, p = 0.97; Column 2). This helps address the concern that the text message containing ownership language is longer and could increase the perceived complexity of getting vaccinated.

Table 23: Effects of Text Messages on Likelihood of Scheduling and Persuasiveness

| | Scheduling Likelihood | | Persuasiveness | |
|---|---|---|---|---|
| | (1) | (2) | (3) | (4) |
| Ownership | -0.15 | -0.00 | 0.16* | 0.47*** |
| | (0.10) | (0.14) | (0.07) | (0.11) |
| Video | 0.25* | 0.40** | 1.41*** | 1.72*** |
| | (0.10) | (0.13) | (0.07) | (0.10) |
| Ownership x Video | | -0.30 | | -0.63*** |
| | | (0.19) | | (0.15) |
| Gender-Female | -0.01 | -0.01 | 0.17* | 0.17* |
| | (0.10) | (0.10) | (0.07) | (0.07) |
| Gender-Other | 0.92* | 0.92* | 0.38 | 0.38 |
| | (0.41) | (0.41) | (0.39) | (0.39) |
| Age | -0.01 | -0.01 | 0.01** | 0.01** |
| | (0.00) | (0.00) | (0.00) | (0.00) |
| Hispanic | 0.18 | 0.18 | 0.14 | 0.12 |
| | (0.23) | (0.23) | (0.18) | (0.18) |
| Non-Hispanic Black | -0.51** | -0.52** | 0.14 | 0.12 |
| | (0.20) | (0.20) | (0.16) | (0.16) |
| Non-Hispanic Asian | 0.74*** | 0.74*** | 0.02 | 0.01 |
| | (0.13) | (0.13) | (0.12) | (0.12) |
| Ethnicity-Other/Unknown | 0.02 | 0.02 | -0.20 | -0.19 |
| | (0.22) | (0.22) | (0.16) | (0.16) |
| Missing Demographics | -0.87 | -0.87 | 0.07 | 0.07 |
| | (0.66) | (0.66) | (0.57) | (0.56) |
| Constant | 5.16*** | 5.09*** | 3.36*** | 3.22*** |
| | (0.18) | (0.19) | (0.13) | (0.14) |
| $N$ | 2003 | 2003 | 2003 | 2003 |
| $R^2$ | 0.026 | 0.027 | 0.166 | 0.174 |

Robust standard errors are reported in parentheses

* $p < 0.05$, ** $p < 0.01$, *** $p < 0.001$

*Notes*: Video is a dummy variable coded as one when participants received one of the text messages containing the video. Ownership is a dummy variable coded as one when participants received one of the text messages containing the ownership language.

##### 2.2.4 Mechanisms for the Video

Since we are interested in how the educational video may change people's beliefs and perceptions about COVID-19 and the vaccine, we focus on the main effect of the video. Table 25 Columns 1-6 report the results of OLS regressions that predict each belief or perception measure as a function of an indicator for whether people watched the video and the aforementioned control variables. As shown in Column 1, our video significantly increased people's predicted likelihood of getting COVID-19 without the vaccine by 2.79 percentage points (B = 2.79, SE = 1.11, p = 0.01), suggesting that the video made people believe COVID-19 is more prevalent and easier to transmit. As shown in Column 2, *Infection Likelihood with Vaccine* was significantly lower by 2.21 percentage points among people who watched the video than among those who did not watch the video (B = -2.21, SE = 0.83, p = 0.01), suggesting that watching the video led people to believe they would be less likely to get COVID-19 with the vaccine. Furthermore, Column 3 shows that *Vaccine Effectiveness*—which we calculated by subtracting *Infection Likelihood with Vaccine* from *Infection Likelihood without Vaccine*—was significantly higher by 5.01 percentage points in the two conditions involving the video than in the other two conditions (B = 5.01, SE = 1.28, p < 0.001). This suggests that watching

Table 24: Effects of Ownership Language on Psychological Ownership and Ease of Getting Vaccinated

| | Psychological Ownership | Easiness |
|---|---|---|
| | (1) | (2) |
| Ownership | 0.38*** | 0.00 |
| | (0.08) | (0.07) |
| Gender-Female | 0.24** | 0.18** |
| | (0.08) | (0.07) |
| Gender-Other | 0.63 | 0.24 |
| | (0.36) | (0.30) |
| Age | 0.00 | -0.00 |
| | (0.00) | (0.00) |
| Hispanic | 0.09 | -0.19 |
| | (0.21) | (0.18) |
| Non-Hispanic Black | -0.19 | -0.27 |
| | (0.18) | (0.16) |
| Non-Hispanic Asian | -0.14 | -0.26* |
| | (0.14) | (0.11) |
| Ethnicity-Other/Unknown | -0.24 | -0.10 |
| | (0.19) | (0.15) |
| Missing Demographics | 0.69 | -0.55 |
| | (0.61) | (0.81) |
| Constant | 4.27*** | 5.50*** |
| | (0.15) | (0.12) |
| $N$ | 1987 | 1987 |
| $R^2$ | 0.018 | 0.009 |

Robust standard errors are reported in parentheses

* $p < 0.05$, ** $p < 0.01$, *** $p < 0.001$

*Notes*: Ownership is a dummy variable coded as one when participants received one of the text messages containing the ownership language.

Table 25: Effects of Video on Beliefs and Perceptions

| | Infection_Without | Infection_With | Effectiveness | Worry | Anticipated Regret | Trust |
|---|---|---|---|---|---|---|
| | (1) | (2) | (3) | (4) | (5) | (6) |
| Video | 2.79* | -2.21** | 5.01*** | 0.26** | 0.13 | 0.00 |
| | (1.11) | (0.83) | (1.28) | (0.09) | (0.10) | (0.08) |
| Gender-Female | 8.82*** | 6.18*** | 2.63* | 0.48*** | 0.18 | -0.27*** |
| | (1.12) | (0.83) | (1.27) | (0.09) | (0.10) | (0.08) |
| Gender-Other | 14.79** | 3.40 | 11.39 | 1.21*** | 1.19*** | 0.76* |
| | (4.98) | (3.52) | (6.02) | (0.27) | (0.24) | (0.38) |
| Age | -0.15*** | 0.03 | -0.19*** | -0.01*** | -0.00 | -0.02*** |
| | (0.04) | (0.03) | (0.05) | (0.00) | (0.00) | (0.00) |
| Hispanic | 6.66* | 5.09* | 1.57 | 0.17 | 0.23 | -0.07 |
| | (2.63) | (2.44) | (3.10) | (0.21) | (0.23) | (0.19) |
| Non-Hispanic Black | 1.24 | 7.40*** | -6.16* | -0.21 | 0.01 | -0.59*** |
| | (2.31) | (2.14) | (2.91) | (0.18) | (0.20) | (0.16) |
| Non-Hispanic Asian | 2.38 | 0.96 | 1.42 | 0.27* | 0.72*** | 0.24* |
| | (1.85) | (1.36) | (2.05) | (0.14) | (0.13) | (0.11) |
| Ethnicity-Other/Unknown | -0.70 | 1.61 | -2.31 | -0.06 | 0.30 | -0.14 |
| | (2.55) | (1.82) | (2.60) | (0.20) | (0.20) | (0.17) |
| Missing Demographics | -2.74 | -0.64 | -2.10 | -0.77 | -0.63 | -0.82 |
| | (8.58) | (6.83) | (10.68) | (0.61) | (0.46) | (0.60) |
| Constant | 46.32*** | 12.86*** | 33.46*** | 4.84*** | 5.14*** | 5.05*** |
| | (2.01) | (1.47) | (2.29) | (0.16) | (0.17) | (0.14) |
| *N* | 1992 | 1992 | 1992 | 1991 | 1991 | 1988 |
| $R^2$ | 0.044 | 0.040 | 0.022 | 0.034 | 0.019 | 0.033 |

Robust standard errors are reported in parentheses

* $p < 0.05$, ** $p < 0.01$, *** $p < 0.001$

*Notes*: Video is a dummy variable coded as one when participants received one of the text messages containing the video. Note that we multiplied participants' original responses by 100 for Infection Likelihood without Vaccine (ranging from 0 to 100), Infection Likelihood with Vaccine (ranging from 0 to 100), and Vaccine Effectiveness (ranging from -100 to 100).

the video boosted people's belief that the vaccine could reduce their infection likelihood.

In addition, watching the video led participants to feel more worried about the possibility of transmitting COVID-19 to others (B = 0.26, SE = 0.09, p = 0.003; Column 4). Our video did not significantly change people's anticipated regret of not getting vaccinated (B = 0.13, SE = 0.10, p = 0.17; Column 5) or their trust in the safety and development process of the authorized vaccines (B = 0.003, SE = 0.08, p = 0.97; Column 6).

We also conducted a mediation analysis to explore which of these beliefs and perceptions underlies the effect of the video on scheduling likelihood. We ran a multi-mediator model (model 4 in Hayes 2013) that included *Infection Likelihood without Vaccine*, *Vaccine Effectiveness*, *Worry of Transmission*, *Anticipated Regret*, and *Trust in Vaccine* as potential mediators and kept the aforementioned demographic controls. We used 5,000 bootstrapped samples. We found evidence of partial mediation. Specifically, we estimated a positive indirect effect of *Vaccine Effectiveness* (B = 0.03, SE = 0.01), and the 95% bias-corrected confidence interval (CI) did not include zero ([0.01, 0.06]). We also estimated a positive indirect effect of *Worry of Transmission* (B = 0.015, SE = 0.007), and the 95% bias-corrected CI excluded zero ([0.004, 0.03]). These results, along with the results earlier about differences in beliefs and perceptions between people who watched the video versus did not, suggest that watching the video significantly boosted people's beliefs in the effectiveness of the existing vaccine and elevated their worry of transmission to others, which further increased their reported likelihood of scheduling a vaccination appointment.

##### 2.2.5 Heterogeneous Treatment Effects

Finally, as pre-registered, we explored how the effects of our interventions on scheduling likelihood and persuasiveness differed across the different sub-populations studied. All results are presented in Tables 27-32. In general, we found that estimated treatment effects did not differ statistically significantly whether we looked at participants who identified themselves as Democrats or Republicans, participants who did or did not receive the flu shot at least once during the two most recent seasons, participants with or without a bachelor's degree, or participants who self-reported washing their hands at a high or low frequency when coming back home from outside.[17]

When comparing participants based on the frequency with which they wore masks outside (Table 31), we saw that the simple effect of adding ownership language (i.e., the difference between *Ownership Text* and *Simple Text*) on scheduling likelihood was more negative among people who did not always wear masks than those who always wore masks. Also, the non-additive nature of video and ownership language (in terms of the negative interaction between video and ownership language) was exacerbated among people who always wore masks than those who did not always wear masks. Regarding participants with or without children under 18, the only significant difference is that the positive effect of ownership language without the presence of video (vs. the simple text) on scheduling likelihood was smaller among participants with children than among participants without children.

[17]For the frequency with which people washed their hands when coming back home and the frequency with which people wore masks when outside, we pre-registered that we would do a median split. The median response is 5 for both questions, corresponding to "Always or almost always." Thus, we essentially separated participants into two buckets for each question based on whether or not they reported always washing their hands (or always wearing masks).

Table 26: Main Effects of Text Message on Scheduling Likelihood and Persuasiveness for Democrats and Republicans

| | Scheduling Likelihood | | | Persuasiveness | | |
|---|---|---|---|---|---|---|
| | Democrat | Republican | Both | Democrat | Republican | Both |
| | (1) | (2) | (3) | (4) | (5) | (6) |
| Ownership | -0.03 | -0.24 | -0.24 | 0.24** | 0.09 | 0.09 |
| | (0.10) | (0.15) | (0.15) | (0.09) | (0.11) | (0.11) |
| Video | 0.17 + | 0.32* | 0.31* | 1.49*** | 1.33*** | 1.32*** |
| | (0.10) | (0.15) | (0.15) | (0.09) | (0.11) | (0.11) |
| Democrat | | | 2.07*** | | | 0.63*** |
| | | | (0.15) | | | (0.13) |
| Ownership x Democrat | | | 0.21 | | | 0.15 |
| | | | (0.18) | | | (0.14) |
| Video x Democrat | | | -0.14 | | | 0.18 |
| | | | (0.18) | | | (0.14) |
| Gender-Female | 0.01 | -0.25 | -0.13 | 0.06 | 0.22 | 0.13 |
| | (0.09) | (0.15) | (0.09) | (0.09) | (0.11) | (0.07) |
| Gender-Other | 0.04 | 0.85* | 0.07 | -0.14 | 2.97*** | 0.06 |
| | (0.43) | (0.37) | (0.40) | (0.39) | (0.25) | (0.40) |
| Age | 0.00 | 0.01* | 0.01 | 0.01*** | 0.01** | 0.01*** |
| | (0.00) | (0.01) | (0.00) | (0.00) | (0.00) | (0.00) |
| Hispanic | -0.25 | -0.14 | -0.15 | 0.09 | -0.27 | -0.02 |
| | (0.21) | (0.46) | (0.21) | (0.21) | (0.29) | (0.17) |
| Non-Hispanic Black | -1.36*** | -0.35 | -1.05*** | -0.19 | 0.26 | -0.06 |
| | (0.21) | (0.41) | (0.19) | (0.17) | (0.34) | (0.16) |
| Non-Hispanic Asian | -0.12 | 0.99** | 0.23 | -0.28* | 0.14 | -0.17 |
| | (0.13) | (0.32) | (0.13) | (0.14) | (0.24) | (0.12) |
| Ethnicity-Other/Unknown | -0.45 | 0.27 | -0.09 | -0.29 | -0.25 | -0.25 |
| | (0.24) | (0.36) | (0.21) | (0.19) | (0.25) | (0.15) |
| Missing Demographics | -0.28 | -0.80 | -0.09 | 0.67 | -2.59*** | 0.36 |
| | (0.77) | (0.77) | (0.67) | (0.58) | (0.64) | (0.59) |
| Constant | 6.01*** | 3.49*** | 3.72*** | 3.54*** | 2.87*** | 2.89*** |
| | (0.19) | (0.27) | (0.19) | (0.17) | (0.20) | (0.15) |
| $N$ | 1037 | 942 | 1979 | 1037 | 942 | 1979 |
| $R^2$ | 0.067 | 0.026 | 0.236 | 0.230 | 0.146 | 0.212 |

Robust standard errors are reported in parentheses

$^{+}$ $p < 0.1$, $^{*}$ $p < 0.05$, $^{**}$ $p < 0.01$, $^{***}$ $p < 0.001$

*Notes*: Heterogeneous treatment effects by political affiliation. Columns 1 and 4 focus on participants who self-identified as Democrats in our survey, Columns 2 and 5 focus on participants who self-identified as Republicans in our survey, and Columns 3 and 6 focus on the combined sample (excluding people who did not identify with either party). Video is a dummy variable coded as one when participants received one of the text messages containing the video. Ownership is a dummy variable coded as one when participants received one of the text messages containing the ownership language.

Table 27: Effects of Text Message on Scheduling Likelihood and Persuasiveness for Democrats and Republicans

| | Scheduling Likelihood | | | Persuasiveness | | |
|---|---|---|---|---|---|---|
| | Democrat | Republican | Both | Democrat | Republican | Both |
| | (1) | (2) | (3) | (4) | (5) | (6) |
| Ownership | 0.15 | -0.19 | -0.17 | 0.56*** | 0.37* | 0.37* |
| | (0.14) | (0.21) | (0.21) | (0.14) | (0.16) | (0.16) |
| Video | 0.35** | 0.37 | 0.38 | 1.81*** | 1.60*** | 1.60*** |
| | (0.13) | (0.21) | (0.21) | (0.13) | (0.15) | (0.15) |
| Ownership x Video | -0.36 | -0.09 | -0.14 | -0.64*** | -0.55* | -0.56* |
| | (0.19) | (0.30) | (0.30) | (0.18) | (0.22) | (0.22) |
| Democrat | | | 2.02*** | | | 0.61*** |
| | | | (0.17) | | | (0.15) |
| Ownership x Democrat | | | 0.31 | | | 0.19 |
| | | | (0.25) | | | (0.22) |
| Video x Democrat | | | -0.04 | | | 0.22 |
| | | | (0.25) | | | (0.20) |
| Ownership x Video x Democrat | | | -0.22 | | | -0.09 |
| | | | (0.35) | | | (0.29) |
| Gender-Female | 0.02 | -0.25 | -0.12 | 0.08 | 0.21 | 0.14 |
| | (0.09) | (0.15) | (0.09) | (0.09) | (0.11) | (0.07) |
| Gender-Other | 0.04 | 0.87* | 0.06 | -0.15 | 3.10*** | 0.05 |
| | (0.43) | (0.37) | (0.40) | (0.38) | (0.26) | (0.40) |
| Age | 0.00 | 0.01 | 0.01 | 0.01*** | 0.01** | 0.01*** |
| | (0.00) | (0.01) | (0.00) | (0.00) | (0.00) | (0.00) |
| Hispanic | -0.27 | -0.14 | -0.16 | 0.06 | -0.25 | -0.03 |
| | (0.21) | (0.46) | (0.21) | (0.21) | (0.30) | (0.17) |
| Non-Hispanic Black | -1.36*** | -0.36 | -1.05*** | -0.20 | 0.22 | -0.08 |
| | (0.21) | (0.42) | (0.19) | (0.17) | (0.35) | (0.16) |
| Non-Hispanic Asian | -0.12 | 0.98** | 0.23 | -0.28* | 0.12 | -0.18 |
| | (0.13) | (0.32) | (0.13) | (0.14) | (0.24) | (0.12) |
| Ethnicity-Other/Unknown | -0.44 | 0.27 | -0.08 | -0.27 | -0.26 | -0.24 |
| | (0.23) | (0.37) | (0.21) | (0.19) | (0.25) | (0.15) |
| Missing Demographics | -0.23 | -0.82 | -0.07 | 0.75 | -2.74*** | 0.38 |
| | (0.78) | (0.77) | (0.67) | (0.56) | (0.63) | (0.58) |
| Constant | 5.93*** | 3.47*** | 3.69*** | 3.40*** | 2.75*** | 2.76*** |
| | (0.19) | (0.28) | (0.21) | (0.18) | (0.21) | (0.16) |
| $N$ | 1037 | 942 | 1979 | 1037 | 942 | 1979 |
| $R^2$ | 0.070 | 0.026 | 0.237 | 0.239 | 0.152 | 0.220 |

Robust standard errors are reported in parentheses

* $p < 0.05$, ** $p < 0.01$, *** $p < 0.001$

*Notes*: Heterogeneous treatment effects by political affiliation. Columns 1 and 4 focus on participants who self-identified as Democrats in our survey, Columns 2 and 5 focus on participants who self-identified as Republicans in our survey, and Columns 3 and 6 focus on the combined sample (excluding people who did not identify with either party). Video is a dummy variable coded as one when participants received one of the text messages containing the video. Ownership is a dummy variable coded as one when participants received one of the text messages containing the ownership language.

Table 28: Effects of Text Message on Scheduling Likelihood and Persuasiveness for People who Got vs. Did Not Get the Flu Shot in Two Recent years

| | Scheduling Likelihood | | | Persuasiveness | | |
|---|---|---|---|---|---|---|
| | Flu Shot=0 | Flu Shot=1 | Both | Flu Shot=0 | Flu Shot=1 | Both |
| | (1) | (2) | (3) | (4) | (5) | (6) |
| Ownership | -0.12 | -0.02 | -0.11 | 0.41** | 0.47** | 0.42** |
| | (0.21) | (0.17) | (0.21) | (0.16) | (0.15) | (0.16) |
| Video | 0.30 | 0.39** | 0.30 | 1.60*** | 1.81*** | 1.60*** |
| | (0.20) | (0.15) | (0.20) | (0.15) | (0.13) | (0.15) |
| Ownership x Video | 0.05 | -0.34 | 0.06 | -0.51* | -0.64*** | -0.50* |
| | (0.28) | (0.23) | (0.29) | (0.21) | (0.19) | (0.21) |
| Flu Shot | | | 1.51*** | | | 0.38* |
| | | | (0.18) | | | (0.15) |
| Ownership x Flu Shot | | | 0.11 | | | 0.06 |
| | | | (0.27) | | | (0.22) |
| Video x Flu shot | | | 0.10 | | | 0.20 |
| | | | (0.25) | | | (0.20) |
| Ownership x Video x Flu Shot | | | -0.42 | | | -0.14 |
| | | | (0.37) | | | (0.29) |
| Gender-Female | -0.04 | 0.06 | -0.02 | 0.15 | 0.23* | 0.17* |
| | (0.14) | (0.11) | (0.09) | (0.11) | (0.10) | (0.07) |
| Gender-Other | 0.79 | 1.00** | 0.95* | 0.20 | 0.51 | 0.37 |
| | (0.65) | (0.36) | (0.38) | (0.66) | (0.42) | (0.38) |
| Age | -0.03*** | 0.01 | -0.01** | -0.00 | 0.02*** | 0.01** |
| | (0.01) | (0.00) | (0.00) | (0.00) | (0.00) | (0.00) |
| Hispanic | 0.34 | 0.18 | 0.25 | 0.20 | 0.08 | 0.14 |
| | (0.34) | (0.25) | (0.23) | (0.25) | (0.26) | (0.18) |
| Non-Hispanic Black | -0.65* | -0.22 | -0.45* | -0.06 | 0.37 | 0.14 |
| | (0.26) | (0.24) | (0.18) | (0.23) | (0.20) | (0.15) |
| Non-Hispanic Asian | 0.77*** | 0.40** | 0.48*** | -0.10 | 0.01 | -0.07 |
| | (0.23) | (0.14) | (0.13) | (0.19) | (0.15) | (0.12) |
| Ethnicity-Other/Unknown | -0.29 | 0.03 | -0.13 | -0.72** | 0.17 | -0.23 |
| | (0.33) | (0.25) | (0.20) | (0.24) | (0.18) | (0.15) |
| Missing Demographics | 0.76 | 0.43 | 0.48 | 1.23 | -0.78 | 0.43 |
| | (1.08) | (0.40) | (0.78) | (1.05) | (0.45) | (0.76) |
| Constant | 5.17*** | 5.33*** | 4.45*** | 3.51*** | 3.02*** | 3.07*** |
| | (0.28) | (0.21) | (0.20) | (0.21) | (0.18) | (0.16) |
| $N$ | 1000 | 987 | 1987 | 1000 | 987 | 1987 |
| $R^2$ | 0.052 | 0.020 | 0.144 | 0.155 | 0.226 | 0.192 |

*Notes*: Heterogeneous treatment effects by whether participants reported getting the flu shot in either the 2019-2020 or the 2020-2021 flu season. Columns 1 and 4 focus on participants who got the flu shot, Columns 2 and 5 focus on participants who did not get the flu shot, and Columns 3 and 6 focus on the combined sample. Video is a dummy variable coded as one when participants received one of the text messages containing the video. Ownership is a dummy variable coded as one when participants received one of the text messages containing the ownership language.

Table 29: Effects of Text Message on Scheduling Likelihood and Persuasiveness by Education Level

| | Likelihood of Scheduling | | | Persuasiveness | | |
|---|---|---|---|---|---|---|
| | Below Bachelor | Bachelor or Above | Both | Below Bachelor | Bachelor or Above | Both |
| | (1) | (2) | (3) | (4) | (5) | (6) |
| Ownership | -0.05 | -0.02 | -0.04 | 0.37* | 0.51*** | 0.38* |
| | (0.22) | (0.18) | (0.22) | (0.16) | (0.15) | (0.16) |
| Video | 0.54* | 0.21 | 0.55** | 1.76*** | 1.67*** | 1.77*** |
| | (0.21) | (0.17) | (0.21) | (0.16) | (0.13) | (0.16) |
| Ownership x Video | -0.33 | -0.17 | -0.33 | -0.52* | -0.66*** | -0.53* |
| | (0.31) | (0.25) | (0.31) | (0.22) | (0.19) | (0.22) |
| Bachelor or Above | | | 0.76*** | | | 0.19 |
| | | | (0.19) | | | (0.15) |
| Video x Bachelor or Above | | | -0.33 | | | -0.09 |
| | | | (0.27) | | | (0.21) |
| Ownership x Bachelor or Above | | | 0.03 | | | 0.14 |
| | | | (0.28) | | | (0.22) |
| Ownership x Video x Bachelor or Above | | | 0.13 | | | -0.15 |
| | | | (0.39) | | | (0.30) |
| Gender-Female | -0.08 | 0.10 | 0.00 | 0.18 | 0.17 | 0.17* |
| | (0.16) | (0.12) | (0.10) | (0.11) | (0.10) | (0.07) |
| Gender-Other | 1.48** | 0.38 | 0.96* | 0.35 | 0.41 | 0.38 |
| | (0.53) | (0.59) | (0.43) | (0.47) | (0.63) | (0.39) |
| Age | -0.02** | 0.00 | -0.01* | 0.01 | 0.01* | 0.01** |
| | (0.01) | (0.01) | (0.00) | (0.00) | (0.00) | (0.00) |
| Hispanic | 0.44 | -0.02 | 0.29 | 0.14 | 0.17 | 0.16 |
| | (0.31) | (0.38) | (0.24) | (0.23) | (0.30) | (0.18) |
| Non-Hispanic Black | -0.69* | -0.34 | -0.51** | -0.14 | 0.34 | 0.12 |
| | (0.28) | (0.26) | (0.19) | (0.24) | (0.20) | (0.16) |
| Non-Hispanic Asian | 0.63* | 0.71*** | 0.68*** | -0.06 | 0.03 | -0.00 |
| | (0.25) | (0.15) | (0.13) | (0.21) | (0.14) | (0.12) |
| Ethnicity-Other/Unknown | 0.23 | -0.08 | 0.07 | -0.07 | -0.29 | -0.17 |
| | (0.31) | (0.31) | (0.22) | (0.22) | (0.23) | (0.16) |
| Missing Demographics | 0.00 | 0.00 | 0.00 | 0.00 | 0.00 | 0.00 |
| | (.) | (.) | (.) | (.) | (.) | (.) |
| Constant | 5.03*** | 5.07*** | 4.70*** | 3.16*** | 3.28*** | 3.12*** |
| | (0.29) | (0.25) | (0.22) | (0.21) | (0.19) | (0.16) |
| *N* | 872 | 1110 | 1982 | 872 | 1110 | 1982 |
| $R^2$ | 0.048 | 0.018 | 0.050 | 0.184 | 0.171 | 0.178 |

*Notes*: Heterogeneous treatment effects by whether participants have at least a bachelor's degree. Columns 1 and 4 focus on participants who do not have a bachelor's degree, Columns 2 and 5 focus on participants who have a bachelor's degree or higher, and Columns 3 and 6 focus on the combined sample. Video is a dummy variable coded as one when participants received one of the text messages containing the video. Ownership is a dummy variable coded as one when participants received one of the text messages containing the ownership language.

Table 30: Effects of Text Message on Scheduling Likelihood and Persuasiveness by Handwashing Frequency

| | Scheduling Likelihood | | | Persuasiveness | | |
|---|---|---|---|---|---|---|
| | Handwashing Low | Handwashing High | Both | Handwashing Low | Handwashing High | Both |
| | (1) | (2) | (3) | (4) | (5) | (6) |
| Ownership | 0.02 | -0.11 | 0.02 | 0.35* | 0.51*** | 0.35* |
| | (0.21) | (0.19) | (0.21) | (0.15) | (0.15) | (0.15) |
| Video | 0.48* | 0.24 | 0.49* | 1.68*** | 1.71*** | 1.69*** |
| | (0.20) | (0.18) | (0.20) | (0.14) | (0.14) | (0.14) |
| Ownership x Video | -0.45 | -0.06 | -0.46 | -0.56** | -0.62** | -0.57** |
| | (0.29) | (0.26) | (0.29) | (0.21) | (0.20) | (0.21) |
| Handwashing High | | | 0.56** | | | 0.34* |
| | | | (0.19) | | | (0.15) |
| Ownership x Handwashing High | | | -0.13 | | | 0.16 |
| | | | (0.28) | | | (0.22) |
| Handwashing High x Video | | | -0.24 | | | 0.03 |
| | | | (0.27) | | | (0.20) |
| Ownership x Video x Handwashing High | | | 0.39 | | | -0.06 |
| | | | (0.39) | | | (0.29) |
| Gender-Female | 0.16 | -0.26* | -0.06 | 0.19 | 0.05 | 0.12 |
| | (0.15) | (0.13) | (0.10) | (0.11) | (0.10) | (0.07) |
| Gender-Other | 1.25* | 0.63 | 0.97* | 0.57 | 0.21 | 0.41 |
| | (0.59) | (0.50) | (0.42) | (0.45) | (0.70) | (0.39) |
| Age | -0.01* | -0.01 | -0.01* | -0.00 | 0.01** | 0.01* |
| | (0.01) | (0.01) | (0.00) | (0.00) | (0.00) | (0.00) |
| Hispanic | 0.58 | -0.22 | 0.07 | 0.38 | -0.16 | 0.03 |
| | (0.33) | (0.32) | (0.24) | (0.29) | (0.23) | (0.19) |
| Non-Hispanic Black | -0.37 | -0.75** | -0.59** | 0.18 | -0.03 | 0.05 |
| | (0.33) | (0.25) | (0.20) | (0.26) | (0.20) | (0.16) |
| Non-Hispanic Asian | 0.87*** | 0.49** | 0.62*** | 0.11 | -0.18 | -0.08 |
| | (0.21) | (0.17) | (0.13) | (0.20) | (0.15) | (0.12) |
| Ethnicity-Other/Unknown | -0.09 | -0.04 | -0.06 | -0.31 | -0.22 | -0.26 |
| | (0.38) | (0.27) | (0.22) | (0.27) | (0.19) | (0.16) |
| Missing Demographics | -2.60*** | 1.04 | -0.26 | 0.42 | 1.10 | 0.82 |
| | (0.66) | (0.54) | (1.02) | (0.52) | (1.35) | (0.88) |
| Constant | 4.92*** | 5.54*** | 4.95*** | 3.36*** | 3.34*** | 3.16*** |
| | (0.28) | (0.26) | (0.21) | (0.20) | (0.19) | (0.15) |
| $N$ | 933 | 1052 | 1985 | 933 | 1052 | 1985 |
| $R^2$ | 0.035 | 0.031 | 0.039 | 0.177 | 0.181 | 0.188 |

*Notes*: Heterogeneous treatment effects by whether participant wash their hands frequently after coming back home from outside. The median value of people's self-reported hand-washing frequency is 5, corresponding to "Always or almost always." Columns 1 and 4 focus on participants who self-reported always washing their hands, Columns 2 and 5 focus on participants who self-reported not always washing their hands, and Columns 3 and 6 focus on the combined sample. Video is a dummy variable coded as one when participants received one of the text messages containing the video. Ownership is a dummy variable coded as one when participants received one of the text messages containing the ownership language.

Table 31: Effects of Text Message on Scheduling Likelihood and Persuasiveness by Mask-Wearing Frequency

| | Scheduling Likelihood | | | Persuasiveness | | |
|---|---|---|---|---|---|---|
| | MaskWearing Low | MaskWearing High | Both | MaskWearing Low | MaskWearing Low | Both |
| | (1) | (2) | (3) | (4) | (5) | (6) |
| Ownership | -0.49 | 0.18 | -0.47 | 0.07 | 0.63*** | 0.07 |
| | (0.26) | (0.15) | (0.26) | (0.18) | (0.13) | (0.18) |
| Video | 0.23 | 0.45** | 0.24 | 1.41*** | 1.86*** | 1.40*** |
| | (0.27) | (0.14) | (0.26) | (0.19) | (0.12) | (0.19) |
| Ownership x Video | 0.42 | -0.54** | 0.40 | -0.33 | -0.72*** | -0.32 |
| | (0.37) | (0.20) | (0.36) | (0.27) | (0.17) | (0.27) |
| MaskWearing High | | | 1.61*** | | | 0.34* |
| | | | (0.21) | | | (0.15) |
| Video x MaskWearing High | | | 0.21 | | | 0.45* |
| | | | (0.30) | | | (0.22) |
| Ownership x MaskWearing High | | | 0.64* | | | 0.56* |
| | | | (0.30) | | | (0.22) |
| Ownership x Video x MaskWearing High | | | -0.94* | | | -0.40 |
| | | | (0.42) | | | (0.32) |
| Gender-Female | -0.34 | -0.09 | -0.18 | -0.01 | 0.14 | 0.10 |
| | (0.18) | (0.10) | (0.09) | (0.14) | (0.09) | (0.07) |
| Gender-Other | 2.87*** | 0.25 | 0.33 | -0.53*** | 0.21 | 0.13 |
| | (0.20) | (0.44) | (0.43) | (0.15) | (0.40) | (0.38) |
| Age | -0.01 | -0.01* | -0.01* | 0.00 | 0.01** | 0.01** |
| | (0.01) | (0.00) | (0.00) | (0.01) | (0.00) | (0.00) |
| Hispanic | 0.95 | -0.41 | -0.17 | 0.15 | -0.05 | -0.03 |
| | (0.53) | (0.26) | (0.24) | (0.35) | (0.21) | (0.18) |
| Non-Hispanic Black | -0.38 | -0.86*** | -0.73*** | -0.15 | 0.07 | 0.02 |
| | (0.44) | (0.20) | (0.19) | (0.34) | (0.17) | (0.15) |
| Non-Hispanic Asian | 0.10 | 0.32* | 0.31* | -0.22 | -0.14 | -0.16 |
| | (0.42) | (0.13) | (0.12) | (0.38) | (0.13) | (0.12) |
| Ethnicity-Other/Unknown | 0.47 | -0.23 | -0.01 | -0.23 | -0.21 | -0.21 |
| | (0.43) | (0.24) | (0.22) | (0.29) | (0.18) | (0.15) |
| Missing Demographics | 0.00 | -0.02 | -0.34 | 0.00 | 0.84 | 0.89 |
| | (.) | (1.11) | (1.11) | (.) | (0.90) | (0.89) |
| Constant | 4.20*** | 5.76*** | 4.17*** | 3.33*** | 3.30*** | 3.07*** |
| | (0.36) | (0.20) | (0.23) | (0.24) | (0.17) | (0.16) |
| $N$ | 612 | 1373 | 1985 | 612 | 1373 | 1985 |
| $R^2$ | 0.035 | 0.033 | 0.168 | 0.131 | 0.212 | 0.214 |

*Notes*: Heterogeneous treatment effects by whether participant wear a mask frequently when outside. The median value of people's self-reported mask wearing frequency is 5, corresponding to "Always or almost always." Columns 1 and 4 focus on participants who reported not always wearing masks, Columns 2 and 5 focus on participants who reported always wearing masks, and Columns 3 and 6 focus on the combined sample. Video is a dummy variable coded as one when participants received one of the text messages containing the video. Ownership is a dummy variable coded as one when participants received one of the text messages containing the ownership language.

Table 32: Effects of Text Message on Scheduling Likelihood and Persuasiveness by Whether Participants Have Children

| | Persuasiveness | | | Scheduling Likelihood | | |
|---|---|---|---|---|---|---|
| | Do Not have Children | Have Children | Both | Do Not have Children | Have Children | Both |
| | (1) | (2) | (3) | (4) | (5) | (6) |
| Ownership | 0.11 | -0.82* | 0.10 | 0.57*** | 0.04 | 0.57*** |
| | (0.17) | (0.33) | (0.17) | (0.14) | (0.26) | (0.14) |
| Video | 0.34* | 0.19 | 0.32 | 1.76*** | 1.38*** | 1.75*** |
| | (0.17) | (0.30) | (0.17) | (0.13) | (0.23) | (0.13) |
| Ownership x Video | -0.43 | 0.45 | -0.39 | -0.73*** | -0.06 | -0.71*** |
| | (0.24) | (0.45) | (0.24) | (0.19) | (0.34) | (0.19) |
| Have Children | | | -0.35 | | | 0.22 |
| | | | (0.25) | | | (0.20) |
| Ownership x Have Children | | | -0.82* | | | -0.50 |
| | | | (0.38) | | | (0.29) |
| Video x Have Children | | | -0.04 | | | -0.33 |
| | | | (0.35) | | | (0.27) |
| Ownership x Video xHave Children | | | 0.70 | | | 0.59 |
| | | | (0.51) | | | (0.39) |
| Gender-Female | 0.20 | -0.31 | 0.05 | 0.32*** | -0.03 | 0.22** |
| | (0.12) | (0.22) | (0.11) | (0.09) | (0.17) | (0.08) |
| Gender-Other | 0.75 | 2.48*** | 1.02* | 0.05 | 0.39 | 0.09 |
| | (0.49) | (0.45) | (0.46) | (0.45) | (0.30) | (0.39) |
| Age | -0.01** | 0.01 | -0.01* | 0.00 | 0.01 | 0.01* |
| | (0.00) | (0.01) | (0.00) | (0.00) | (0.01) | (0.00) |
| Hispanic | 0.05 | 0.56 | 0.19 | -0.01 | 0.25 | 0.06 |
| | (0.27) | (0.55) | (0.24) | (0.22) | (0.52) | (0.20) |
| Non-Hispanic Black | -1.07*** | 0.38 | -0.54* | -0.07 | 0.32 | 0.07 |
| | (0.28) | (0.39) | (0.23) | (0.23) | (0.31) | (0.18) |
| Non-Hispanic Asian | 0.42** | 1.71*** | 0.71*** | -0.10 | 0.61* | 0.05 |
| | (0.15) | (0.32) | (0.14) | (0.14) | (0.29) | (0.13) |
| Ethnicity-Other/Unknown | -0.04 | -0.00 | -0.00 | -0.24 | 0.01 | -0.18 |
| | (0.25) | (0.59) | (0.24) | (0.19) | (0.41) | (0.17) |
| Missing Demographics | 0.00 | 0.00 | 0.00 | 0.00 | 0.00 | 0.00 |
| | (.) | (.) | (.) | (.) | (.) | (.) |
| Constant | 5.50*** | 4.19*** | 5.31*** | 3.25*** | 3.23*** | 3.19*** |
| | (0.23) | (0.55) | (0.22) | (0.17) | (0.41) | (0.16) |
| *N* | 1163 | 430 | 1593 | 1163 | 430 | 1593 |
| $R^2$ | 0.039 | 0.072 | 0.053 | 0.184 | 0.148 | 0.168 |

*Notes*: Heterogeneous treatment effects by whether participants reported having children under 18. We did not ask this question to the first 410 participants. Columns 1 and 4 focus on participants who reported not having children under 18, Columns 2 and 5 focus on participants who reported having children under 18, and Columns 3 and 6 focus on the combined sample. Video is a dummy variable coded as one when participants received one of the text messages containing the video. Ownership is a dummy variable coded as one when participants received one of the text messages containing the ownership language.

Table 33: Experiment 1: Effects of Text Messages Likelihood of Scheduling and Persuasiveness

| | Scheduling Likelihood | | Persuasiveness | |
|---|---|---|---|---|
| | (1) | (2) | (3) | (4) |
| Ownership | -0.16 | -0.03 | 0.13 | 0.42** |
| | (0.13) | (0.18) | (0.09) | (0.14) |
| Video | 0.22+ | 0.35+ | 1.43*** | 1.72*** |
| | (0.13) | (0.18) | (0.09) | (0.13) |
| Ownership x Video | | -0.25 | | -0.59** |
| | | (0.26) | | (0.19) |
| Gender-Female | -0.15 | -0.15 | 0.18 | 0.18 |
| | (0.13) | (0.13) | (0.10) | (0.10) |
| Gender-Other | 1.51** | 1.50** | 1.00 | 0.99 |
| | (0.53) | (0.53) | (0.59) | (0.60) |
| Age | 0.00 | 0.00 | 0.01** | 0.01** |
| | (0.01) | (0.01) | (0.00) | (0.00) |
| Hispanic | 0.45 | 0.45 | 0.14 | 0.13 |
| | (0.34) | (0.34) | (0.25) | (0.26) |
| Non-Hispanic Black | -0.22 | -0.23 | 0.21 | 0.20 |
| | (0.24) | (0.24) | (0.19) | (0.19) |
| Non-Hispanic Asian | 0.76*** | 0.75*** | 0.00 | -0.01 |
| | (0.20) | (0.20) | (0.17) | (0.17) |
| Ethnicity-Other/Unknown | -0.33 | -0.32 | -0.43 | -0.41 |
| | (0.33) | (0.34) | (0.24) | (0.24) |
| Missing Demographics | -2.15* | -2.14* | -1.16 | -1.15 |
| | (0.94) | (0.94) | (0.86) | (0.87) |
| Constant | 4.84*** | 4.79*** | 3.22*** | 3.10*** |
| | (0.25) | (0.26) | (0.18) | (0.18) |
| $N$ | 1163 | 1163 | 1163 | 1163 |
| $R^2$ | 0.023 | 0.024 | 0.177 | 0.183 |

Robust standard errors are reported in parentheses

$^{+}$ $p < 0.1$, $^{*}$ $p < 0.05$, $^{**}$ $p < 0.01$, $^{***}$ $p < 0.001$

*Notes*: Video is a dummy variable coded as one when participants received one of the text messages containing the video. Ownership is a dummy variable coded as one when participants received one of the text messages containing the ownership language.

Table 34: Experiment 1: Effects of Ownership Language on Psychological Ownership and Ease of Getting Vaccinated

| | Psychological Ownership | Easiness |
|---|---|---|
| | (1) | (2) |
| Ownership | 0.33** | 0.02 |
| | (0.11) | (0.09) |
| Gender-Female | 0.20 | 0.12 |
| | (0.11) | (0.09) |
| Gender-Other | 0.91 | 0.11 |
| | (0.54) | (0.51) |
| Age | 0.01* | -0.00 |
| | (0.00) | (0.00) |
| Hispanic | -0.10 | 0.00 |
| | (0.30) | (0.20) |
| Non-Hispanic Black | -0.17 | -0.06 |
| | (0.22) | (0.17) |
| Non-Hispanic Asian | -0.13 | -0.19 |
| | (0.19) | (0.15) |
| Ethnicity-Other/Unknown | -0.32 | -0.26 |
| | (0.27) | (0.24) |
| Missing Demographics | 1.12* | 0.76 |
| | (0.53) | (0.49) |
| Constant | 3.92*** | 5.43*** |
| | (0.21) | (0.16) |
| $N$ | 1155 | 1155 |
| $R^2$ | 0.020 | 0.005 |

Robust standard errors are reported in parentheses

* $p < 0.05$, ** $p < 0.01$, *** $p < 0.001$

*Notes*: Ownership is a dummy variable coded as one when participants received one of the text messages containing the ownership language.

Table 35: Experiment 1: Effects of Video on Beliefs and Perceptions

| | Infection_Without | Infection_With | Effectiveness | Worry | Anticipated Regret | Trust |
|---|---|---|---|---|---|---|
| | (1) | (2) | (3) | (4) | (5) | (6) |
| Video | 3.41* | -1.65 | 5.06** | 0.29* | 0.12 | -0.05 |
| | (1.44) | (1.12) | (1.69) | (0.12) | (0.13) | (0.10) |
| Gender-Female | 10.17*** | 7.58*** | 2.58 | 0.61*** | 0.11 | -0.43*** |
| | (1.45) | (1.11) | (1.67) | (0.12) | (0.13) | (0.11) |
| Gender-Other | 23.12*** | 8.04 | 15.08 | 1.77*** | 1.52*** | 1.14** |
| | (7.00) | (5.68) | (9.22) | (0.34) | (0.32) | (0.36) |
| Age | -0.09 | -0.04 | -0.05 | -0.01* | 0.01 | -0.01 |
| | (0.06) | (0.05) | (0.07) | (0.00) | (0.01) | (0.00) |
| Hispanic | 6.53 | 4.79 | 1.75 | -0.07 | 0.15 | 0.11 |
| | (3.44) | (3.23) | (4.83) | (0.29) | (0.36) | (0.29) |
| Non-Hispanic Black | 0.95 | 7.72** | -6.77* | -0.09 | 0.27 | -0.43* |
| | (2.71) | (2.63) | (3.25) | (0.22) | (0.23) | (0.20) |
| Non-Hispanic Asian | 2.47 | 1.82 | 0.65 | 0.30 | 0.73*** | 0.26 |
| | (2.52) | (2.01) | (3.00) | (0.18) | (0.20) | (0.16) |
| Ethnicity-Other/Unknown | -4.75 | 1.01 | -5.76 | -0.45 | -0.09 | -0.53* |
| | (3.54) | (2.55) | (3.90) | (0.30) | (0.31) | (0.24) |
| Missing Demographics | -3.07 | 17.89** | -20.96* | 0.26 | -0.49 | -1.89*** |
| | (7.60) | (5.88) | (9.39) | (0.37) | (0.34) | (0.40) |
| Constant | 42.18*** | 15.05*** | 27.13*** | 4.55*** | 4.72*** | 4.74*** |
| | (2.75) | (2.09) | (3.23) | (0.22) | (0.23) | (0.19) |
| $N$ | 1155 | 1155 | 1155 | 1155 | 1155 | 1155 |
| $R^2$ | 0.053 | 0.052 | 0.018 | 0.040 | 0.015 | 0.033 |

Robust standard errors are reported in parentheses

* $p < 0.05$, ** $p < 0.01$, *** $p < 0.001$

*Notes*: Video is a dummy variable coded as one when participants received one of the text messages containing the video.

Table 36: Experiment 2: Effects of Text Messages Likelihood of Scheduling and Persuasiveness

| | Scheduling Likelihood | | Persuasiveness | |
|---|---|---|---|---|
| | (1) | (2) | (3) | (4) |
| Ownership | -0.12 | 0.07 | 0.20 | 0.54** |
| | (0.15) | (0.22) | (0.11) | (0.17) |
| Video | 0.30* | 0.49* | 1.40*** | 1.73*** |
| | (0.15) | (0.20) | (0.11) | (0.16) |
| Ownership x Video | | -0.38 | | -0.68** |
| | | (0.29) | | (0.23) |
| Gender-Female | 0.17 | 0.19 | 0.14 | 0.16 |
| | (0.15) | (0.15) | (0.11) | (0.11) |
| Gender-Other | 0.31 | 0.31 | -0.30 | -0.29 |
| | (0.62) | (0.65) | (0.44) | (0.43) |
| Age | -0.02** | -0.02** | 0.01 | 0.01 |
| | (0.01) | (0.01) | (0.00) | (0.00) |
| Hispanic | -0.11 | -0.12 | 0.11 | 0.10 |
| | (0.33) | (0.33) | (0.26) | (0.26) |
| Non-Hispanic Black | -1.11** | -1.13** | 0.00 | -0.03 |
| | (0.35) | (0.35) | (0.28) | (0.28) |
| Non-Hispanic Asian | 0.67*** | 0.67*** | 0.03 | 0.03 |
| | (0.17) | (0.17) | (0.17) | (0.17) |
| Ethnicity-Other/Unknown | 0.29 | 0.29 | -0.01 | -0.02 |
| | (0.28) | (0.28) | (0.20) | (0.20) |
| Missing Demographics | 0.26 | 0.26 | 1.18 | 1.17 |
| | (0.88) | (0.88) | (0.63) | (0.61) |
| Constant | 5.43*** | 5.34*** | 3.49*** | 3.32*** |
| | (0.27) | (0.28) | (0.20) | (0.22) |
| $N$ | 840 | 840 | 840 | 840 |
| $R^2$ | 0.049 | 0.051 | 0.163 | 0.172 |

*Notes*: Video is a dummy variable coded as one when participants received one of the text messages containing the video. Ownership is a dummy variable coded as one when participants received one of the text messages containing the ownership language.

Table 37: Experiment 2: Effects of Ownership Language on Psychological Ownership and Ease of Getting Vaccinated

| | Psychological Ownership | Easiness |
|---|---|---|
| | (1) | (2) |
| Enhanced | 0.46*** | -0.02 |
| | (0.13) | (0.11) |
| Gender-Female | 0.28* | 0.26* |
| | (0.13) | (0.11) |
| Gender-Other | 0.28 | 0.50 |
| | (0.48) | (0.29) |
| Age | -0.00 | -0.00 |
| | (0.00) | (0.00) |
| Hispanic | 0.20 | -0.39 |
| | (0.29) | (0.29) |
| Non-Hispanic Black | -0.13 | -0.70* |
| | (0.31) | (0.32) |
| Non-Hispanic Asian | -0.18 | -0.36* |
| | (0.20) | (0.16) |
| Ethnicity-Other/Unknown | -0.21 | 0.04 |
| | (0.26) | (0.17) |
| Missing Demographics | 0.74 | -1.21 |
| | (0.79) | (0.96) |
| Constant | 4.54*** | 5.57*** |
| | (0.21) | (0.17) |
| $N$ | 832 | 832 |
| $R^2$ | 0.025 | 0.025 |

Robust standard errors are reported in parentheses

* $p < 0.05$, ** $p < 0.01$, *** $p < 0.001$

*Notes*: Ownership is a dummy variable coded as one when participants received one of the text messages containing the ownership language.

Table 38: Experiment 2: Effects of Video on Beliefs and Perceptions

| | Infection_Without | Infection_With | Effectiveness | Worry | AnticipatedRegret | Trust |
|---|---|---|---|---|---|---|
| | (1) | (2) | (3) | (4) | (5) | (6) |
| Video | 1.82 | -2.84* | 4.67* | 0.20 | 0.13 | 0.07 |
| | (1.75) | (1.26) | (1.94) | (0.13) | (0.14) | (0.12) |
| Gender-Female | 6.80*** | 4.48*** | 2.32 | 0.30* | 0.25 | -0.07 |
| | (1.77) | (1.27) | (1.96) | (0.14) | (0.15) | (0.12) |
| Gender-Other | 6.33 | -1.71 | 8.04 | 0.72 | 0.92* | 0.42 |
| | (6.36) | (2.41) | (7.06) | (0.39) | (0.38) | (0.69) |
| Age | -0.22** | 0.09 | -0.31*** | -0.02*** | -0.02** | -0.03*** |
| | (0.07) | (0.05) | (0.07) | (0.01) | (0.01) | (0.00) |
| Hispanic | 6.26 | 5.62 | 0.64 | 0.34 | 0.24 | -0.28 |
| | (3.95) | (3.60) | (4.03) | (0.30) | (0.29) | (0.26) |
| Non-Hispanic Black | 2.69 | 6.34 | -3.65 | -0.40 | -0.50 | -0.94*** |
| | (4.39) | (3.68) | (5.97) | (0.34) | (0.37) | (0.25) |
| Non-Hispanic Asian | 2.06 | 0.44 | 1.62 | 0.22 | 0.66*** | 0.17 |
| | (2.72) | (1.80) | (2.74) | (0.20) | (0.18) | (0.16) |
| Ethnicity-Other/Unknown | 2.63 | 2.09 | 0.55 | 0.28 | 0.65* | 0.20 |
| | (3.66) | (2.62) | (3.44) | (0.26) | (0.27) | (0.23) |
| Missing Demographics | 0.31 | 1.12 | -0.81 | -0.91 | -0.74 | -0.53 |
| | (10.34) | (6.75) | (12.10) | (0.74) | (0.61) | (0.87) |
| Constant | 50.87*** | 11.44*** | 39.43*** | 5.16*** | 5.56*** | 5.33*** |
| | (2.96) | (2.11) | (3.26) | (0.23) | (0.25) | (0.20) |
| $N$ | 837 | 837 | 837 | 836 | 836 | 833 |
| $R^2$ | 0.039 | 0.034 | 0.035 | 0.035 | 0.041 | 0.058 |

Robust standard errors are reported in parentheses

* $p < 0.05$, ** $p < 0.01$, *** $p < 0.001$

*Notes*: Video is a dummy variable coded as one when participants received one of the text messages containing the video.

### 3 Online Survey Comparing Beliefs and Perceptions Across People with Different Vaccination Intentions

We conducted an online survey in January 2021 to explore factors predicting the general public's vaccination intentions.

#### 3.1 Method

##### 3.1.1 Participants

Given that our RCTs targeted patients at UCLA Health, most of whom live in California, we aimed to recruit 500 survey respondents living in California. To speed up data collection and get a relatively more diverse sample, we recruited survey respondents from both MTurk and Prolific and used these platforms' location screening tools to identify California residents. Our final sample consists of 515 participants (48.93% female, M_age = 33.91). If a person took the survey more than once, we kept their first complete response. Participants received $1.00 on MTurk or $1.20 on Prolific for completing our 9-minute survey.

##### 3.1.2 Procedure

At the introduction, we informed participants that they should consider the RNA vaccines developed by Pfizer-BioNTech and Moderna (both of which had been authorized by the FDA for emergency use) when answering questions about COVID-19 vaccines during the survey. We collected participants' vaccination intentions, beliefs, and perceptions about COVID-19, attitudes toward the authorized COVID-19 vaccines, general preventative measures, and demographics. The complete survey is available at `https://osf.io/qn8hr/?view_only=cf7b2bc590054aee8c4a2bae99ef20c5`.

- *Vaccination Intentions*: Following a popular national survey conducted by Pew Research Center (Funk and Tyson, 2020), we measured participants' vaccination intentions by asking, "If one of the COVID-19 vaccines were available to you today, would you get the vaccine?" Participants chose one from four options: "Definitely would get the vaccine," "Probably would get the vaccine," "Probably would not get the vaccine," "Definitely would not get the vaccine." We asked participants to explain why they were unsure about whether to get the vaccine (if they indicated they probably would or would not get it) or why they would definitely not get the vaccine (if they indicated they would definitely not get it).

- *Prediction about Others' Intentions*: We told participants that a large sample of US adults were recently surveyed about their vaccination intentions. We asked participants to predict the percentage of U.S. adults who indicated that they would definitely or probably get the vaccine (from 0% to 100%).

- *Beliefs about Herd Immunity*: Participants indicated the percentage of Americans they thought need to get the COVID-19 vaccine in order to have society protected by herd immunity (from 0% to 100%).[18]

- *Infection Likelihood* and *Vaccine Effectiveness*: We asked participants to report their likelihood of getting infected with COVID-19 if they did not get the COVID-19 vaccine (*Infection Likelihood without*

[18]The order in which participants reported the three aforementioned questions (about their vaccination intentions, others' intentions, and herd immunity) versus the following questions about their beliefs and perceptions about COVID-19 and the authorized vaccines were counterbalanced. The order did not make a meaningful difference and thus will not be discussed.

*Vaccine*) and if they got the COVID-19 vaccine (*Infection Likelihood with Vaccine*) (from 0% = I certainly won't get infected with COVID-19 to 100% = I certainly will get infected with COVID-19). For each participant, we subtracted *Infection Likelihood with Vaccine* from *Infection Likelihood without Vaccine* to capture her belief in the effectiveness of the COVID-19 vaccine in reducing her chance of getting infected with the COVID-19. The value of this variable in theory could range from -100% to 100%.

- *Infection Severity*: We used two measures to capture people's beliefs in the severity of their symptoms if they got infected with COVID-19: "In the unfortunate scenario you became infected with COVID-19, how ill do you think you would be?" (from 0 = No symptoms to 10 = Severely ill/hospitalized) and "In the unfortunate scenario you became infected with COVID-19, how confident are you that you can fully recover from it?" (from 0 = Not at all confident to 10 = Extremely confident). Participants' responses to these two questions were significantly correlated ($r = -0.59$, $p < 0.0001$) and thus were averaged (after the second item was reverse coded) to form a composite score of *Infection Severity.*

- *Vulnerability*: Participants rated the extent to which they agreed with the statement, "Without getting the COVID-19 vaccine, I would feel very vulnerable to COVID-19" (from 0 = Strongly disagree to 10 = Strongly agree).

- *Fear of Infections*: Participants rated the extent to which they agreed with the statement, "I am scared of getting COVID-19" (from 0 = Strongly disagree to 10 = Strongly agree).

- *Worry of Transmission*: Participants rated the extent to which they agreed with the statement, "The possibility of passing COVID-19 to other people worries me" (from 0 = Strongly disagree to 10 = Strongly agree).

- *Anticipated Regret*: Participants rated the extent to which they agreed with the statement, "I would regret not getting a COVID-19 vaccine if I end up getting infected with COVID-19" (from 0 = Strongly disagree to 10 = Strongly agree).

- *Trust in Vaccine*: We measured trust in the safety and development process of the vaccine by asking participants to rate their agreement with seven statements (from 0 = Strongly disagree to 10 = Strongly agree). The statements captured worries about side effects ("I am concerned about potential side effects of the COVID-19 vaccine"), the possibility of getting COVID-19 from the vaccine ("I worry that the COVID-19 vaccine will lead me to be infected with COVID-19"), and potential negative long-term impacts of the vaccine ("I worry that the COVID-19 vaccine will have long-term negative impacts on my health"). The statements also captured concerns about the speed of the vaccine authorization process ("I am concerned that the current COVID-19 vaccines were approved too quickly")[19], overall trust in the development of the vaccine ("I trust the research and development process of COVID-19 vaccines"), perceived safety of the vaccine ("I am confident that the research and development process of COVID-19 vaccine has produced a safe vaccine"), and hesitancy about the vaccine ("Before feeling comfortable to get the COVID-19 vaccine myself, I would want to wait for enough people to get the vaccine."). Participants' responses to these seven questions reached a high inter-item reliability (Cronbach's alpha = 0.91); thus, we averaged responses to these questions (after reverse coding responses to the negatively framed statements) to form a composite score of *Trust in Vaccine*. The higher one's composite score is, the more they trusted the safety of vaccine and its development process.

- *Vaccination Importance*: To evaluate how important getting vaccinated was to participants, we had them indicate agreement with four statements (from 0 = Strongly disagree to 10 = Strongly agree): (1) "Getting the COVID-19 vaccine is essential for my life to go back to normal"; (2) "Having enough people get the COVID-19 vaccine is essential for society to go back to normal"; (3) "Getting the COVID-19 vaccine is important for protecting myself"; and (4) "Getting the COVID-19 vaccine is important for protecting others around me." Participants' responses to these four questions reached a high inter-item reliability (Cronbach's alpha = 0.93); thus, we averaged responses to these questions to form a composite score of *Vaccination Importance*. The higher one's composite score is, the more they viewed vaccination as important for themselves and the society.

- *Prevalence of COVID-19*: To assess participants' beliefs in the prevalence of COVID-19, we asked, "How many Americans do you think, on average, test positive for COVID-19 every day in January 2021?" and "How many Americans do you think, on average, die from COVID-19 every day in January 2021?" For both questions, participants entered a number.

- *Personal Experience with COVID-19*: Participants reported whether they had ever been tested for COVID-19 and if yes, whether they had ever tested positive. Those who tested positive were further asked about what happened to them.

- *Demographics*: Participants reported their age, gender, race/ethnicity, highest level of education completed, pre-tax household income in 2020, and the state they were currently living in.[20] Participants also reported the political party they identified the most with ("The Democratic Party", "The Republican Party", "I am independent", and "Other") and those who chose "I am independent" were further asked to indicate if they identified relatively more with the Democrat Party or the Republican Party or if they identified equally with both parties. We treat people who identified more with the Democratic (Republican) Party (regardless of whether they initially chose "I am independent") as Democrats (Republicans).

- *Second-hand Experience with COVID-19*: Participants reported if they personally knew anyone who was hospitalized or died as a result of COVID-19 and if they had ever been in close contact with someone who had COVID-19 at the time of the contact.

- *Familiarity with Authorized Vaccines*: Participants reported their familiarity for each of the authorized vaccines at the time the study was conducted (i.e., the Pfizer-BioNTec COVID-19 vaccine and the Moderna COVID-19 vaccine).

- *Preventative Measures*: Participants rated how often they wore a mask when going outside as well as how often they washed their hands when coming home from outside (from 1 = Never or almost never

[19]We realized later that we should have asked whether people were concerned that the vaccines were *authorized for use*, rather than *approved*, too quickly. Since this item was highly correlated with the other six items and the general population may not know the difference between FDA's approval and FDA's emergency use authorization, we kept this item. We corrected this mistake in the same measure for our online experiments, as reported in Section 2.

[20]Since 98% of participants reported that they indeed were living in California, we did not exclude participants based on their state of residence.

Table 39: Vaccination Intentions Overall and by Political Affiliation and Race/Ethnicity

| Vaccination Intentions | Full Sample | Democrats | Republicans | Other Political Orientation | |
|---|---|---|---|---|---|
| Definitely Yes | 51.84% | 62.19% | 20.31% | 31.40% | |
| Probably Yes | 28.93% | 27.12% | 43.75% | 25.58% | |
| Probably No | 11.07% | 8.22% | 15.63% | 19.77% | |
| Definitely No | 8.16% | 2.47% | 20.31% | 23.26% | |
| Sample | 515 | 365 | 64 | 86 | |
| Vaccination Intentions | Non-Hispanic White | Non-Hispanic African American | Hispanic | Non-Hispanic Asian | Other |
| Definitely Yes | 56.56% | 21.05% | 35.56% | 52.22% | 56.00% |
| Probably Yes | 23.98% | 52.63% | 31.11% | 33.33% | 24.00% |
| Probably No | 9.50% | 15.79% | 22.22% | 9.44% | 12.00% |
| Definitely No | 9.95% | 10.05% | 11.11% | 5.00% | 4.00% |
| Sample | 221 | 19 | 45 | 230 | 50 |

to 5 = Always or almost always). They also reported whether they got the flu shot in the current (2020-2021) flu season.

#### 3.2 Results

**Descriptive Statistics**. We first report summary statistics about participants' vaccination intentions, predictions about others' intentions, belief about herd community, general familiarity with the vaccine, and experiences with COVID-19. In our sample, 51.84% of participants reported that they definitely would get the vaccine if it was available to them today, 28.93% reported that they probably would get the vaccine, 11.07% probably would not get the vaccine, and the remaining 8.16% definitely would not get it. Consistent with results of recent national surveys among US adults (Funk and Tyson, 2020; Hamel et al., 2020; Ruiz and Bell, 2021), we found that Democrats reported stronger intentions to get the vaccine than Republicans and that both non-Hispanic African American and Hispanic participants indicated weaker intentions than non-Hispanic white participants (see Table 39).[21]

On average, participants predicted that 62.5% of American adults, when surveyed about their vaccination intentions, indicated they would definitely or probably get the vaccine (SD = 15.36%). This prediction is fairly accurate, relative to the actual percentage of American adults who indicated definitely or probably yes when surveyed by the Pew Research CenterFunk and Tyson (2020). Participants on average estimated that 76% of people need to get vaccinated to reach herd immunity (SD = 20.52%). Participants tend to underestimate the prevalence and severity of COVID-19 infections. Specifically, among our survey respondents, the

[21] In an OLS regression with robust standard errors that predicts intent to vaccinate (1 = definitely yes, 2 = probably yes, 3 = probably no, 4 = definitely no) as a function of political affiliation, the difference between Democrats and Republicans is statistically significant ($p < 0.0001$). And an OLS regression with robust standard errors that predicts intentions as a function of race/ethnicity shows that the difference between Hispanic and non-Hispanic white participants was significant ($p = 0.03$) and that between non-Hispanic African American participants and non-Hispanic white participants was also significant ($p = 0.04$). The results are robust if we use OLS or logistics regressions to predict a binary indicator for whether or not participants definitely would get the vaccine.

median estimate of daily confirmed cases in the U.S. in January 2021 was 30,000, and the median estimate of daily deaths caused by COVID-19 in the U.S. in January 2021 was 2,000; both were much lower than the actual numbers.[22]

People were similarly familiar with the Pfizer-BioNTech and Moderna vaccines: 7.06% of participants had not heard about the Pfizer-BioNTech vaccine, 53.53% had heard about it but had not read articles and information about it closely, 38.24% had closely read articles and information about it, and 1.18% had personal experiences with it (e.g., had received the vaccine or administered it to others as a healthcare worker); the choice share for the Moderna vaccine was 6.67%, 58.04%, 33.73%, and 1.57%, respectively.

Most (68.62%) of participants had not been tested for COVID-19. Among those who had been tested, 7.5% tested positive. About 30% of participants knew someone who were hospitalized or died as a result of COVID-19, and 13% had ever been in close contact with someone who had COVID-19 at the time of the contact.

**Beliefs and Perceptions**. Next, we compare beliefs and perceptions about COVID-19 and the authorized vaccines across participants based on whether they definitely would get the vaccine, were uncertain, or definitely would not get the vaccine. Table 40 represents the mean and standard deviation of each measure for each of the three groups of participants. Given that we were primarily interested in developing educational materials to shift the intentions of people who were uncertain, we focus on performing statistical tests to compare people definitely would get the vaccine with those who were uncertain. Compared to those who were uncertain, people who definitely would get the vaccine believed they were more likely to get infected with COVID-19 if they did not get the vaccine and they would be less likely to get infected with COVID-19 if they got the vaccine, believed more strongly in the vaccine's effectiveness, felt more vulnerable to COVID-19, were more scared of getting infected, were more worried about transmitting the virus to others, anticipated feeling a greater degree of regret if they passed on the opportunity to get vaccinated and ended up being infected, and trusted the safety and development process of the vaccine more. The two groups of participants did not differ in their beliefs about how severe their symptoms would be if they got infected with COVID-19.

**The COVID-19 Video**. Based on these results, we decided to create a video with two major components. We aimed to first highlight the threat of COVID-19 (i.e., illustrating the problem) and to then immediately propose vaccination as an easy solution. To that end, the first part of the video highlights the prevalence of COVID-19 (e.g., by presenting the number of confirmed cases in Los Angeles and worldwide) and the virus's ease of transmission. Our goal was to make viewers feel that their chance of being infected is high, correcting potential misconceptions about their likelihood of getting infected and transmitting the virus to others, and increasing feelings of vulnerability and worry of spreading the virus. The second part of the video presents vaccination as an easy solution by reassuring viewers about the safety and effectiveness of the authorized vaccines. Specifically, we explain that the authorized vaccines have gone through rigorous evaluations after large-scale clinical trials involving volunteers of all genders/ethnicities/ages/occupations and that these trials have found that the authorized vaccines can reduce symptomatic COVID by up to 95%. The goal of this portion of the video is to shift beliefs about the effectiveness of the vaccine and instill a

[22] According to `https://covid.cdc.gov/covid-data-tracker/#trends_dailytrendscases`, the average daily number of COVID-19 cases and deaths in the US reported to the Centers for Disease Control and Prevention was 233,181 and 3,002, respectively, during January 1-January 12, 2021 (the day prior to our survey).

Table 40: Comparisons of COVID-19-related Beliefs and Perceptions by Vaccination Intentions

| Measure | Definitely Yes | Uncertain | Definitely No | Definitely Yes vs. Uncertain (p-value of t-test and Cohen's d) |
|---|---|---|---|---|
| Infection Likelihood w/ Vaccine | 57.47% (24.61%) | 48.14% (24.97%) | 30.78% (26.93%) | p<0.0001, d=0.38 |
| Infection Likelihood w/o Vaccine | 16.11% (20.75%) | 19.91% (20.16%) | 37.60% (31.94%) | p=0.047, d=0.19 |
| Vaccine Effectiveness | 41.36% (29.13%) | 28.24% (25.35%) | -6.81% (36.12%) | p<0.0001, d=0.48 |
| Infection Severity | 4.92 (2.14) | 4.61 (2.16) | 2.95 (2.32) | p=0.12, d=0.14 |
| Vulnerability | 8.13 (2.20) | 6.26 (2.78) | 2.21 (3.12) | p<0.0001, d=0.76 |
| Fear of Infections | 7.51 (2.62) | 6.47 (2.97) | 2.90 (3.53) | p<0.0001, d=0.37 |
| Worry of Transmission | 8.61 (2.00) | 7.69 (2.67) | 2.83 (3.52) | p<0.0001, d=0.40 |
| Anticipated Regret | 9.10 (1.82) | 7.21 (2.69) | 1.26 (2.46) | p<0.0001, d=0.85 |
| Trust in Vaccine | 8.46 (1.61) | 5.56 (1.83) | 2.74 (1.45) | p<0.0001, d=1.70 |

Note: We report the mean value and standard deviation (in parentheses) for each measure by intention type.

sense of trust in the vaccine. In addition, the solution part of our video intends to elicit anticipated regret by truthfully telling viewers that 80% of California residents in a recent survey said they would regret not being vaccinated if they ended up getting infected. The number 80% came from this survey where we found that 78.56% of participants gave a rating above the midpoint of the scale in response to the question about anticipated regret.
